# Calcification of abdominal aorta is a high risk underappreciated cardiovascular disease factor in a general population

**DOI:** 10.1101/2020.05.07.20094706

**Authors:** Anurag Sethi, Leland Taylor, J Graham Ruby, Jagadish Venkataraman, Elena Sorokin, Madeleine Cule, Eugene Melamud

## Abstract

**Background:** Abdominal aortic calcification (AAC) is an important contributor to cardiovascular disease, however, prevalence of the pathology, risk factors, and disease outcomes in a general population have not been systematically analyzed.

**Methods:** We have analyzed the prevalence of AAC in the UK Biobank cohort using machine learning models across 38,264 whole body dual-energy X-ray absorptiometry (DEXA) scans. We associated severity of calcification across a wide range of physiological parameters, clinical biomarkers, and environmental risk factors (406 in total). We further performed a rare and common variant genetic association study to identify genetic loci relevant to AAC and evaluated the prognostic value of AAC across 174 disease classes.

**Results:** We find that AAC is highly prevalent in a general population (>10.4% of the cohort) despite low prevalence of diabetes (2.5%) and CKD (0.5%). Increased level of AAC is a strong prognostic indicator of a wide range of cardiovascular outcomes, including stenosis of precerebral arteries (HR∼1.5), myocardial infarction (HR∼1.36), ischemic heart disease (HR∼1.3), and chronic obstructive pulmonary disease (HR∼1.3). We report four genetic loci associated with AAC, three of which have not been previously reported. Surprisingly, we find that elevated but still within clinically normal levels of serum phosphate, serum calcium, and glycated hemoglobin are linked to increased vascular calcification.

**Conclusions:** By our estimate, AAC is an LDL-independent risk factor for cardiovascular outcomes, with risk similar to elevated LDL. Development of anti-calcification therapeutics would complement lipid lowering strategies in reducing CVD burden.

## Introduction

Calcification of the vasculature is a commonly observed pathology in patients with end stage renal disease (ESRD) and diabetes ^1^. At least two distinct forms of vascular calcification are known to exist - intimal layer calcification associated with atherosclerotic plaque and medial layer calcification internal to the tunica media layer of blood vessels. Intimal layer calcification is attributed to inflammatory hypercholesterolemia and calcification of plaque deposits, while medial layer calcification is attributed to growth of hydroxyapatite crystals within the blood vessel walls as a consequence of hyperphosphatemia and/or hypercalcemia ^2,3^.

Although calcification can occur in all major arterial blood vessels, the prevalence of this pathology is site specific, and due to lack of routine monitoring, progression of calcification across various sites is hard to ascertain. In populations with high to moderate cardiovascular risk, calcification is most commonly monitored in the coronary artery via computed tomography measurement and in the carotid artery via ultrasound measurement. At both sites, calcification has been linked to the degree of atherosclerosis and severity of calcification was found to be a prognostic indicator of poor cardiovascular outcomes. ^4–6^.

In this study, we present the largest epidemiological and genetic analysis of AAC to date. AAC has been shown to be superior to calcification in other arteries in predicting mortality for peritoneal dialysis patients^7^. Using machine learning (ML) models, we developed an automated method to quantify the level of AAC in 29,957 DEXA scans from the UK Biobank (UKBB) cohort. This cohort is biased towards healthier volunteers within middle age to elderly UK population but can be used to study associations between different risk factors and diseases ^8^. To date more than 30,000 DEXA images have been collected and up to 100,000 will be collected in the next five years.

Leveraging the extensive measures of biomarkers and disease outcomes in UKBB, we estimate the contribution of hyperlipidemia, hyperphosphatemia, hypercalcemia, diabetes, and kidney health to the severity of AAC ^9^. We utilize genotype data to identify and characterize genetic loci associated with AAC. Finally, using prospective electronic records, we report the association of AAC with risk of development of the most common diseases over an eight-year median follow up time.

Our analysis suggests that AAC arises as a consequence of transformation of arterial tissue into an osteogenic-like state, due to a combination of genetic drivers of vascular osteogenesis and age dependent physiological imbalances in the calcium-phosphate regulatory network. We find that calcification of abdominal aorta is a strong prognostic indicator of cardiovascular morbidity and show that this pathology can occur in a population not enriched for specific disease states.

## Results

### Baseline Characteristics of Cohort

The UKBB imaging study aims to generate detailed images on 100,000 of the 502,604 total UKBB participants by 2023 ^10^. These images span a variety of modalities including DEXA scans of the whole body and targeted regions such as the upper torso. From 2014 to August 2020, 48,705 participants have participated in the imaging cohort. In the latest (February 2020) release, DEXA scans were released for 40,200 participants. We were able to quantify AAC on 38,264 participants during the first imaging visit, herein referred to as the “calcification subcohort.” In addition to the different biochemical and physiological measurements made during visits to the assessment center, the UKBB also releases both retrospective and prospective data on health outcomes derived from electronic health records.

We characterized the entire UKBB cohort and these 38,264 participants across a variety of measures ascertained at time of recruitment and, when available, at time of imaging (∼8 years after initial recruitment; Table 1). Basic anthropometric and clinical measures revealed few differences between the calcification subcohort and the entire UKBB cohort.

**Table 1:** Baseline characteristics of the UK Biobank (UKBB) cohort and the calcification subcohort. The following second level ICD10 codes were used to identify the number of participants with different comorbidities: E10 - Type 1 Diabetes (T1D), E11 - Type 2 Diabetes (T2D), I25 - Myocardial Infarction (MI), I64 - Stroke, I21 - Coronary Artery Disease (CAD), and N18 - Chronic Kidney Disease (CKD). BMI - body mass index, SBP - systolic blood pressure, HbA1c - glycated haemoglobin, Trig - triglycerides, LDL - low density lipoprotein, HDL - high density lipoprotein. The calcification cohort revisited the centers for imaging ∼8 years after the baseline visit for all UKBB cohort participants.

|  | UKBB Cohort<br>(baseline) | Calcification subcohort<br>(baseline) | Calcification subcohort<br>(imaging baseline) |
| --- | --- | --- | --- |
| n | 502,604 | 29,957 | **38,264** |
| Age (years) | 56.3 (8.1) | 54.9 (7.5) | **63.6 (7.6)** |
| % Females | 54.40% | 51.10% | **48.26%** |
| BMI | 27.43 (4.80) | 26.63 (4.23) | **26.56 (4.41)** |
| SBP | 139 (20) | 137 (19) | **138 (18.5)** |
| Pulse | 68 (11.7) | 68.0 (11.0) | **69.0 (12.1)** |
| Smoker (current) | 10.50% | 6.50% | **3.55%** |
| Smoker (previous) | 34.50% | 32.50% | **33.02%** |
| **Comorbidities** |  |  |  |
| % Hypertension | 7.88% | 4.13% | **12.80%** |
| % T1D | 0.42% | 0.15% | **0.28%** |
| % T2D | 1.83% | 0.71% | **2.48%** |
| % MI | 2.12% | 1.12% | **3.62%** |
| % Stroke | 0.14% | 0.07% | **0.10%** |
| % CAD | 0.99% | 0.57% | **1.33%** |
| % CKD | 0.13% | 0.02% | **0.50%** |
| **Biomarkers** |  |  |  |
| Glucose (mmol/L) | 5.12 (1.24) | 5.00 (0.98) |  |
| HbA1c (mmol/mol ) | 35.2 (6.78) | 35.0 (5.07) |  |
| Trig (mmol/L) | 1.75 (1.45) | 1.67 (0.98) |  |
| LDL (mmol/L) | 3.56 (0.87) | 3.58 (0.83) |  |
| HDL (mmol/L) | 1.45 (0.38) | 1.47 (0.37) |  |
| Cholesterol (mmol/L) | 5.69 (1.14) | 5.72 (1.09) |  |
| Serum Phosphate (mmol/L) | 1.16 (0.16) | 1.15 (0.16) |  |
| Serum Calcium (mmol/L) | 2.38 (0.09) | 2.37 (0.09) |  |
| Serum Creatinine (umol/L) | 72.31 (18.55) | 72.63(14.16) |  |
| Cystatin C (mg/L) | 0.91 (0.18) | 0.87 (0.13) |  |

We calculated the percentage of participants with comorbidities based on inpatient hospital records (ICD10 only; main or secondary diagnoses; Field 41270) at any time prior to the baseline visit, and also at any time prior to the imaging visit for the calcification subcohort. Compared to the entire UKBB cohort, the calcification subcohort exhibited slightly fewer comorbidities (Table 1). Within the calcification subcohort, comorbidities increased in the ∼8 years that spanned between recruitment and imaging recall. Overall, the calcification subcohort was mostly free of inpatient diagnosis with only 2.8% of participants diagnosed with diabetes and 0.5% with CKD (Table 1).

Finally, we analyzed biomarkers data from the date of initial recruitment into the UKBB cohort and found that most of the calcification subcohort had values within normal ranges, with frequencies of hyperphosphatemia (Phosphate>1.46 mmol/L) at 2.39%, hypercalcemia (Calcium>2.5 mmol/L) at 7.96%, hypercholesterolemia (LDL>190 mg/dL) at 6.15%, and hypervitaminosis D (25(OH)D3>140 ng/mL) at 0.06%.

### Automatic Quantification of Abdominal Aortic Calcification

Four human annotators quantified AAC in 1,300 randomly chosen participants by assessing lateral DEXA scans of lumbar spine (**Figure S1 and Tables S1-S2**). We used a previously established scoring method ^11^, where each segment of abdominal aorta adjacent to the L1-L4 vertebra is given a score from 0 to 6. The individual scores for each segment are summed for a maximum score of 24. Intra-annotator variability did not have a large effect on the median annotation scores (Pearsons’ correlation 0.93, **Figure S2**). We used 1,000 manually annotated images to train ML models and withheld 300 images to test the accuracy of the models (Methods).

For the ML models, we first considered a single-step regression model to directly predict AAC scores from the images; however, such an approach performed poorly due to substantial background noise within the DEXA images (**Figure S3**). We therefore developed a three step process to isolate and score the aortic region while reducing background noise (**Figures 1 and S4-S12**):

**Figure 1.**
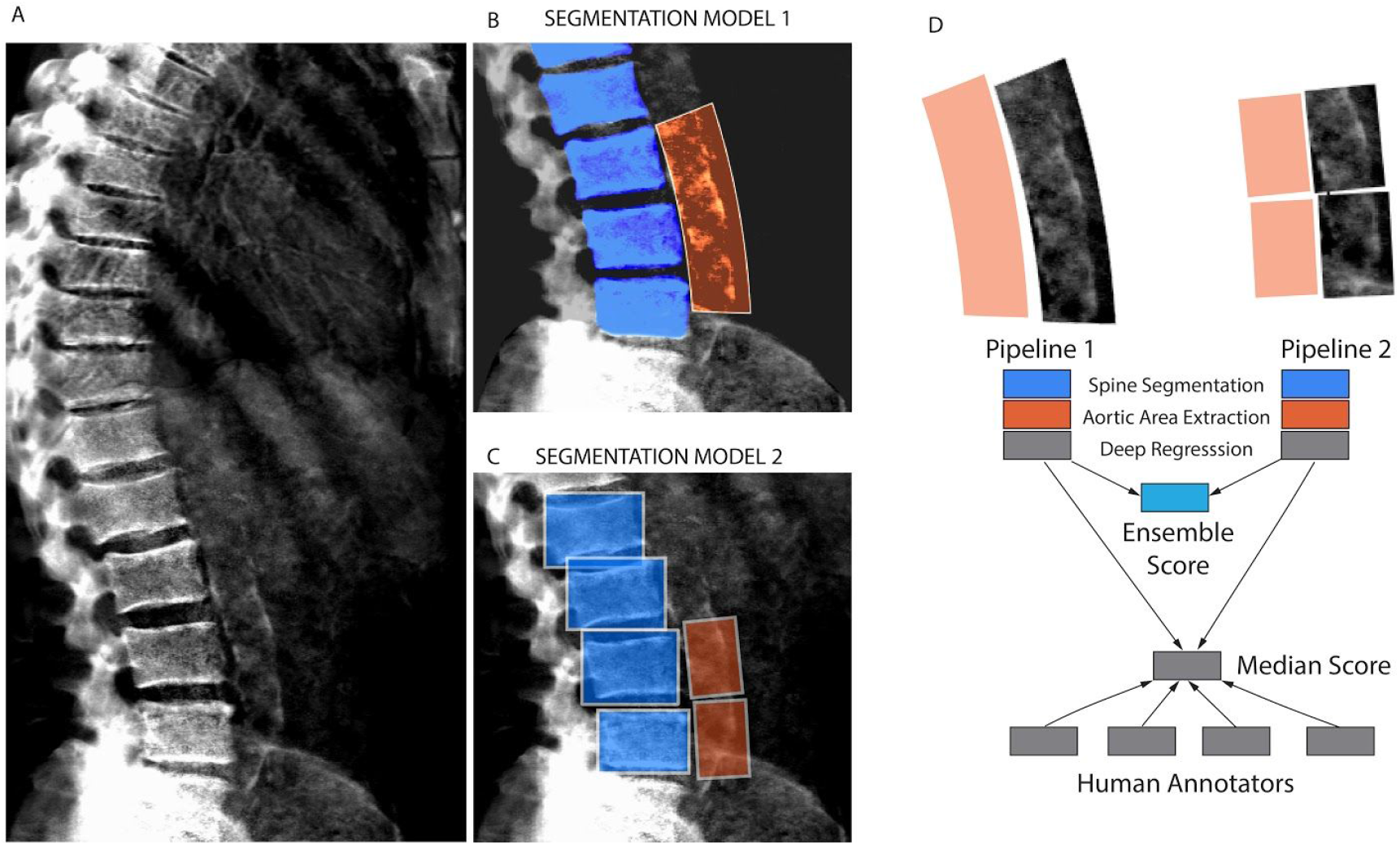
Schematic of the machine learning pipeline to quantify AAC. Overview of the two independent machine learning pipelines used to quantify AAC. (A) Representative DEXA scan (manual annotation score = 15). (B) Pipeline 1: segmentation of the lower spine using U-Net architecture (blue). (C) Pipeline 2: box segmentation of individual lumbar vertebrae (blue). Extracted aortic regions are highlighted in orange in (B) and (C). (D) AAC scores are quantified using a neural network (NN) based regression against human-derived aortic calcification scores. An ensemble AAC score, combining predictions from both pipelines, performed the best in an unseen test dataset.

1. Identify the lower lumbar vertebrae (segmentation model).
2. Extract the abdominal aortic region based on the segmented lumbar vertebrae.
3. Use the abdominal aortic region to predict AAC scores (regression model).

We developed two independent pipelines to perform each of these steps (Methods). Each pipeline achieved similar accuracy on the test dataset (Pearson’s correlation∼0.6), but an ensemble score, generated by averaging the score of each pipeline, exhibited the best performance on the test dataset (Pearson’s correlation∼0.67; **Table 2 and Figure 2**). We therefore used the ensemble method to predict AAC scores in the whole dataset (29,957 images).

**Table 2:** Accuracy of machine learning pipelines. The accuracy of the machine learning pipelines and the ensemble model were calculated using Pearsons’ and Spearman’s correlation coefficient over the test dataset. Correlation of each individual annotator with the median for the test dataset is also shown for comparison.

|  | Pearson correlation with median score | Spearman correlation with median score |
| --- | --- | --- |
| Annotator 1 | 0.94 | 0.83 |
| Annotator 2 | 0.93 | 0.88 |
| Annotator 3 | 0.85 | 0.84 |
| Annotator 4 | 0.73 | 0.57 |
| Machine Learning Models |  |  |
| Model 1 | 0.62 | 0.41 |
| Model 2 | 0.59 | 0.47 |
| Ensemble Model | 0.67 | 0.49 |

**Figure 2.**
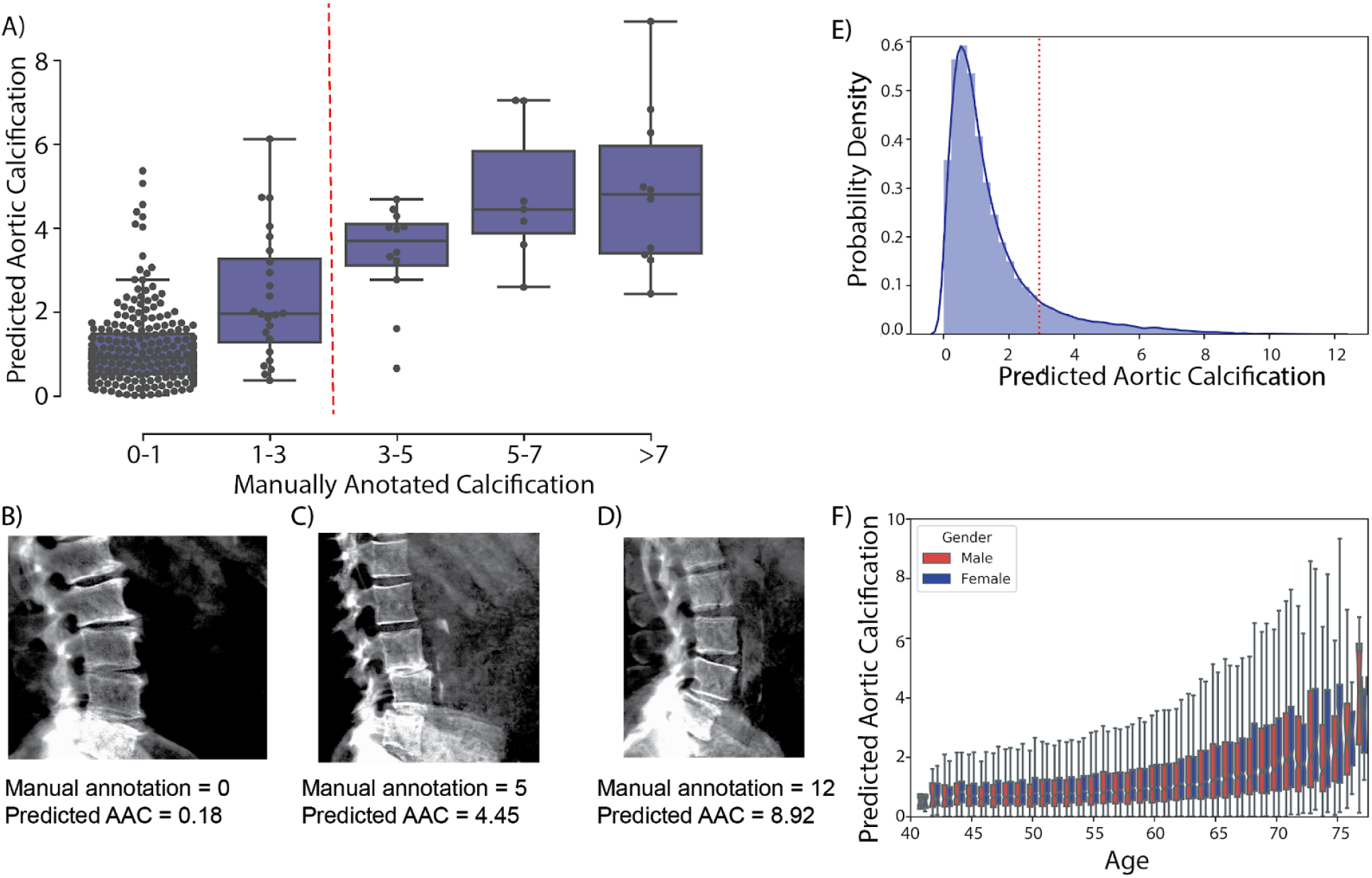
Comparison of machine learning based AAC scores to manually annotated AAC scores. (A) Correlation of manually annotated calcification scores with ensemble machine learning (ML) scores on the 300 test images. (B-D) Comparison of ML and manual scores for participants with low, medium and high levels of calcification within the test dataset. (E) Distribution of predicted AAC scores across 29,957 participants. (F) Relationship between predicted AAC scores and age of the participants. Red line in (A) and (E) indicates calcification score at 1 standard deviation (score=2.93).

The distribution of calcification scores in the cohort was highly skewed towards low values (**Figure 2E**); the majority of participants (∼88%) had no detectable or mild calcification (score< 3), with a long tail, defined as >1 standard deviation from the mean, representing ∼11.6% of the population. We confirmed these estimates in the manually annotated subset of scans, where a comparable fraction (∼10.4%) had moderate to high calcification (AAC score > 3). Both AAC severity and variability increased with age (**Figure 2F**). To our knowledge this is the first estimate of AAC prevalence from a large cohort not enriched for a specific disease. While it is known that prevalence of medial layer calcification in patients with diabetes and ESRD can be as high as 41% ^2^, we were surprised to observe such high prevalence in UKBB given that only 3.3% of the cohort has CKD and diabetes at baseline.

### Risk factors for Abdominal Aortic Calcification

We sought to identify potential risk factors for calcification with molecular biomarkers, physiological parameters, and complete blood cell counts (CBC) measures. We performed a univariate risk analysis with two different models: (i) adjusted for age and sex and (ii) adjusted for age, sex, BMI, socioeconomic factors, and smoking status (**Figures 3, S13-S14 and Table S4**).

**Figure 3:**
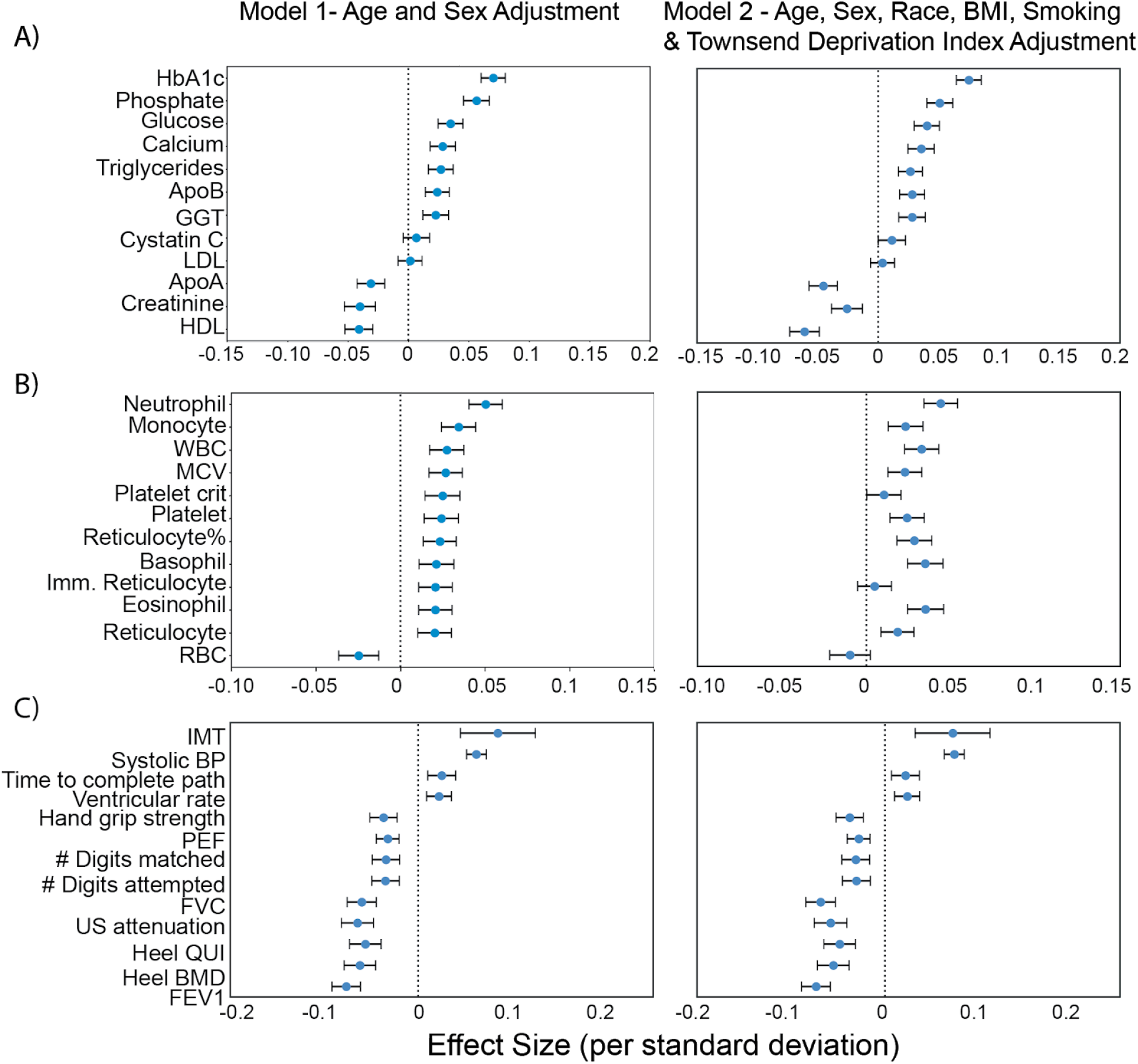
Association of AAC with (A) molecular biomarkers, (B) CBC measures, and (C) physiological parameters. Briefly, AAC is modelled as a linear function of each covariate adjusted for confounders (Methods). The blue dot represents the mean effect size per standard deviation of different covariates while error bars represent 95% confidence interval. With the exception of LDL and cystatin C, only covariates that passed multiple hypothesis testing in model 1 are shown (Bonferroni corrected p-value<1.2e-4). We include LDL and cystatin C as they are discussed in the text. Age during imaging visit was used to adjust for calcification levels. All other covariates and measurements were taken during the baseline visit.

#### Association with Biomarkers

After multiple hypothesis correction, we found that higher levels of glycated hemoglobin (HbA1c), phosphate, glucose, **calcium**, triglycerides, and gamma glutamyltransferase (GGT) in the serum are associated with increased AAC (**Figures 3 and S15 & Table S4)**. Four of the **six** associated biomarkers (HbA1c, glucose, triglycerides, and GGT) tend to be higher in diabetic patients, an observation that is consistent with diabetes being a risk factor for AAC ^12^.

Calcification is known to occur in ESRD patients due to phosphate dysregulation ^13^. However, in our study we find that the majority of participants (∼96.8%) with significant AAC are not hyperphosphatemic (serum phosphate > 1.46mM/liter, **Figure S18**). Furthermore, we observe that AAC is anticorrelated with creatinine levels (FWER p-value=3.03e-7) and is not associated with cystatin C. We estimated glomerular filtration rates (eGFR) from the cystatin C levels for all participants (FWER p-value = 1) (Methods). Across the first four quintiles of AAC, eGFR estimates were within normal kidney function ranges (eGFR ∼90, **Figure S19**). Participants in the highest quintile of AAC were older and had slightly reduced kidney function (eGFR ∼80). Given the low prevalence of CKD in this cohort (<0.5%), these results strongly suggest that calcification of abdominal arteries can occur in the absence of severe kidney disease. While coronary calcification is known to be associated with serum phosphate levels within healthy populations ^14^, as far as we know, this is the first study that implicates higher calcium levels within physiological range to also be associated with aortic calcification (Figure S18). As aging is the most predictive risk factor for calcification (Figure 2E), our analysis suggests that the risk of calcification increases with age even when serum phosphate and calcium levels are maintained at physiologic levels.

We also observe that AAC is anticorrelated with high density lipoproteins (HDLs) (FWER p-value=2.46e-9), and not associated with low density lipoproteins (LDL) (FWER p-value = 1). Presence of AAC in the absence of hypercholesterolemia and lack of association with LDL is consistent with the hypothesis that AAC is likely to be a medial layer type of calcification ^15^. We further explore the independent nature of AAC and LDL using a variety of epidemiological models to predict myocardial infarction outcomes in the final section of this study.

#### Association with Complete Blood Cell Count

Previous studies describe the frequent co-occurrence of calcification and inflammation (reviewed in ^16^). We assessed the relationship between AAC and CBC measures (**Figures 3 and S16 & Table S5)** and found AAC was positively correlated with markers of inflammation. Specifically, AAC was associated with increased cell counts of white blood cells, neutrophils, monocytes, platelets, and reticulocytes. In addition, AAC was correlated with increased mean corpuscular volume and hemoglobin concentration of red blood cells.

#### Association with Physiological Markers

We also identified several associations between AAC and physiological parameters (cutoff FWER p-value<1.2e-4; **Figures 3 and S17 & Table S6**). AAC was most strongly associated with increased intima medial thickness (IMT) of the carotid artery, which occurs due to build up of plaque and/or calcification within this artery, suggesting that calcification within the carotid artery is correlated with AAC. Similar association between AAC and IMT has been observed earlier in elderly women^17^. As expected, increased AAC was correlated with increased systolic blood pressure, increased ventricular rate ^18^, decreased bone mineral density ^19^, decreased forced vital capacity, and decreased forced expiratory volume ^20^. Overall, after taking into account age related decline, presence of aortic calcification was associated with reduced physiological function across a number of organ systems.

We carried out a replication study in the MrOS cohort to confirm biochemical and physiological associations with AAC ^21^. All major UKBB findings showed comparable associations in MrOS, including: increased level of AAC with phosphate, HbA1C, glucose, triglycerides, systolic blood pressure, as well as anticorrelation with HDL (Figure S13).

#### Genetic architecture of AAC

To identify genetic contributors to AAC, we performed rare and common variant genome wide association studies (GWASs; Methods).

For the common variant association study, we tested 9,572,557 single-nucleotide polymorphisms (SNPs) across 31,786 participants (Methods). We estimated the SNP-based heritability to be 12.4% (s.e. 1.7%; Methods). To increase our power to identify genetic contributors we combined these summary statistics with those from a previous AAC study^22^ and performed a meta-analysis spanning 5,348,079 common SNPs and 41,203 participants.

In this meta-analysis, we identified four loci associated with AAC (p-value<5×10^−8^; Figure 4A). To identify tissues and cell types relevant to AAC we partitioned the heritability of AAC genetic signals across the genome using tissue/cell type annotations derived from chromatin marks and marker genes (Methods). After multiple hypothesis correction, we found enrichment of heritability in blood vessels, vascular endothelial cells, arteries, fat-related tissues, as well as a number of other tissues (FDR<5%; Figure S20).

**Figure 4:**
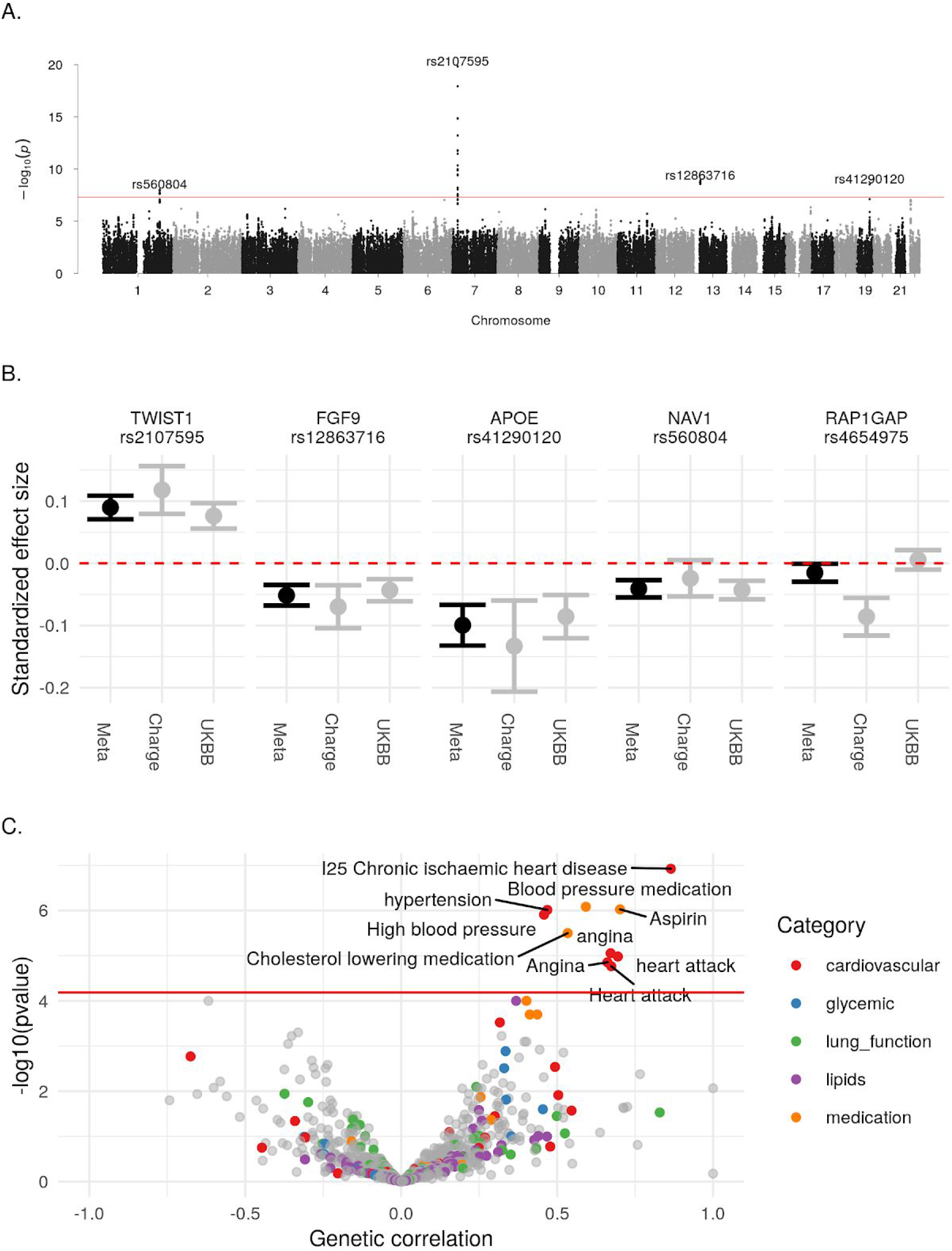
Genetic analysis of AAC. (A) Manhattan plot. Association of each SNP -log(p-values) ordered by chromosome. Genome wide significance cut-off (p-value 5×10^−8^) shown as a red line. (B) Approximate standardized effect size estimates for the UK Biobank data, CHARGE consortium data, and the metaanalysis at the four genome wide significant loci in this study, and one additional locus previously reported by CHARGE. Genetic correlation between AAC and other complex traits. X-axis: Estimated correlation. Y-axis: −log10(p-value). Horizontal line is at the Bonferroni-corrected p-value threshold.

We calculated the genetic correlation between AAC and 785 other traits (Methods). After adjustment for multiple testing, AAC was genetically correlated (p-value<6×10^−5^) with cardiovascular-related diseases and medications (Figure 4C; Supplementary Table S9). Notably, we did not find evidence of genetic correlation for other phenotypes identified as correlated with AAC in this study including inflammation markers, lung function, or lipid levels. Such results may indicate independent genetic risk factors for these phenotypes; however, lack of power may also explain these results.

#### Identification of AAC-associated loci

To better understand the biological mechanisms at each of the three identified loci, we performed in-depth analyses at each locus (Methods). First, to identify independent signals and potential causal variants, we performed conditional and credible set analysis. Second, to identify candidate effector genes, we performed colocalization analysis using associations with gene expression (i.e., expression quantitative trait loci; eQTLs) from GTEx v7. Finally, to identify other diseases and traits that likely share the same underlying causal variant, we performed genetic colocalization analysis using UKBB phenotypes that have an association (p-value<5×10-8) within 500kb of the lead SNP.

At the chromosome 7 locus, we identified two independent signals. For the primary association the lead SNP, rs2107595 (p-value=1.47×10^−20^) was the only SNP in the 95% credible set. There were 2 SNPs in the 95% credible set of the secondary association (Supplementary Table S7). rs2107595 lies in a non-coding region near *HDAC9* and *TWIST1*. This region is identified as a promoter flanking region (ENSR00000818178) active in cell types related to smooth muscle, bone, and epithelial tissue. The minor (risk) allele strongly disrupts a predicted transcription factor (TF) binding site for the E2F family of TFs and ZNF75A, increasing binding of the NOTCH signaling protein RBPJ (^23^, http://www.ensembl.org)

The rs2107595 signal colocalizes with *TWIST1* expression in aortic artery, but not with *HDAC9* expression in any tissue (Figure 5; Supplementary Table S10). We did not have any bone tissue in our expression dataset, so were unable to evaluate changes in bone. We tested if either *TWIST1* or *HDAC9* expression was correlated with calcification diagnosis from GTEx histological images of vascular tissue. We found *TWIST1* expression was associated with tibial artery calcification (beta=0.265, SE=0.0649, p-value_Bonferroni_=0.0002616), but not *HDAC9* in any tissue considered (minimum p-value_Bonferroni_=0.2488).

**Figure 5:**
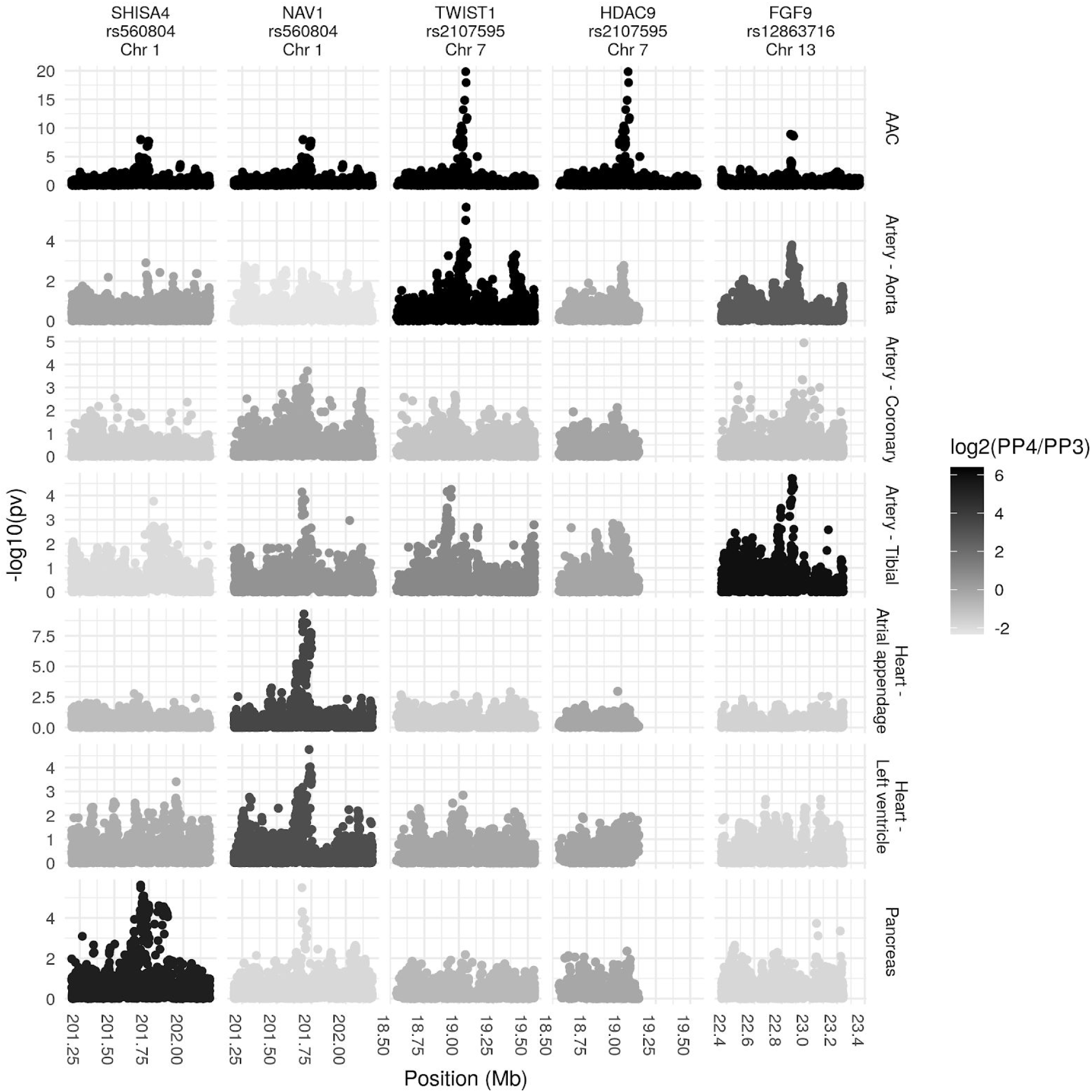
Colocalization analysis at the *NAV1/SHISA4, TWIST1/HDAC9*, and *FGF9* locus. (A) Association (y-axis) of genetic variants (x-axis) with AAC and with expression of nearby genes in relevant tissues. Only the main signal at each locus is shown.

We explored mouse aorta single cell expression data (Methods) and found *Twist1* was highly expressed in fibroblasts, vascular smooth muscle cells, and endovascular progenitor endothelial cells that also express mesenchymal marker genes (Figure S22). Finally, we also found strong evidence of colocalization between this AAC signal and several cardiovascular phenotypes: coronary artery disease (CAD), ischemic heart disease, occlusion and stenosis of precerebral arteries, and systolic blood pressure (SBP; Supplementary Table S11). We replicated the colocalization with CAD, SBP, and pulse pressure in non-UKBB datasets (Supplementary Table S12).

While we cannot draw causal conclusions of the relationship between *TWIST1*, AAC, SBP, and CAD, one hypothesis consistent with our observations is that rs2107595 increases *TWIST1* expression in vascular cells, which increases AAC, leading to increased SBP and CAD risk. Such a hypothesis is consistent with the observed increase of TWIST1 in adult human calcified aortic valves ^24,25^, the possible role of *Twist1* in promoting endothelial–mesenchymal transitions that drive vascular calcification ^26^, and theories that vascular calcification can be caused by progenitor cells (reviewed in ^27–29^).

We identified an association at the rs12863716 locus (p-value=1.2×10^−9^, 7 SNPs in 95% credible set), 600kb from the nearest coding gene *FGF9*. This signal colocalizes with *FGF9* expression in the tibial artery (Figure 5; Supplementary Table S10). Although it does not meet our threshold of significance (the sum of posterior probabilities of both traits being affected by the same locus PP3+PP4=0.73; Methods), the data is also suggestive of colocalization of expression of *FGF9* in aortic artery. rs12863716 is in LD with the lead SNP in a study of aortic aneurysm (r^2^=0.816, ^30^), and also with the lead SNP in a recent study of ascending aorta area (r^2^=0.9614, ^31^). As summary statistics were unavailable for these traits at the time of writing, we were unable to perform a formal colocalization analysis. While this signal did not colocalize with any trait in the UK Biobank (we considered traits with p-value<5×10^−8^ for colocalization), the top association in a PheWAS at this SNP was aortic aneurysm (p-value=0.00017), intracerebral hemorrhage (p-value=0.00043), and aneurysm (p-value=0.00043). *FGF9* expression has been shown to be increased in aneurysm compared to control tissue ^32^.

We found an association intronic to *NAV1* (lead SNP rs560804, p-value=9.30×10^−9^, 10 SNPs in 95% credible set). This signal colocalizes with *NAV1* expression in heart atrial appendage, with *SHISA4* in pancreas (Figure 5; Supplementary Table S10), and diastolic blood pressure (Supplementary Figure S21; Supplementary Tables 5-6). A recent study ^33^ identified *NAV1* as a candidate gene for aortic valve stenosis. The lead SNP in the aortic valve stenosis study lies in a promoter flanking region active in aorta as well as other tissues (ENSR00000382940) and is in LD (r^2^=0.62) with rs560804. The precise function of this gene and the mechanism of this connection remains unknown.

At the rs41290120 locus (p-value=2.9×10^−9^), we found no evidence for colocalization with the expression of any gene in any tissue; however, we did find strong evidence of colocalization with traits related to heart disease, red blood cell measurements, and dietary traits (Supplementary Tables 24-7). In the UK-Biobank only dataset, the lead SNP at this locus, rs1065853, is in LD (r^2^>0.99) with the APOE e2 allele, rs7412, consistent with the well-established role of APOE and lipid metabolism ^34^ The link, however, with AAC is less clear and warrants further investigation in future studies.

The CHARGE consortium also identified a possible association near *RAP1GAP*. This association does not replicate in our study (p-value=0.49) or our metaanalysis (p-value=0.04; Figure 4B; Supplementary Table 2).

We tested 18,102 genes using a generalized linear mixed model implemented in SAIGE-GENE (Zhou et al, 2020) in the AAC and exome sequencing subcohort (n=11,749 participants). Treating AAC as a binary or continuous phenotype, we detected no associations in either model (p-value_Bonferroni_<2.8×10^−6^; Methods; Supplementary Figure S23).

### Aortic Calcification is a risk factor for cardiovascular, cerebrovascular, and lung related chronic diseases

The EHR of most UKBB participants is available post imaging and these records extend to approximately 2.9 years post imaging visit (median followup time). We estimated the prognostic value of AAC using a Cox proportional hazard (CoxPH) model ^35^ on 174 disease classes. Briefly, in our analysis, only individuals within the imaging cohort that have not experienced prior history of disease are considered at risk, and time to event is calculated as time between time of imaging visit and the first occurrence of disease, thus allowing us to establish predictive value of AAC for time to disease occurrence (see Methods, and Table S13).

We considered two models: (i) adjusted for age and sex (Model1) and (ii) adjusted for age, sex, BMI, socieconomic status, race, and smoking (Model 2).

After multiple hypothesis correction (p-value_Bonferroni_<0.05), we find that AAC levels are a strong prognostic risk factor for 10 disease conditions, including a multitude of cardiovascular diseases such as: occlusion of precerebral arteries (HR∼1.5), nonrheumatic aortic valve disorders (HR∼1.35), myocardial infarction (HR∼1.36), chronic ischemic heart disease (HR∼1.3), heart failure (HR∼1.25), angina pectoris (HR∼1.2), hypertension (HR∼1.15), emphysema (HR∼1.45), respiratory diseases related to bronchitis (HR∼1.3), and emphysema (HR∼1.5; Figures 6, S24). While some conditions such as stroke, aortic valve disorders, and MI have been linked to AAC before ^36–40^, to our knowledge this is the first time AAC has been implicated in prognostic prediction of type 2 diabetes and bronchitis related breathing disorders and COPD. These observations support the hypothesis that calcification of vasculature is a commonly shared disease pathophysiology that manifests in long term negative outcomes across the entire cardiovascular system.

**Figure 6:**
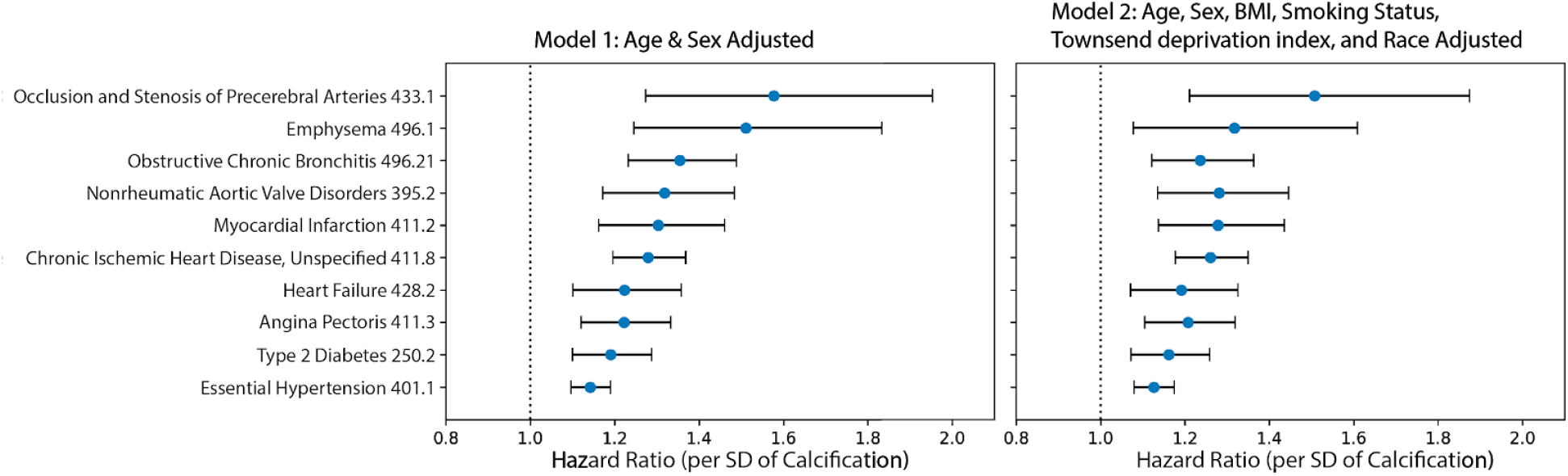
CoxPH association of AAC with prognosis of diseases. Hazard ratio for the subset of statistically significant disease associations after multiple hypothesis testing. The blue dots represent mean hazard ratio, the error bars represent 95% confidence intervals. We tested 174 diseases with more than 25 events post baseline for association with AAC using Cox PH model. Only 10 associations that passed multiple hypothesis tests in model 1 (Bonn Ferroni corrected p-value < 2.87e-4) were reported here.

Addressing the question of how calcification contributes to cardiovascular outcomes has proven to be challenging, as a number of epidemiological studies have suggested that intimal layer calcification can occur as a secondary consequence of hyperlipidemia, and that in some cases can be beneficial by stabilizing plaque formation ^41^. It is known that adding calcification scores to Framingham risk scores do improve the predictive accuracy of risk for future coronary events ^42^. Abdominal aortic calcification is suspected to mostly occur in the medial layer ^15^ but we cannot confirm this from our imaging analysis. Our analysis of biomarker data suggests that calcification of the abdominal artery is independent of LDL, however it was not clear to us if this could be due to statin confounding. Hence, we tried to elucidate whether calcification risk is independent of LDL using various models that were also corrected for statin usage.

To further investigate the hypothesis that AAC is a risk factor for myocardial infarction (MI) events independent of hypercholesterolemia, we used multivariate CoxPH models to compare additive risk and interactions between AAC and LDL. As LDL was measured during the baseline visit and AAC was measured during the imaging visit, we repeated the analysis starting from both the baseline visit (Figure 7) as well as the imaging visit (Figure S29). The imaging visit had 2.9 years of median follow up time while the baseline visit has 11.8 years of median follow up time. Herein, we describe the results from the baseline visit due to the greater statistical power. The analysis starting at the imaging visit showed similar results and can be found in a supplementary figure S29.

**Figure 7.**
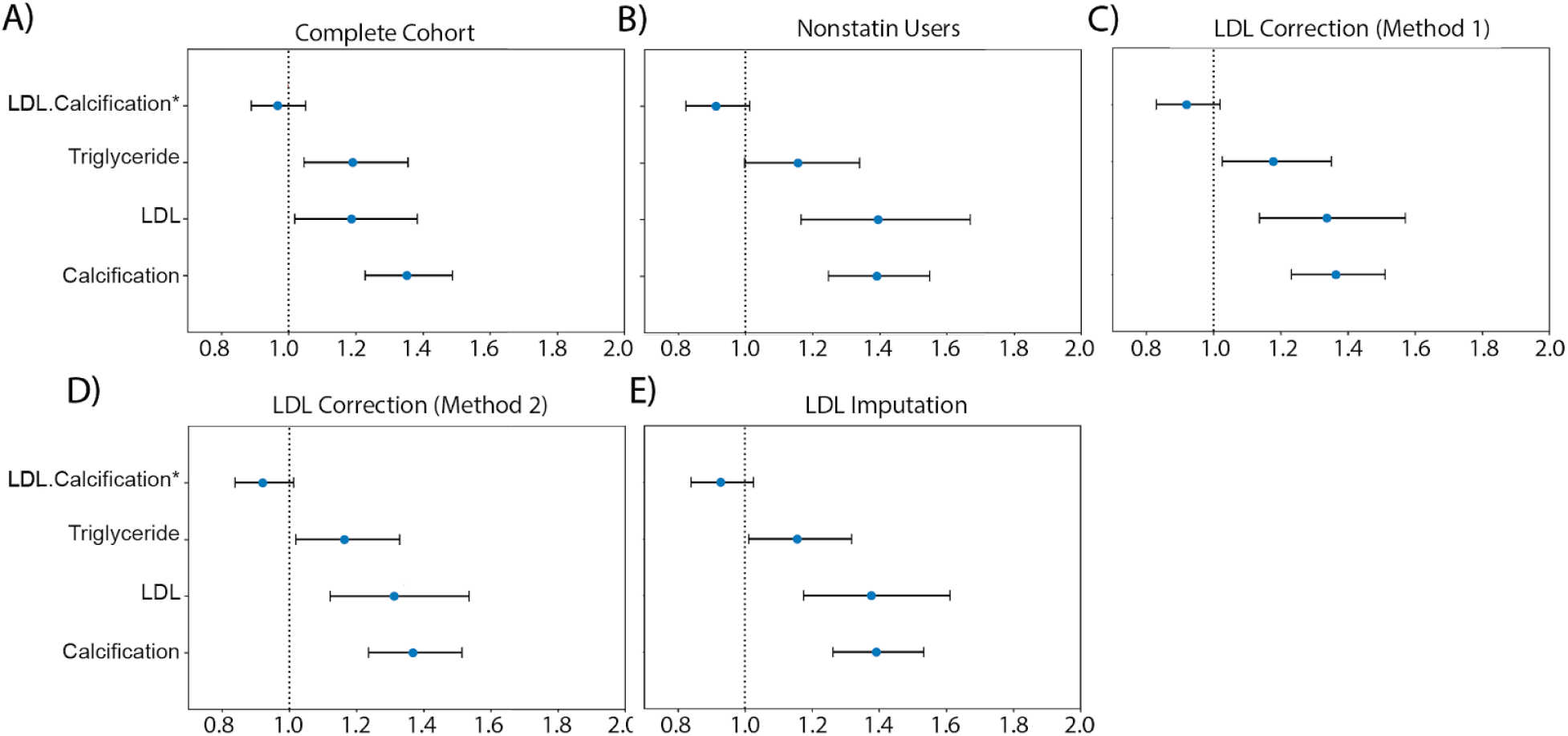
CoxPH association of AAC and LDL for acute MI events following baseline visit. We compared the hazard ratios for AAC, LDL, and triglycerides using a multivariate CoxPH model with age and sex covariates. (A) Whole cohort - no statin adjustment. (B) Subset of only non-statin users. (C) Whole cohort after adjusting for statin using method 1 - assumes that LDL levels are reduced by 1.25 mmol/L in statin users. (D) Whole cohort method 2 - assumes that LDL levels are reduced by 35% in statin users. (E) Whole cohort imputation of LDL levels based on remaining biomarkers and physiological measurements (Methods). The blue dots represent mean hazard ratio, while error bars represent 95% confidence interval. Interaction term between LDL and calcification is starred. This analysis was done with 11.8 years of median follow up time post baseline visit during which LDL was measured.

Because ∼15% of the cohort have been prescribed statins to lower LDL, this model may underestimate the contribution of LDL (HR_LDL_∼1.24, HR_AAC_∼1.4; Figure 7A). Furthermore, use of statins does not reverse cumulative exposure of elevated LDL levels and pre-existing plaque, which leads to residual risk for CVD events^43,44^. As shown in randomized clinical trials, statin usage also reduces the level of triglycerides and the CoxPH models will need to take that effect into account ^45,46^.

Therefore, to obtain an unbiased estimate for LDL and AAC MI risk, we estimated LDL risk after adjusting for statin usage using three different models (Methods; Figures S25-S27). In model 1, we adjusted the LDL and triglycerides levels for statin users by estimated average effect after increasing measured LDL and triglycerides by an amount of 1.25 mmol/L ^46^ (Figures 7C and S25). In model 2, similar model 1, LDL and triglycerides were increased by a relative increase of 54% in statin users ^46^ (Figures 7D and S25). In model 3, we imputed the untreated LDL and triglyceride levels for statin users based on blood pressure, pulse, age, sex, and non-lipid molecular biomarkers (Figures 7E and S27). In all three models, there were no adjustments to LDL and triglyceride levels for statin nonusers.

In all statin adjusted models, MI risk associated with severity of AAC (HR∼1.4 per standard deviation) was comparable to LDL risk (HR∼1.4 per standard deviation). We did not find evidence for an interaction between LDL and AAC (p-value>0.05) in any of the models, suggesting that AAC is a hypercholesterolemia-independent risk factor for MI. These results are also consistent with our genetic analysis which did not find genetic correlation between AAC and lipid biomarkers (Figure 4C; Supplementary Table Tab 8). In all statin adjusted models, we find that the adjusted LDL and triglyceride levels deconfounds statin usage from acute MI risk (Figure S28).

Despite our best effort to reduce statin bias in models 1-3, there is a potential that models are biased and do not completely remove the effects of statins usage. Therefore, we also assessed risk of AAC for MI events independent of hypercholesterolemia in the statin nonuser subgroup (Figure 7B). Statin nonusers are expected to represent a healthier subpopulation. In this group, AAC is still a significant risk factor for MI (HR=1.4), the value that is comparable to LDL (HR=1.4), with no significant interaction detected between LDL and AAC (p-value > 0.05). These results are consistent with all three statin adjusted models.

It has been reported that use of statins could increase the progression of calcification levels in coronary arteries ^47,48^. To ensure that this effect does not bias our risk estimates, we re-evaluated statin adjusted models with additional separate statin covariate. We did not find a significant contribution of statins to MI risk after LDL, AAC, triglyceride, age, and sex adjustment (p.value ∼ 0.15-0.85 Figure S28). This implies that statin usage does not confound our estimation of AAC and LDL risk levels. Finally, both statin users and non-users show similar risk profiles, results consistent with the conclusions that AAC is an LDL independent risk factor for CVD.

## Discussion

Vascular calcification is associated with cardiovascular morbidity and can arise as a complication of diabetes and chronic kidney disease ^1^. It has been poorly studied in healthy individuals due to lack of routine monitoring and lack of tools to quantitate AAC. While prevalence and heritability of AAC has been investigated previously in smaller longitudinal cohorts ^49,50^, here, we present the largest study of AAC to date, spanning 29,957 participants from a biobank approximately representative of the general UK population. To enable this study, we developed a method to automatically quantify AAC from DEXA scans. We demonstrate that these methods are broadly useful and can be applied to quantify AAC from low intensity X-ray DEXA scans. Our method is publically available (https://github.com/calico/AAC_scoring) and accurately predicts severity of calcification as judged by comparison to manual quantifications.

In our study, we observed a high prevalence of AAC (>10% of participants), even though <3.3% of the cohort is diagnosed with CKD and diabetes. We found that severity of AAC is strongly associated with elevated serum phosphate, calcium, and HbA1c, even though the levels of these biomarkers would be considered within clinically normal ranges. We found that aortic calcification was not associated with LDL, and vitamin D levels. These observations suggest that even in adults without diabetes and kidney disease, moderate elevation of serum phosphate and glucose levels are associated with higher risk of calcification and as a consequence cardiovascular outcomes. Association of normal but elevated phosphate to AAC is consistent with similar observations made in the Framingham Offspring Study ^51^ that the risk for cardiovascular disease increases with phosphate levels even in the absence of hyperphosphatemia.

The physiological rationale for ossification of abdominal aorta in a largely healthy population is not clear. Calcium/phosphate balance is tightly regulated by complex interactions between the parathyroid gland, kidney, bone, and gut. Normal response to excess calcium and phosphate is to restore balance by increased renal clearance and absorption of excess ions into bones. However, if a normal physiological response cannot be achieved, it has been suggested that smooth muscle cells transform into osteoblast-like state and hydroxyapatite is progressively deposited across various parts of cardiovascular system including heart valves, coronary arteries, aorta, abdominal aorta, tibial arteries, and kidneys ^52,53^.

In support of this “phenotypic-switch” hypothesis, our analyses suggest that vascular cells play an important role in the development of AAC. First, we found enrichment of AAC heritability in regions near marker genes for blood vessels, vascular endothelial cells, and adipose tissue. Second, we replicated a previously reported association at the *TWIST1/HDAC9* locus. Our analysis suggests this association may be mediated by an increase of *TWIST1* expression resulting in osteogenic phenotype within smooth muscle and/or vascular endothelial cells. We additionally identified three novel loci, close to *APOE, FGF9*, and *NAV1*, with evidence for genetic colocalization with *FGF9* and *NAV1* expression in cardiovascular tissue. *FGF9* and *NAV1* have been implicated in abdominal aortic aneurysm and aortic valve stenosis respectively, which may indicate the observed calcification is systemic and not isolated to abdominal aorta.

In addition to the exploration of the rs2107595 presented in this study, two recent studies investigated this locus in more depth. Nurnberg et al. ^23^ found that disrupting the rs2107595 locus in coronary artery smooth muscle alters *TWIST1* expression and that *TWIST1* expression affects proliferation and calcification phenotypes in vascular smooth muscle cells.. Malhotra et al ^22^ also identified rs2107595 as a variant associated with atherosclerotic AAC and investigated the potential role for *HDAC9* in calcification of vascular smooth muscle cells. While we cannot exclude a role for *HDAC9* in this phenotype, our analysis supports at least some involvement of *TWIST1* at this locus.

Finally, we evaluated the role of calcification as a prognostic marker of disease outcomes. We found that the severity of AAC is strongly predictive of future MI, chronic ischemic heart disease, heart failures, chronic obstructive bronchitis, type 2 diabetes, aortic valve disorders, and brain disorders. We further considered **five** epidemiological models to evaluate the independence of AAC and hypercholesterolemia. In all models, AAC was a separate additive contributor to MI outcomes, independent of LDL risk. We did not observe significant interaction in any of the models with LDL levels—supporting a hypothesis that AAC is not plaque driven but rather arises through conversion of smooth muscle cells to osteoblast-like cells and tunica media layer calcification.

Collectively, our results highlight the potential of a simple, non-invasive spine X-ray as a tool to assess the risk for a broad range of cardiovascular events, and warrants further investigation of routine AAC measurement as a prognostic indicator of cardiovascular disease. High accessibility and low X-ray exposure risk of this technique potentially opens a real opportunity to extend our methods into a real clinical setting.

The strong link between AAC and cardiovascular outcomes, that is independent of hypercholesterolemia and statin usage, suggest that development of anti-calcification therapeutics could be complementary to lipid lowering strategies. Recent clinical trials of a new class of inositol-hexaphosphate inhibitors show that it is possible to slow progression of calcification in CKD populations with negligible side effects ^54^. These studies highlight the potential for reducing cardiovascular disease burden due to calcification in a general population.

## Supporting information

Supporting Information, Genetics Tables

## Data Availability

Data:This research has been conducted using the UK Biobank Resource.

## Study Limitations

1. Measurement of LDL and AAC took place at two different visits, which limits our ability to directly compare these factors.
2. Imaging data is only available for a subset of cohort (approximately 40,000 participants).
3. The prospective disease outcome analysis with AAC was carried starting at the imaging baseline where average follow up time was 2.9 years (Figure 6).
4. Interaction between hypercholesterolemia and AAC interaction for predicting prognostic events was evaluated with both baseline and imaging visits (Figures 7 and S29), but there is on average a 9 year difference between these two visits limiting the power of imaging visits to detect significant effects due to fewer events.
5. Distinguishing between different types of calcification was not possible due to lack of histology data.

## Acknowledgements

The authors would like to thank Nick van Bruggen, Garret Fitzgerald, Amoolya Singh, Aarif Khakoo, David Kelley, Magdalena Lopez, Tian-Quan Cai, and Dan Eaton for discussions and inputs regarding the manuscript. This research has been conducted using the UK Biobank Resource application number 18448. This work was supported by Calico Life Sciences LLC.

## Abbreviations

HbA1c: glycated hemoglobin
GGT: gamma glutamyl transferase
ApoA: apolipoprotein
HDL: high density lipoprotein
LDL: low density lipoprotein
WBC: White blood cell
MCV: mean corpuscular volume
MCHC: mean corpuscular haemoglobin concentration
RBC: red blood cell, Imm.
Reticulocyte: immature reticulocyte
IMT: intima media thickness
SBP: systolic blood pressure
PEF: peak expiratory flow
FVC: forced vital capacity
QUI: quantitative ultrasound index, US- ultrasound
BMD: bone mineral density
FEV1: forced expiratory volume in 1 second

## Supporting Information

### Methods

#### Imaging Data

In 2006 through 2010, 503,000 adults (aged 40-70 years) were recruited from the general population in the United Kingdom into a prospective cohort study (Sudlow et al. 2015). The UK Biobank aims to scan 100,000 participants by the end of 2023 with various imaging modalities (Petersen et al. 2013); the current analysis includes 31,494 participants for whom lumbar spine DEXA imaging scans (Lunar iDXA densitometer; GE Healthcare, Chicago, Illinois) were collected for body composition and bone mineral density assessments (April 2014-September 2019). At the imaging assessment visit, information was collected on a range of demographic and lifestyle factors, including ethnicity, education, occupation, alcohol consumption, smoking status, socioeconomic status, and physical activity. Various measurements were also taken, including height, weight, waist and hip circumferences, and blood pressure. Systolic blood pressure and diastolic blood pressure were measured twice after the participant had been at rest for at least 5 minutes in the seated position by using a digital sphygmomanometer (Omron 705 IT; OMRON Healthcare Europe B.V., Hoofddorp, Netherlands) with a suitably sized cuff; the average of the two systolic blood pressure measures was used in all analyses. Further details about the procedural characteristics for the imaging data have been published online^1^.

#### Annotation of Abdominal Aortic Calcification

The baseline abdominal aortic calcification for 1,000 randomly chosen participants was manually quantified by four annotators using the 24-point scheme described by Kauppila and coworkers (Kauppila et al. 1997). Calcification scores of the anterior and posterior wall of the abdominal aorta over 1st lumbar spine to 4th lumbar spine (L1 to L4) were recorded. Calcification score of each section ranges from 0 to 3. Score 0 represents without any calcification; score 1 represents calcification length less than 1/3 of vertebra; score 2 represents the calcification length spanned from 1/3 to 2/3 of vertebra; score 3 represents calcification length greater than 2/3 of vertebra. AAC score is the sum of calcification score from L1 to L4 with a maximum of 24. AAC score 0 represents no abdominal aortic calcification in lateral lumbar x-ray and AAC score 24 represents a most severe degree of abdominal aortic calcification in lateral lumbar X-ray scan. The median annotation values for 1000 images of these images were used to train and test the machine learning models (read Machine Learning section in Methods). The median annotation scores were stable even on repeat assessments by all annotators (**Figure S2**). The median annotation scores were also assessed for accuracy by comparing associations with biomarkers from the same participants in the MrOS cohort (Cawthon et al. 2016). We also created a test dataset of manually annotated 300 randomly chosen images using the procedure above to assess the accuracy of the machine learning pipelines.

#### Machine Learning

We developed an ensemble machine learning approach that combined two pipelines to score aortic calcification levels based only on lumbar spine DEXA scans. Both models developed as part of this work estimate AAC scores via the following 3 steps (details in the methods sections):

1. Segmentation of the lower spine region.
2. Localization of the Aortic region using a spine-curve fitting method.
3. Regression on the localized region to predict the calcification levels from the aortic region.

The DEXA images analyzed in this manuscript were downloaded in three batches. The DEXA images were not all identical in size, but were similar in scale: 444.7(mean) +/- 27.4(SD) pixels wide and 940.0(mean) +/- 8.3(SD) pixels high. The second batch of images that we analyzed had a different distribution of dimensions: 755.1(mean) +/- 65.3(SD) pixels wide and 1665.9(mean) +/- 114.7(SD) pixels high. The third batch of images was similar to the second: 781.5(mean) +/- 43.9(SD) pixels wide and 1654.8(mean) +/- 104.5(SD) pixels high. All images in the second and third batches were rescaled to 55% of their original size along each axis.

##### Pipeline 1

###### Detection of the Vertebrae Using Segmentation

Previous studies (Han et al. 2018; Fan et al. 2019; Lessmann et al. 2019) have successfully used a fully convolutional neural network to segment out the vertebrae and determine the spinal curvature. The segmentation model in Pipeline 1 employs a similar approach using a U-Net to achieve semantic segmentation of the Pelvis and the lower spine.

In order to localize the region of the DEXA scan corresponding to the Aorta, correct anatomic locations of the Pelvis (**P**) and 4 vertebrae (**L3-L5, S1**) are needed. The network to achieve the necessary segmentation is based on a U-Net architecture. The network architecture consists of a contracting path (left side) and an expansive path (right side) as shown in **Figure S4**. The contracting path follows the typical architecture of a convolutional network. It consists of the repeated application of two 3×3 convolutions (unpadded convolutions), each followed by a batch normalization layer, a rectified linear unit (ReLU) and a 2×2 max pooling operation with stride 2 for downsampling. Each downsampling step doubles the number of feature channels. Every step in the expansive path consists of an upsampling of the feature map followed by a 2×2 convolution (“up-convolution”) that halves the number of feature channels, a concatenation with the corresponding feature map from the contracting path (residual connection), and two 3×3 convolutions, each followed by a ReLU. At the final layer a 1×1 convolution is used to map each 16-component feature vector to one of 3 classes - spine, pelvis or background.

The UKBB dataset has a total of 31,494 lumbar spine DEXA scans of patients available to score. Of those, 200 images were randomly chosen for manual annotations of **P** and **L3-L5, S1**. The png images of the scans were loaded on an open sourced annotation tool, QuPath (Bankhead et al. 2017), and the relevant anatomical locations were marked using the polygon tool. The user annotations were converted to binary masks for the 3 classes - Pelvis, vertebrae and background. Of the 200 images, 175 were used for training and cross-validation while the rest were reserved as an unseen test set to quantify the segmentation performance.

Simple data augmentations in the form of crop and zoom, left-right flips and slight rotations (upto +/- 10 degrees) were used to augment the images in the training dataset. The network was trained with the multi-class cross-entropy loss using an Adam optimizer. The evaluation metric is the mean IoU for each of the 3 classes which is defined as (**Figure S5**):

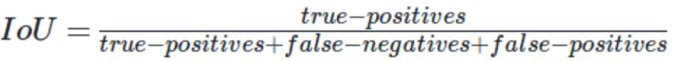

###### Aortic Region Extraction

Once the segmentations of **P** and **L3-L5, S1** are completed, the aortic region is localized and extracted via the following steps:

1. Run the segmentations through post-processing steps to:
  a. Eliminate false positives for **P** (determined through co-localization with **L3-L5, S1**).
  b. Fix broken or missing vertebrae using median height and width estimates for the different vertebrae
2. Determine the centroids of each of **L3, L4, L5, S1** and **P**. Fit a spline curve to pass through each of these points to determine the spinal curvature
3. Along the spinal curvature, move to the right by a fixed offset **A**_**off**_ and extract a rectangular region whose width is **A**_**width**_ and whose height is determined by the vertical distance between the centroids of **L3** and **P**. Crop and/or pad the images to be of size 196×196 pixels.

As illustrated in **Figure S6**, at the output of this step, the Aortic regions will be localized and extracted from the original CT images. These regions are then passed on to the Regression module to be converted into a calcification score.

###### Regression

The final step in translating the DEXA images to a calcification score is to run a regression model that maps the extracted aortic regions to a score. The regression model consists of a backbone feature extractor followed by a fully connected layer and an output layer with a single node that outputs the calcification score. The backbone network is typically borrowed from the classification task on the ImageNet database. This paper explores 3 different backbone feature extractors:

1. InceptionV3 (Szegedy et al. 2015)
2. ResNet50 (He et al. 2015)
3. Custom convolutional neural network consisting of 4 layers of convolution, batch normalization and a ReLu activation layer each.

Of the 1300 user annotated calcification levels, 1000 images were used for training and cross-validation while the other 300 were held out as an unseen test set. As illustrated in **Figure S7**, the ground truth data is highly skewed towards the lower calcification scores creating a high degree of data imbalance. In order to introduce some degree of balance, the dataset was augmented in a stratified manner. Augmentation routines include horizontal and vertical flips, random rotations, zoom and crop, random brightness changes, random contrast changes as well as random hue changes. During agile augmentation, images with higher scores were augmented with different combinations of the routines to produce as many as 32 variations of every image while images with lower calcification scores were augmented only once or not at all.

Since the training set was quite small, both InceptionV3 and ResNet backbones overfit very readily to the dataset and performed quite poorly on the validation sets. The best results on the validation set were obtained when using the custom convolutional network as the backbone feature extractor. All results presented in this paper were, therefore, generated using this custom backbone network shown in the figure.

During training, the regression model used a weighted mean squared error metric wherein errors in the higher calcification scores were weighted higher than those in the lower scores, once again with the intention of offsetting the high degree of skew in the ground truth score distributions. The network is trained with an Adam optimizer and the evaluation metric is the correlation between the predicted scores and the median user scores. The final correlation score between the manually annotated AAC scores and the predicted AAC scores on an unseen validation set is shown in **Figure S7**.

##### Pipeline 2

The second pipeline we developed in this manuscript seeks to score the degree of abdominal aortic calcification through analysis of a DEXA image. The analysis occurs in three steps:

1. identification of the spine, and use of the labelled vertebrae to identify & extract the small region within the image that will contain the aorta;
2. classification of the aortic sub-image using three models (one classifying calcified-vs-not using a low threshold for calcification, another classifying the same using a high threshold for calcification, and a third classifying the extent of background noise in the image into low, medium, or high categories);
3. use of the scores assigned to each category by the above classification models to assign a calcification score using an ML regression model.

###### Detection of the Vertebrae Using Segmentation

The clinical scoring system for AAC involves the assignment of points based on the intensity with which the walls of the abdominal aorta can be visualized in an x-ray image adjacent to the L1-L4 lumbar vertebrae. Here, we attempted to isolate the relevant portion of the DEXA image for such evaluation by first mapping the spine and then capturing a smaller image adjacent to the L3 & L4 vertebrae. For our study, DEXA images were available in bulk. But they are not ideally suited to the detection of AAC: high levels of background intensity would often appear adjacent to the L1 & L2 vertebrae, often due to visualization of the rib cage. We therefore sought to isolate just the part of the image adjacent to vertebrae L3 & L4. That strategy is illustrated by the examples in Figure S8, with the lumbar vertebrae labelled in green and the desired aortic region circled in orange. This goal was achieved through two sequential processes as described below:

The foundation of this AAC scoring system is the annotation of the spine and its constituent vertebrae in the DEXA image. That annotation defines the landmarks that are used to isolate the portion of the image that contains the aorta, a task that cannot be performed independently of these landmarks because the aorta is only clearly visible in these images when it is extensively calcified.

This section describes the steps taken to annotate as many individual vertebrae as possible while minimizing the false-positive annotation of other parts of the image as vertebrae. This task was initially approached with a simple object-detection model. In order to correct mistakes made during the initial segmentation step, we chose to implement a series of analyses & models to detect & correct errors. The end product is an analysis pipeline that is summarized in **Figure S8**.

Initially, vertebrae are boxed using an object-detection model that is applied with a very low score threshold (“draw vertebra boxes”; few false negatives, many false positives). Incorrect annotations that are far from the spine are easily detected and removed using the unusual vertebra-to-vertebra angles that are produced (“trim off-axis vertebrae”). Each remaining vertebra is then scrutinized by a classification model that is trained using high-quality versus low-quality annotations that were produced by the initial object-detection model, i.e. a model to specifically address the first model’s errors (“remove low-quality vertebrae”). Then, missing vertebrae (either missed initially or removed because they were poorly defined) are detected and filled in, using the context of the nearby annotations to increase sensitivity and specificity versus the initial object-detection model. In the first case, skipped vertebrae are detected (“classify vertebra pairs”) and filled in by a model appropriate to the number of consecutive vertebrae that were missed (“fill-in one” or “fill-in multi”). In the second case, missing vertebrae at the bottom of the spine are detected (“have we hit bottom?”) and filled in (“extend down by one”). The performance of each substep is summarized in **Figure S9**.

###### Aortic Region Extraction

After identifying all the vertebrae in the spine, the region of the scan adjacent to the L3 and L4 lumbar vertebrae was extracted: this was the region of the scan with the abdominal aorta.

Given an outline of the spine, it is straightforward to determine which vertebra is which given a single reference point. In these DEXA images, the border between the L4 and L5 vertebrae was often coincident with the top edge of the spine. An object-detection model was trained by transfer learning from the ssd_mobilnet_v1 model. The best box returned by the model was used, irrespective of its score. Performance was measured using IoU (0.893 and 0.818 for training and test sets respectively).

The aortic images were created by stacking rectangular images gathered from adjacent to the L3 and L4 vertebrae, using the following four-step process:

a. definition of a vector pointing towards the aorta for each vertebra;
b. definition offset distances and image widths using the dimensions of the lumbar bounding boxes and aortic vectors;
c. definition of sub-image heights & extraction of the aortic sub-images adjacent to L3 and L4; and
d. rightening & stacking the L3- and L4-adjacent sub-images into a single output image.

For each input image (full DEXA scans), the end result of the analyses described in the links above was a smaller image depicting the regions adjacent to the L3 vertebra (above the white line) and the L4 vertebra (below the white line). The inability to identify both of those vertebrae resulted in no output aortic image.

###### Regression

The overall distribution of AAC values is highly skewed towards little or no aortic calcification. This property was observable in the training set, with most rater-generated scores at zero (Figure 1). We developed two models, each described below, in order to address the problem of sparsity of training data for high-calcification scores. The first model focuses on the lower end of the distribution, and was trained to distinguish between images with zero-value versus non-zero AAC scores. The second focuses on the higher end of the score distribution, where I used a more-efficient but less-precise-than-scoring method to enrich a larger test data set for high-calcification images.

a. **Model 1:** For the training data, four raters scored calcification as an integer, and the median value was taken of those four scores. For this low-threshold AAC model, any image with a rater-median score of 0.5 or greater was considered “calcified”. That approximately split the training set (264 “calcified” images, 332 “non-calcified” images). The ROC curves to measure the accuracy of this model is shown in **Figure S10** while Cohen’s kappa for these models were 0.58 and 0.33 for the training and test sets respectively.
b. **Model 2:** For the purpose of helping to develop higher-threshold calcification models, we designated a set of 5000 “sandbox” images that were non-overlapping with the validation and training sets, and were therefore of potential use to the model developers as training data, but for which they would not provide manual ratings. We used those data by iteratively applying a low-threshold model to those images, sorting out the “calcified” images, then manually enriching those images for yet-higher calcification values by selecting the apparently-more-calcified images from image pairs until we had sufficient data to train another model. We repeated this process until we had arrived at a training set with 170 “calcified” and 4654 “non-calcified” images, at a threshold that we estimated to be at approximately score=5. For evaluation, the original training and test sets suffered from sparsity of high-scoring data, making our evaluation of their performance sensitive to statistical noise. The ROC curves and Cohen’s kappa values for a) the actual, “sandbox”-enriched training set (yellow - 0.71); b) the original training set, with a score threshold of 5 (green - 0.73), and c) the original test set, again with a score threshold of 5 (blue - 0.53) (**Figure S10**).

For the final output value of the model (AAC score), we built and trained a small regression model to input the probability scores from the classification models above and output an AAC estimate. For the binary classification tasks, we used just one of the two outputs. The structure of the model is shown in **Figure S11**. This model has 52 trainable parameters. We experimented with many model structures and multiple attempts at training the model described above, evaluating performance using the “test” set. The statistics for the final model are shown for an unseen validation set in **Figure S12**.

##### Ensemble Model

Due to the paucity of labeled data and to avoid overfitting by using the same labeled data for training both pipelines and the ensemble model, we used an unsupervised ensemble method to combine the scores from both pipelines. In particular, we took the mean calcification levels predicted by both pipelines as the ensemble prediction. We tested the accuracy of the ensemble method on 300 test images that were not used for training either model and the accuracy of the ensemble model was higher than the accuracy of either pipeline (Table 2). Hence, we used the ensemble method to quantify calcification for all participants within the cohort and these scores were used for all downstream analysis in this article.

#### Associations with Biomarkers/Physiological Markers

We used linear regression to examine the association of aortic calcification with various biomarkers and physiological markers. For these models, univariate associations of aortic calcification with each marker were evaluated with two different models. In model 1, all univariate associations were performed after adjusting for age and sex, while in model 2, we also adjusted for BMI, Townsend deprivation index, smoking status, and race in addition to age and sex. We chose these factors based on their associations with traditional cardiovascular outcomes and/or their association with aortic calcification. All p-values were calculated using the two-tailed t-statistic of the estimated association. The associations were considered to be significant after multiple hypothesis testing (i.e., Bonferroni Correction with p-value < 1.2e-4). These estimations were calculated using the statsmodels package v.0.9.0 in python (Seabold and Perktold 2010).

#### Rare variant association study

Exome sequencing variant calls from the raw FE variant calling pipeline (Regier et al, 2020) were downloaded from the UK Biobank website (http://biobank.ctsu.ox.ac.uk/crystal/field.cgi?id=23160). QC was performed in PLINK v.1.90 using the following criteria: removal of samples with discordant sex (no self-reported sex provided, ambiguous genetic sex, or discordance between genetic and self-reported sex), sample-level missingness <0.02, European genetic ancestry as defined by the UK Biobank (Bycroft et al, 2018). Variant annotation was performed using VEP v100, filtered for rare (MAF<0.01) putative loss-of-function variants including predicted high-confidence loss-of-function variants, predicted using the LOFTEE plugin (Karczewski et al, 2019). 11,749 samples and 18,102 genes were analyzed in a generalized linear mixed model as implemented in SAIGE-GENE (Zhou et al, 2020). For the dichotomous study, AAC was binarized into 1,274 cases, defined as raw AAC score >=3, and 10,475 controls, defined as raw AAC score < 3. Rank-normalized AAC and binarized AAC were both regressed on gene carrier status, adjusted for genetic sex, age, and PC1:10 as fixed effects and genetic relatedness as a random effects term. A kinship matrix was built in SAIGE off of a filtered set of genotyped variants (r2<0.2, MAF>=0.05, HWE p>1e-10 in European population). A filtering step of at least 10 loss-of-function carriers per gene was applied, resulting in 8,794 genes.

#### Common variant genome wide association study

We used the UKBB imputed genotypes (Bycroft et al. 2018), excluding SNPs with a minor allele frequency <1% and poor imputation quality (info value <0.9). We removed particpants who were not Caucasian, exhibited sex chromosome aneuploidy, heterozygosity outliers, or genotype call rate outliers. In total, we considered 9,572,557 SNPs and 31,786 individuals (Supplementary Table 1) for genetic analysis.

To conduct the genetic association study, we used BOLT-LMM (Loh et al. 2015) and standardized machine-learned AAC, including genotype SNP chip (Illumina vs Affimetrix), sex, age, age^2^, and recruitment center as fixed effect covariates and genetic relatedness derived from genotyped SNPs as a random effect to control for population structure and relatedness. We verified that the test statistics showed no inflation compared to the expectation using the genomic control lambda coefficient (1.0257) and the intercept (1.0248, s.d. 0.0069) of linkage disequilibrium (LD) score regression (LDSC) (B. K. Bulik-Sullivan et al. 2015).

#### Metaanalysis

We combined the summary statistics from our UK Biobank study with those from the CHARGE consortium (Malhotra et al. 2019) using a fixed effects metaanalysis as implemented in the software METAL (Willer, Li, and Abecasis 2010). As effect sizes were not comparable across studies, and not available for the CHARGE dataset, we used a sample-size weighting scheme to combine the studies. The total sample size was 41,203. For ease of interpretation, we included only SNPs present in all the CHARGE sub-studies and the UK Biobank analysis for downstream analysis. This resulted in a total of 5,348,079 SNPs.

We estimate the effect size and standard error on a standardized scale using the formulae 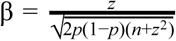 and 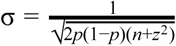

where p is the minor allele frequency, n is the combined sample size, and z the z-statistic (Zhu et al. 2016).

#### Genetic architecture of AAC

##### Identification of distinct association signals

We performed approximate conditional analysis using GCTA (Yang et al. 2012), considering all variants that passed quality control measures and were within 500kb of the locus index variant. As a reference panel for LD calculations, we used genotypes from 5,000 UKBB participants (Bycroft et al. 2018) that were randomly selected after filtering for unrelated, Caucasian participants. We excluded the major histocompatibility complex (MHC) region due to the complexity of LD structure at this locus (GRCh37::6:28,477,797-33,448,354; see https://www.ncbi.nlm.nih.gov/grc/human/regions/MHC). For each locus, we considered variants with locus-wide evidence of association (p-value_joint_<10^−6^) to be conditionally independent.

##### Construction of genetic credible sets

For each distinct signal, we calculated credible sets (Wellcome Trust Case Control Consortium et al. 2012) with 95% probability of containing at least one variant with a true effect size not equal to zero. We first computed the natural log approximate Bayes factor (Wakefield 2007), Λ_j_, for the j th variant within the fine-mapping region:

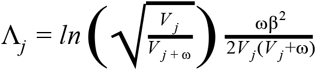

where β_j_ and V_j_ denote the estimated allelic effect (log odds ratio for case control studies) and corresponding variance. The parameter ω denotes the prior variance in allelic effects and is set to (0.2)^2^ for case control studies (Wakefield 2007) and (0.15σ)^2^ for quantitative traits (Giambartolomei et al. 2014), where σ is the standard deviation of the phenotype estimated using the variance of coefficients (Var(β_j_)), minor allele frequency (f_j_), and sample size (n_j_; see the sdY.est function from the coloc R package (Giambartolomei et al. 2014)):

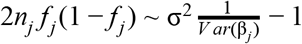

Here, σ^2^ is the coefficient of the regression, estimating σ such that 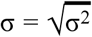.

We calculated the posterior probability, π_j_, that the j th variant is driving the association, given l variants in the region, by:

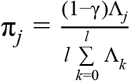

where γ denotes the prior probability for no association at this locus and k indexes the variants in the region (with k=0 allowing for the possibility of no association in the region). We set γ=0.05 to control for the expected false discovery rate of 5%, since we used a threshold of p-value_marginal_<5×10^−8^ to identify loci for fine-mapping. We note that setting γ=0 generates credible sets as proposed by The Wellcome Trust Case Control Consortium et al. (Wellcome Trust Case Control Consortium et al. 2012) and is suitable when one is very confident of the identified loci (e.g., replicated across many studies).

To construct the credible set, we (i) sorted variants by increasing Bayes factors (natural log scale), (ii) included variants until the cumulative sum of the posterior probabilities was >=1−c, where c corresponds to the credible set cutoff of 0.95.

##### Heritability estimates

We estimated the heritability of each trait using the restricted maximum likelihood method (Yang et al. 2010), as implemented in BOLT-LMM with the --reml option.

##### Genetic correlation of AAC with other phenotypes

We estimated the genetic correlation of AAC with phenotypes using an LDSC-based method (B. Bulik-Sullivan et al. 2015), as implemented in the LD Hub web resource (Zheng et al. 2017). For this analysis and all other analyses using LDSC, we followed the recommendation of the developers and (i) removed variants with imputation quality (info) <0.9 because the info value is correlated with the LD score and could introduce bias, (ii) excluded the major histocompatibility complex (MHC) region due to the complexity of LD structure at this locus (GRCh37::6:28,477,797-33,448,354; see https://www.ncbi.nlm.nih.gov/grc/human/regions/MHC), and (ii) restricted to HapMap3 SNPs (International HapMap 3 Consortium et al. 2010). We restricted our analysis to the 836 traits analysed in European populations. We successfully estimated genetic correlation for 754 traits. We additionally analysed 31 blood and urine biomarkers using data from the UK Biobank, to give a total of 785 traits.

##### Partitioning of AAC heritability

We used LDSC to partition the heritability of AAC according to functional categories (Finucane et al. 2015) as well as tissue/cell type specific annotations (Finucane et al. 2018).

For functional categories, we used the baseline v2.2 annotations provided by the developers (https://data.broadinstitute.org/alkesgroup/LDSCORE). Following Finucane et al. (Finucane et al. 2018), we calculated tissue specific enrichments using a model that includes the full baseline annotations as well as annotations derived from (i) chromatin information from the NIH Roadmap Epigenomics (Roadmap Epigenomics Consortium et al. 2015) and ENCODE (ENCODE Project Consortium 2012) projects (including the EN-TEx data subset of ENCODE which matches many of the GTEx tissues, but from different donors), (ii) tissue/cell type specific expression markers from GTEx v6p (GTEx Consortium et al. 2017) and other datasets (Fehrmann et al. 2015; Pers et al. 2015), and (iii) immune cell type expression markers from the ImmGen Consortium (Heng, Painter, and Immunological Genome Project Consortium 2008). For each annotation set, we controlled for the number of tests using the Storey and Tibshirani procedure (Storey and Tibshirani 2003). As noted by (Finucane et al. 2015), although heritability is non-negative, the unbiased LDSC heritability estimate is unbounded; thus, it is possible for the estimated heritability, and therefore enrichment, to be negative (e.g., if the true heritability is near zero and/or the sampling error is large due to small sample sizes).

In order to enable visualization, we grouped tissue/cell types into systems (e.g., “blood or immune”, “central nervous system”). These groupings and labels were the same as those used in Finucane et al. (Finucane et al. 2018), except for (i) the immune expression labels which we extended to include “stem cells” and “stromal cells” according to the ImmGen cell type classifications (http://www.immgen.org) and (ii) the “pancreas” label which we replaced with an “endocrine” label composed of the following tissue/cell types: adrenal gland, ovary, pancreas, pituitary, prostate, testis, thyroid, adrenal cortex, adrenal gland, endocrine gland, gonads, granulosa cells, islets of langerhans, and glucagon sensing cells.

#### Genetic colocalization of AAC with other phenotypes

We performed colocalization analysis using the coloc R package (Giambartolomei et al. 2014) using default priors and all variants within 500kb of the index variant. As performed by Guo et al. (Guo et al. 2015), we considered two genetic signals to have strong evidence of colocalization if PP3+PP4≥0.99 and PP4/PP3≥5 and suggestive evidence of colocalization if PP3+PP4≥0.8 and PP4/PP3≥3. For gene expression colocalizations, we used summary statistics from GTEx v7 (GTEx Consortium et al. 2017). For disease and quantitative trait colocalizations, we used UKBB summary statistics of PheCodes (Zhou et al. 2018), normalized quantitative traits (http://www.nealelab.is/blog/2017/7/19/rapid-gwas-of-thousands-of-phenotypes-for-337000-samples-in-the-uk-biobank). For analysis we selected UKBB phenotypes where the minimum p-value within the +-500kb region around the locus tag SNP was <5×10^−8^.

#### Follow up analysis at the rs2107595 locus

To assess the correlation between *TWIST1* and *HDAC9* expression and calcification visible on histological imaging, we downloaded the sample annotations from https://storage.googleapis.com/gtex_analysis_v7/annotations/GTEx_v7_Annotations_SampleAttributesDS.txt. We defined a participant’s vascular tissue as calcified if the annotation ‘calcification’ appeared in the ‘Pathology categories’ field. We downloaded expression data from GTEx v7 (dbGaP Accession phs000424.v7.p2). For each arterial tissue (coronary, aorta, or tibial) and gene (*TWIST1* or *HDAC9*), we used a logistic regression model, adjusted for age and sex, to assess the association between expression levels and calcificaiton. We used the Bonferroni procedure to correct for multiple models by multiplying each p-value by 6. We explored *Twist1* expression patterns in single cell expression data from mouse aorta (Kalluri et al. 2019) (Figure S22).

In addition to the colocalizations described in the main text, we tested for colocalization with lipid-related quantitative traits directly measured in UKBB, even though no genetic association (p-value<5×10^−8^) was found at this locus, due to the importance of lipids for coronary artery disease (CAD)-related phenotypes. We found no evidence of colocalization (Supplementary Tables 4, 5, 6, 7), suggesting that the genetic effect at the rs2107595 locus on AAC and CAD-related phenotypes is not directly related to lipid biology. We also repeated the colocalization analysis for CAD and blood pressure traits using genetic studies that did not include UKBB participants (Nikpay et al. 2015), (Hoffmann et al. 2017). We found similarly strong evidence of colocalization for both traits (Supplementary Table 7).

Finally, given the strong colocalization of AAC with CAD and SBP signals as well as the substantially larger sample sizes of these traits (n_effective_ for CAD=76,054 and SBP=340,159), we performed fine-mapping at this locus using associations with CAD and SBP in UKBB. Two SNPs, rs2107595 and rs57301765, that are in strong LD with each other (1000GENOMES:phase_3:GBR r^2^>0.99) constituted the 95% credible sets for both traits.

#### Prognostic Analyses of Aortic Calcification

##### Association of different diseases with AAC

We used Cox proportional hazards models to examine the association of aortic calcification with various diseases. The 10th edition of the international classification of diseases (ICD10) diagnosis codes and date of initial diagnoses were extracted from electronic health records of all participants. The ICD10 codes are hierarchically organized and were created for insurance billing purposes. ThePheWAS codes attempt to group the different ICD10 codes into medically meaningful groups (Denny et al. 2010). We converted the ICD10 codes at level 2 hierarchy into PheWAS codes before associating with predicted aortic calcification. Any participant diagnosed with the PheWAS code prior to baseline was removed from analysis before calculating associations with aortic calcification. The CVD events for composite outcomes in Figure S29 were defined similarly to the composite outcome for each PheWAS code.

For these models, univariate associations of aortic calcification with each marker were evaluated with two different models. In model 1, all univariate associations were performed after adjusting for age and sex, while in model 2, we also adjusted for BMI, Townsend deprivation index, and race in addition to age and sex. We chose these factors based on their associations with traditional cardiovascular outcomes and/or their association with aortic calcification. All p-values were calculated using the two-tailed t-statistic of the estimated association. These estimations were calculated using the statsmodels package v.0.9.0 in python (Seabold and Perktold 2010).

##### Comparison of risk from Aortic Calcification and LDL

We used Cox proportional hazards models (Cox 2018) to compare the risk for acute myocardial infarction from aortic calcification and LDL with the following model:

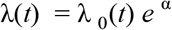

where:

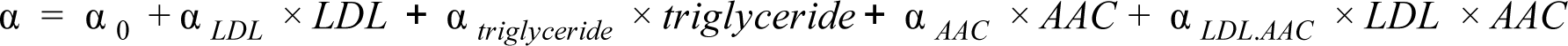

where α _0_ represents the baseline risk, α _*LDL*_ represents the risk due to increase of LDL, α _*AAC*_ represents the risk due to increase in aortic calcification, α _*triglyceride*_ represents the risk due to increase in triglycerides, and α _*LDL.AAC*_ was used to measure the risk due to the interaction between LDL and triglycerides. The LDL and triglyceride levels were logged and standardized while aortic calcification levels were standardized before measuring the risk for acute MI events.

Acute MI events were defined as the first occurence of I21, I22, I23, I24.1 or I25.2 from the ICD10 codes in the electronic health records similar to (Millett, Peters, and Woodward 2018). Any participant diagnosed with acute MI events prior to baseline was removed from analysis before comparing risks from aortic calcification and LDL. We created four different models to correct the risk for statin usage that reduces LDL without reducing systolic blood pressure completely (Figure S25 and S26) in addition to a naive model in which no statin correction was performed. The first model was a naive model built for statin nonusers and the estimates for LDL risk are confounded by survivorship bias due to the nonrandomness of statin usage. In model 2, we adjusted the LDL and triglyceride levels for statin users by adding 1.25 mmol/L to the LDL levels for statin users while no adjustment was performed for non-statin users (Nissen et al. 2005) (Figure S25). In model 3, we adjusted the LDL and triglyceride levels for statin users by dividing the measured LDL for statin users by 0.65 while no adjustment was performed for non-statin users (Nissen et al. 2005) (Figure S25). Finally, in model 4, we imputed the LDL levels for statin users based on systolic blood pressure, diastolic blood pressure, pulse, age, sex, and measured blood biomarker (albumin, alkaline phosphatase, alanine aminotransferase, aspartate aminotransferase, bilirubin, urea, calcium, creatinine, cystatin C, gamma glutamyltransferase, glucose, glycated haemoglobin (HbA1c), insulin growth factor, phosphate, rheumatoid factor. Testosterone, sex hormone binding globin, total protein, urate, and vitamin D) levels in serum. Imputation was performed using multiple imputation by chained equations (MICE) (Azur et al. 2011). In the MICE procedure a series of regression models are run whereby each variable with missing data is modeled conditional upon the other variables in the data. To evaluate the sensitivity of the results to the imputed values, we performed the risk of LDL and aortic calcification with ten different sets of LDL and triglyceride imputations and each MICE imputation was performed with 50 iterations to relax from initial estimates. The estimated risk scores were not sensitive to the imputation set as shown in Figure S27.

### Supporting Information Text

#### Manual Annotation of Abdominal Aortic Calcification

Four annotators quantified the abdominal aortic calcification by assessing the digitized baseline lateral DEXA scans of lumbar spine using a visual semiquantitative method (Kauppila et al. 1997) for 1300 randomly chosen participants. In this method, the severity of calcific deposits in the anterior and posterior walls of the abdominal aorta adjacent to each of the first four lumbar vertebrae (L1-L4) were assessed individually, using the midpoint of the intervertebral space above and below the vertebrae as boundaries. Severity scores for each of these eight segments (0–3) were added to yield an AAC score (0-24).

We utilized 1000 images for training and testing both machine learning pipelines while 300 images were kept aside for estimating accuracy on a validation dataset. The inter-annotator variability was measured over the training data initially (**Table S1**). In general, the inter-annotator variability was lowest for highly calcified and low/noncalcified individuals while annotations were most variable for participants with intermediate levels of calcification (**Figure S1**). To assess the effect of intra-annotator variability of the median annotation scores, we chose 136 images with the highest inter-annotator variability within the training dataset (i.e., the images on which there was the largest disagreement within different annotators) and all four annotators re-annotated these images. While the inter-annotator variability remained within patients with high calcification scores, the reannotated median calcification score showed a Pearson correlation of 0.93 with the original calcification scores indicating that the median calcifiction score was a pretty reliable indicator of calcification within the cohort (**Figure S2**). The calcification scores for the 300 images in the validation dataset were also highly correlated across annotators as well as with the median calcification score (**Tables S2**). The manually annotated median calcification scores also have similar trends of correlations with biomarkers in the MrOS cohort with expert annotated calcification scores (Szulc et al. 2014) (**Table S3**).

#### Automation of Estimating Abdominal Aortic Calcification

The UKBB has collected whole body DEXA scans for 31,494 of the participants to estimate their bone mineral density. As the previous sections explain, the calcification of the Aorta is visible in these scans and can be quantified based on the scoring methodology illustrated in (de Bie et al. 2017) (**Figure S8**). Coronary arteries are visible in DEXA scans only when calcified.

The first approach this paper took towards scoring these images was to train a deep regression model that mapped the complete lumbar spine CT images on to user annotated AAC scores. The model used a ResNet50 architecture (He et al. 2015) as backbone and had a fully connected layer followed by a linear output layer to score the calcification levels in the images. However, due to the highly variable background noise in the images coupled with the high degree of noise in the user annotated scores, the model performed quite poorly as can be seen in **Figure S3** below even on a limited range of scores.

The automatic scoring methodology adopted in both models of this paper, therefore, relies on a segmentation methodology of large structures that would always be visible in these scans such as the Pelvis and the Spine in order to first localize the region where the Aorta can be seen. Once localized, any calcification that is seen in the Aortic region is quantified and mapped to a score via a regression model. Both pipelines developed as part of this work estimates AAC via 3 steps (details in methods).

The main goal of the computer vision and machine learning exercise was to find a way to automate and scale the scoring of aortic calcifications to the entire UKBB dataset in the absence of a large volume of ground truth user annotations. The segmentation, region extraction and regression architectures were all chosen with this constraint in mind. Since the annotated dataset was quite small, we explored 2 separate machine learning models to automate the scoring of these images. Eventually, the scores from these 2 models were averaged to create an ensemble score. As expected, the ensemble score had a higher correlation score to the median annotated scores than either of the 2 models separately.

With this ensemble method, we have managed to score a total of 29,957 DEXA images with only 200 segmentation annotations and 1300 calcification score annotations. Given that the UKBB imaging modality is highly consistent, the models given out in the github page can very well be used to infer the scores on future DEXA scans as they become available.

### SI Figures

**Figure S1:**
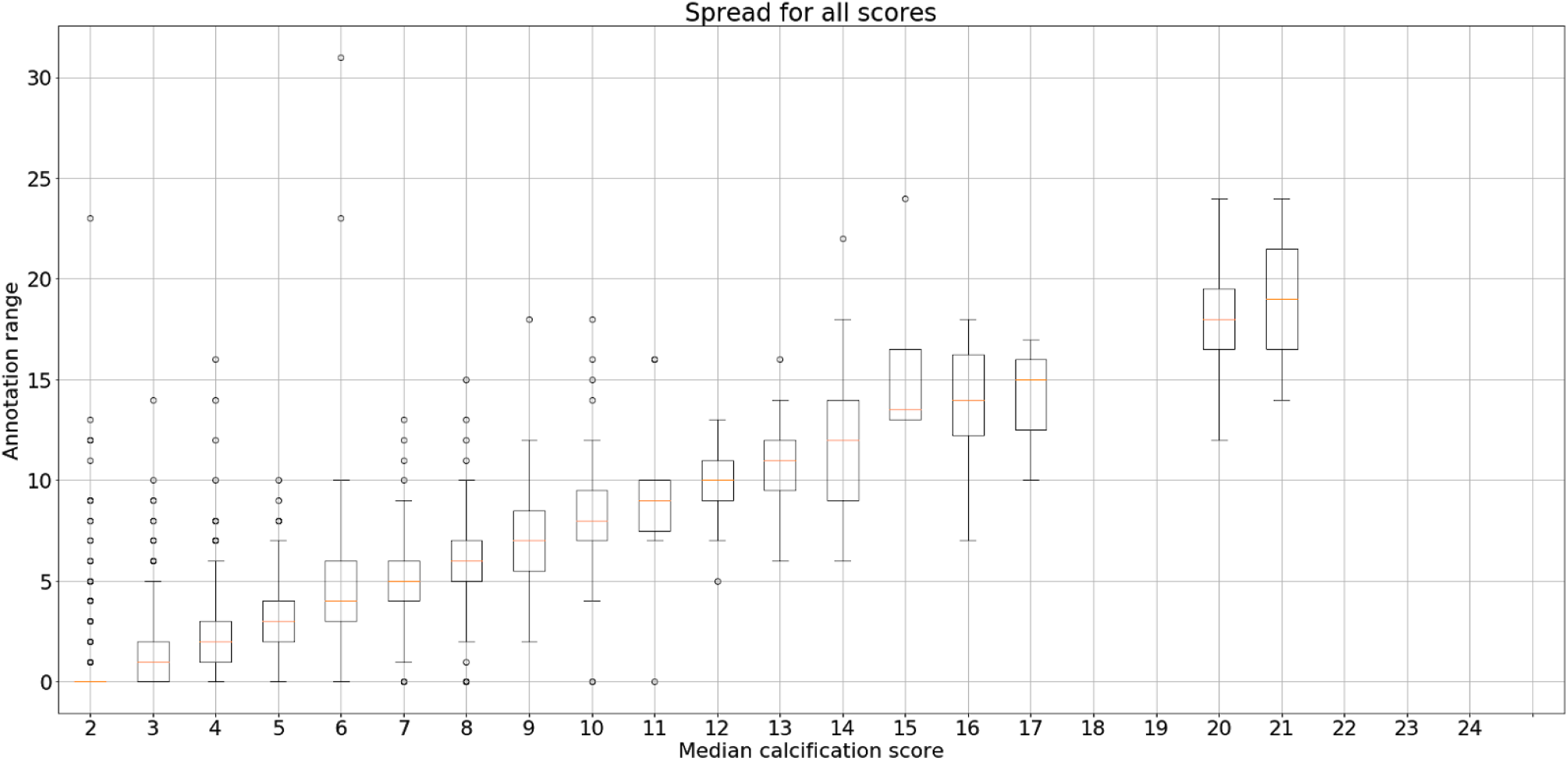
The variability in calcification across annotators increases for medium level calcified participants.

**Figure S2:**
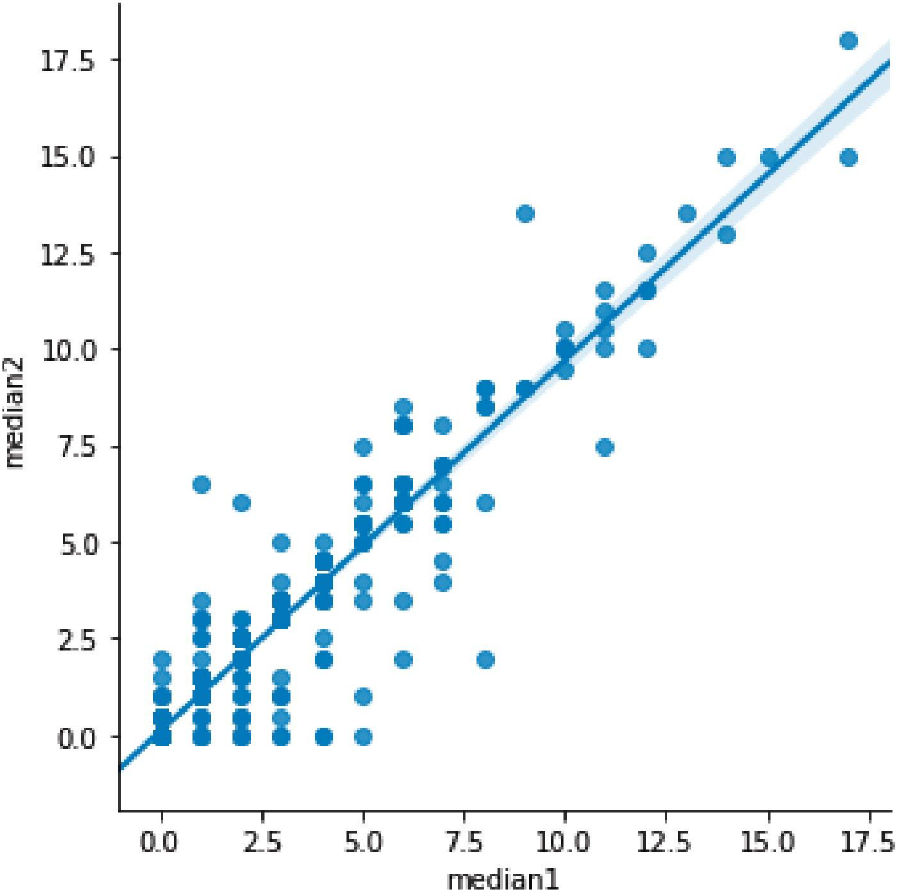
The median scores for 136 manually annotated scans from two rounds of annotation are highly correlated.

**Figure S3:**
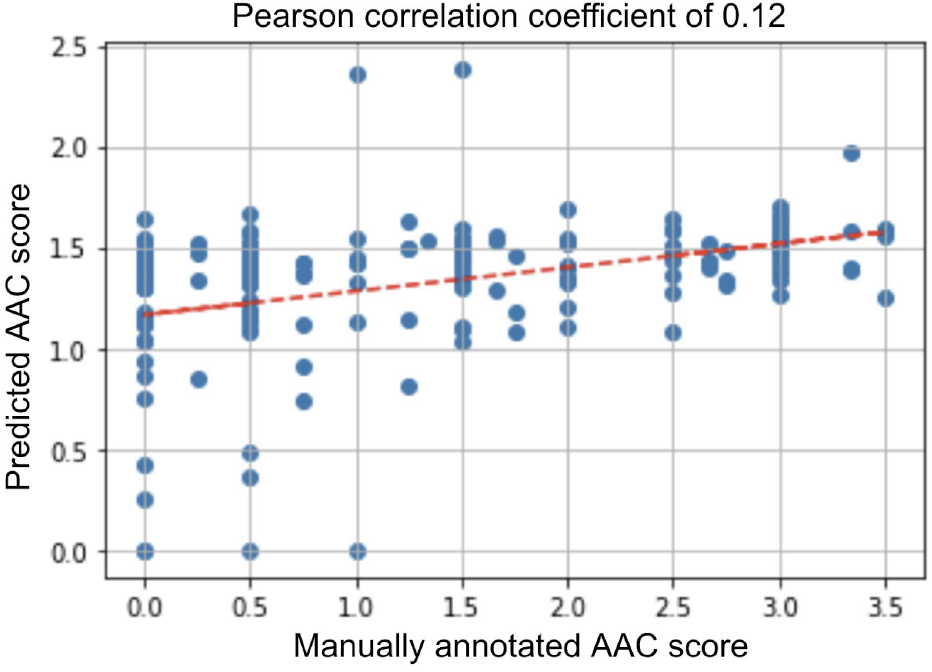
Conventional deep learning regression approaches with ResNet50 architecture performed poorly on a test set while quantifying aortic calcification from the complete lumbar spine DEXA scans.

**Figure S4:**
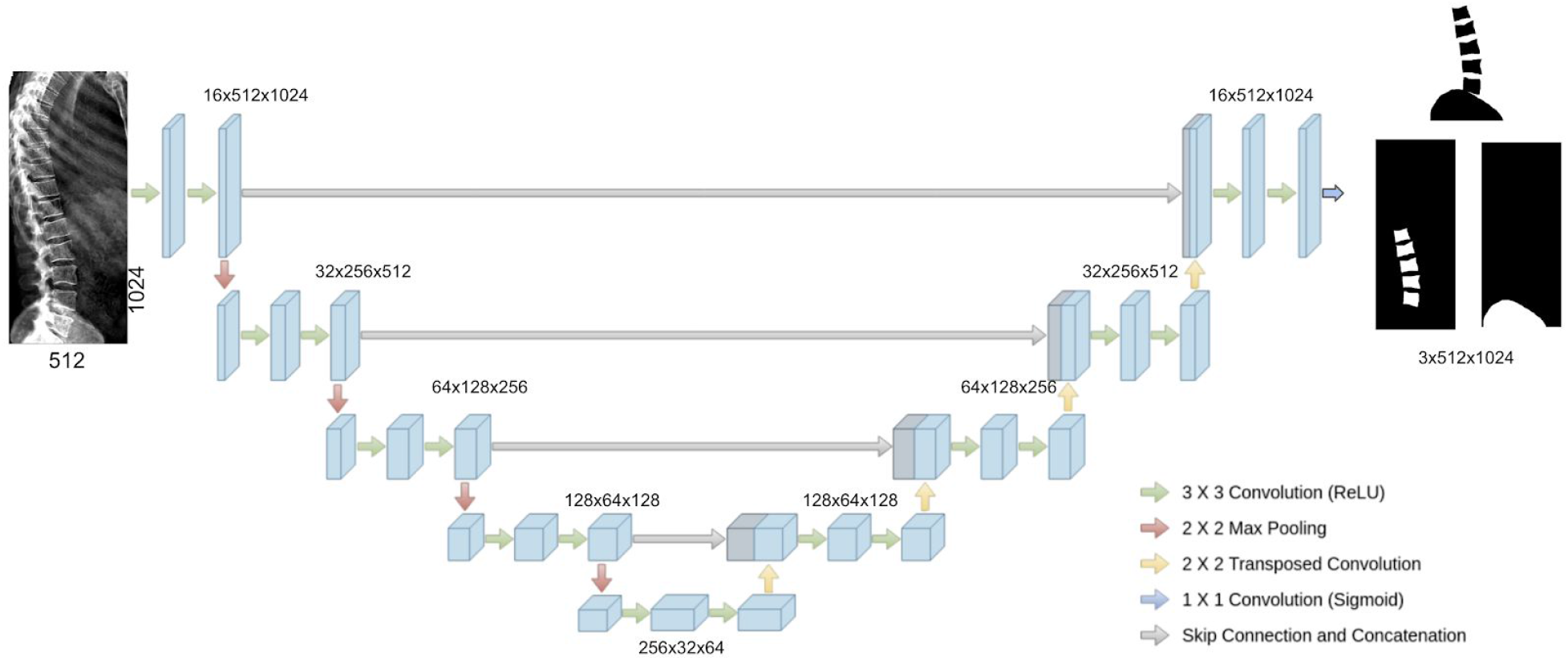
U-net architecture used for segmenting relevant spine and pelvis regions in pipeline 1. The input DEXA image is shown on the left and the binary masks for the 3 output classes - background, lower spine and pelvis - are shown on the right.

**Figure S5:**
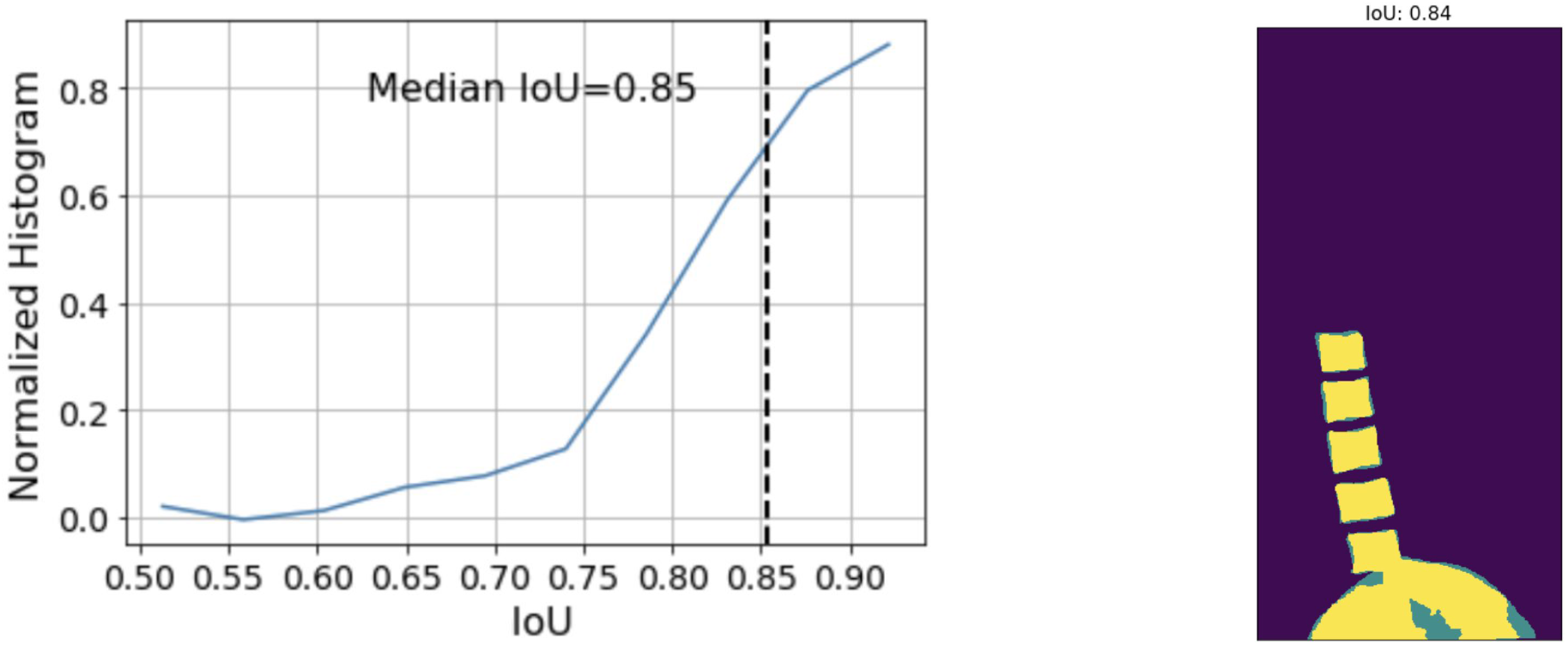
**(a)** shows the histogram of IoU values obtained across the test set along with the median value. **(b)** shows an overlay of the ground truth segmentation mask and the predicted segmentation mask for a typical example image whose IoU is close to the median value. The yellow regions show the regions of overlap while the blue regions show gaps in the prediction mask.

**Figure S6:**
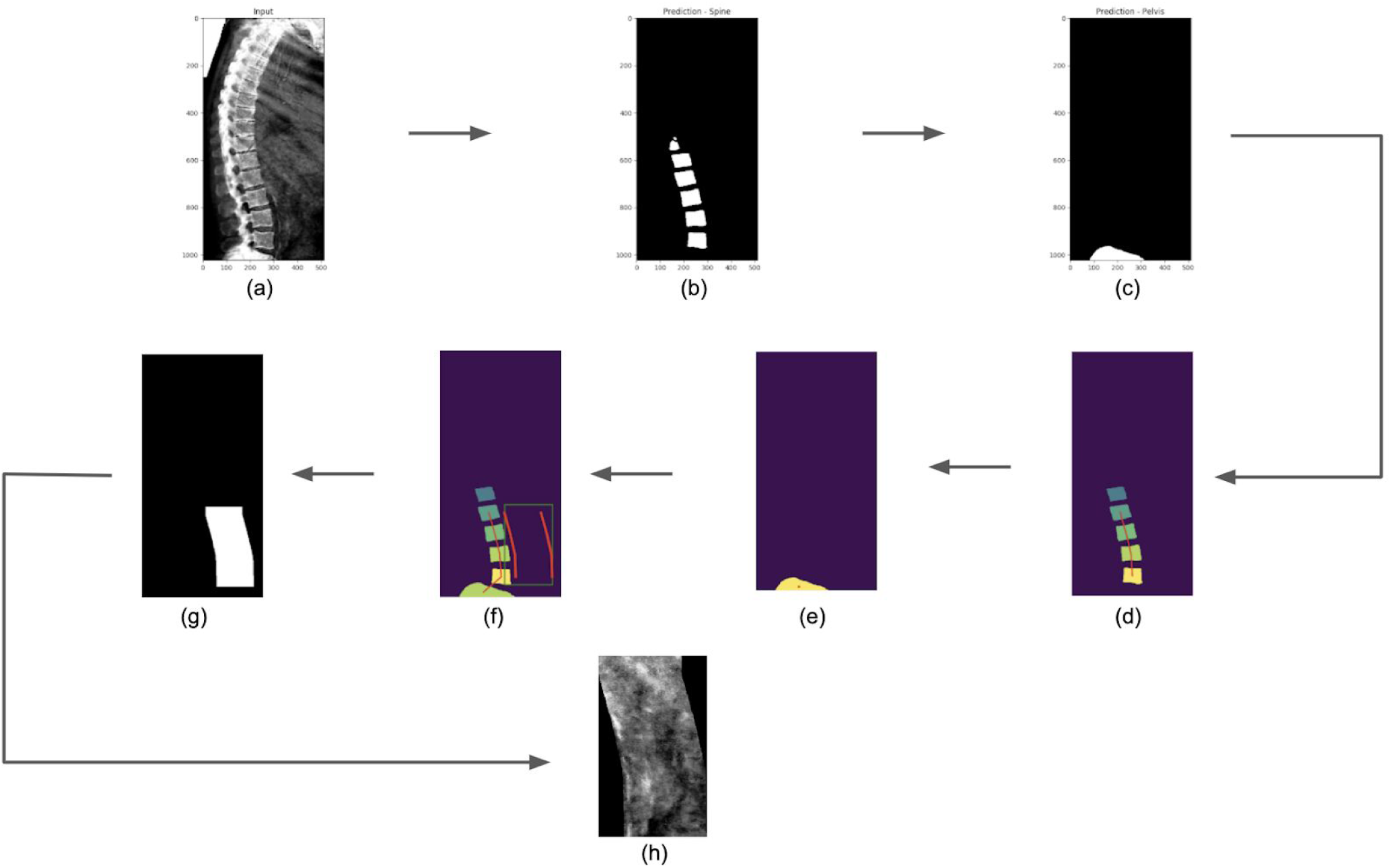
Schematic representation of the steps to extract abdominal aortic region from lumbar spine DEXA scans. (a) shows the input image. (b)-(c) show the predicted segmentation masks of the lower spine and the pelvis. (d)-(f) show the predicted centroids of the different vertebrae (connected), predicted centroid of the pelvis and the overall spinal curvature together with the estimated aortic region to the right of the spine. (g) shows the binary mask of the predicted aortic region. (h) shows the aortic region extracted from the original input image. This is then fed into a regression model to score the calcification level.

**Figure S7:**
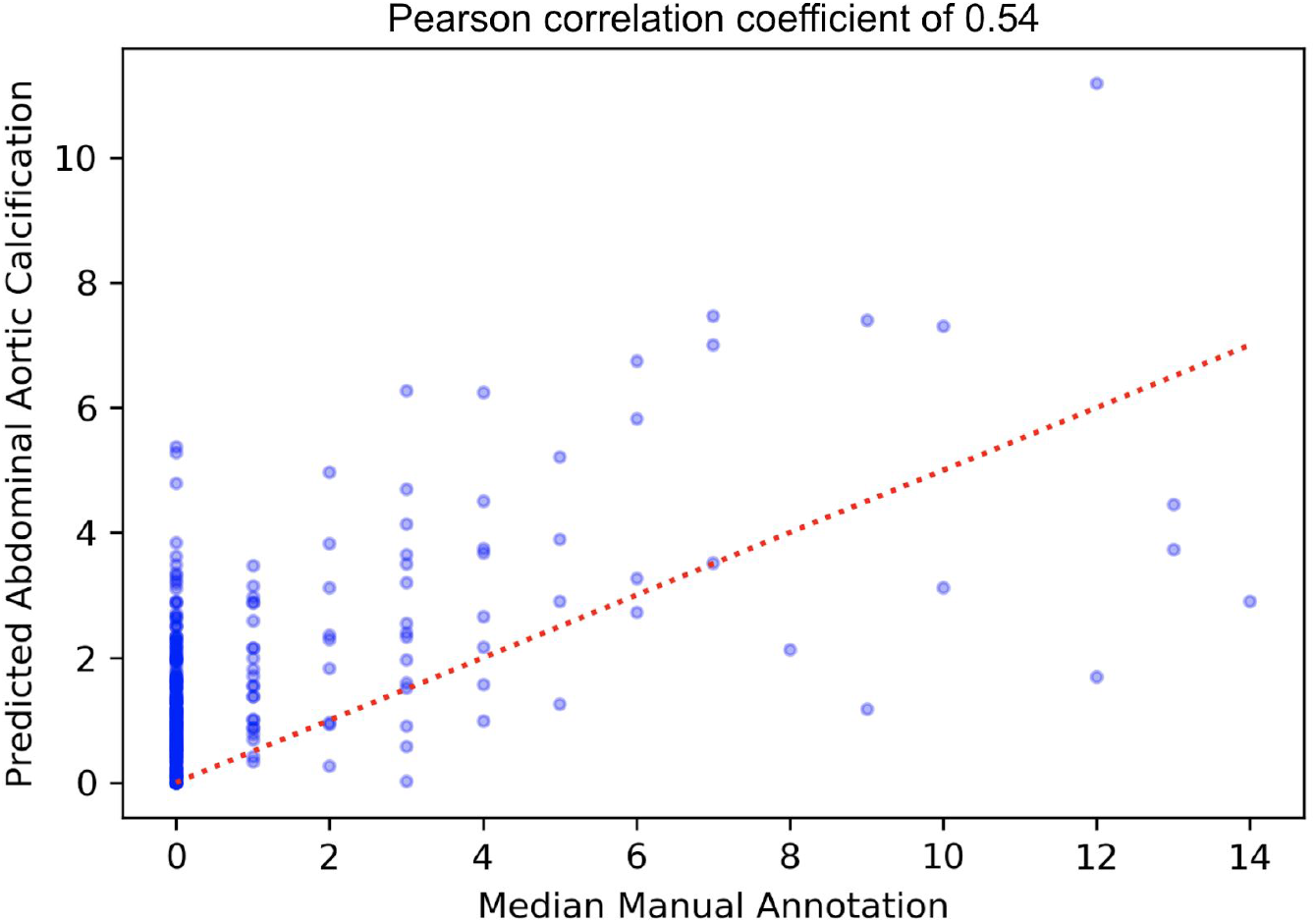
The predicted AAC from pipeline 1 as compared to manually annotated calcification scores for the validation dataset.

**Figure S8:**
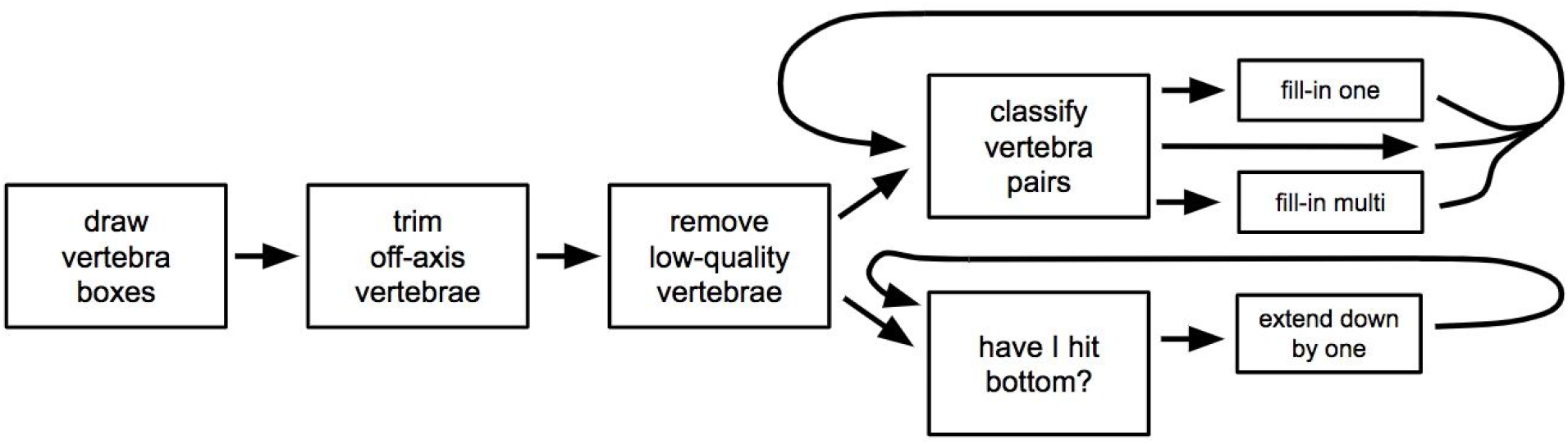
Segmentation steps within machine learning pipeline 2 for predicting AAC from DEXA scans.

**Figure S9:**
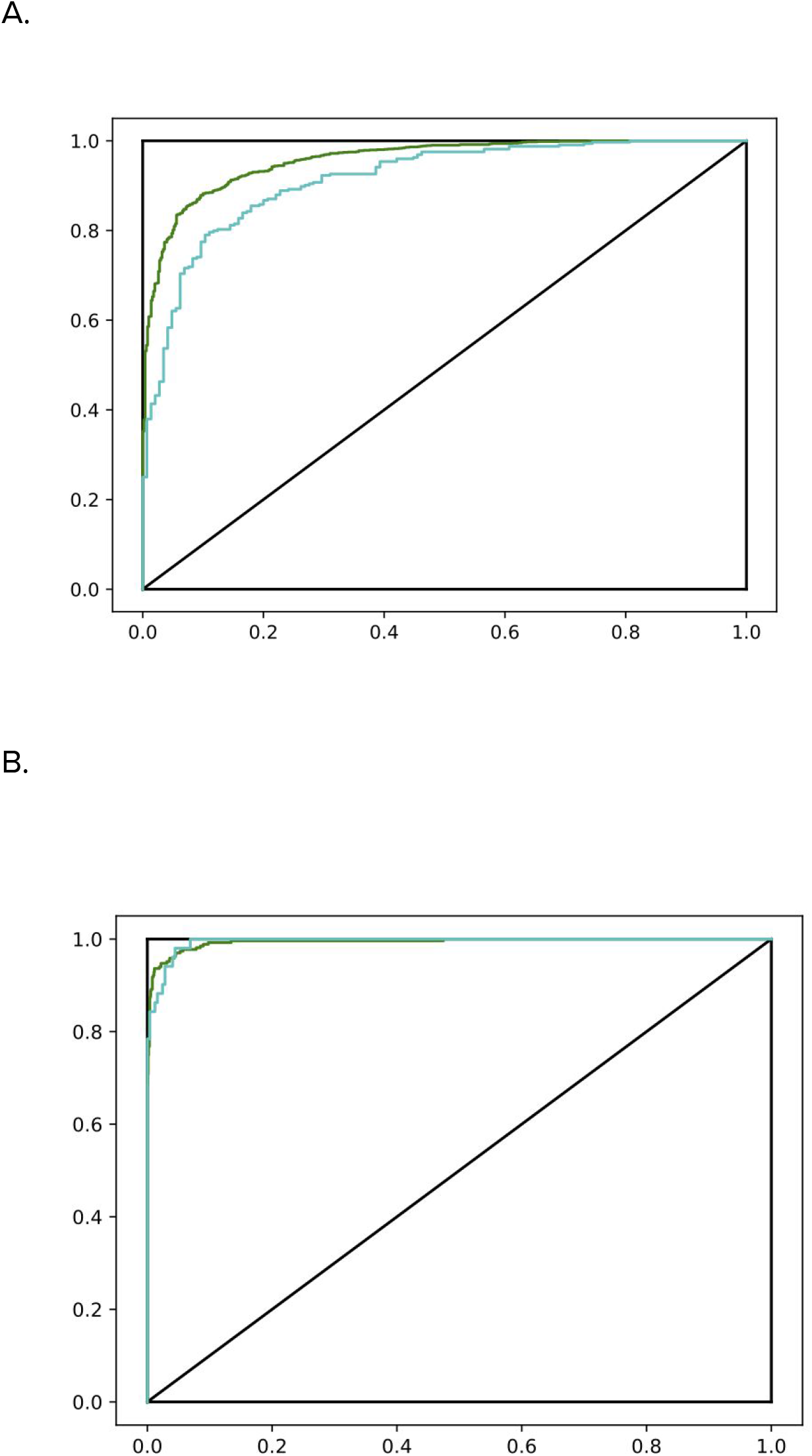
A. ROC curve measuring the accuracy of the models for eliminating bad vertebrae in step b of pipeline 2 for training dataset (green) and test dataset (green). The Cohen’s kappa values for its performance on the training set (green) and test set (blue) are 0.743 and 0.638 respectively. B. ROC curve measuring the accuracy of the models for extending vertebra to the bottom of the spine in step e of the segmentation step of pipeline 2 for training dataset (green) and test dataset (green). The Cohen’s kappa values for its performance on the training set (green) and test set (blue) are 0.917 and 0.874 respectively.

**Figure S10:**
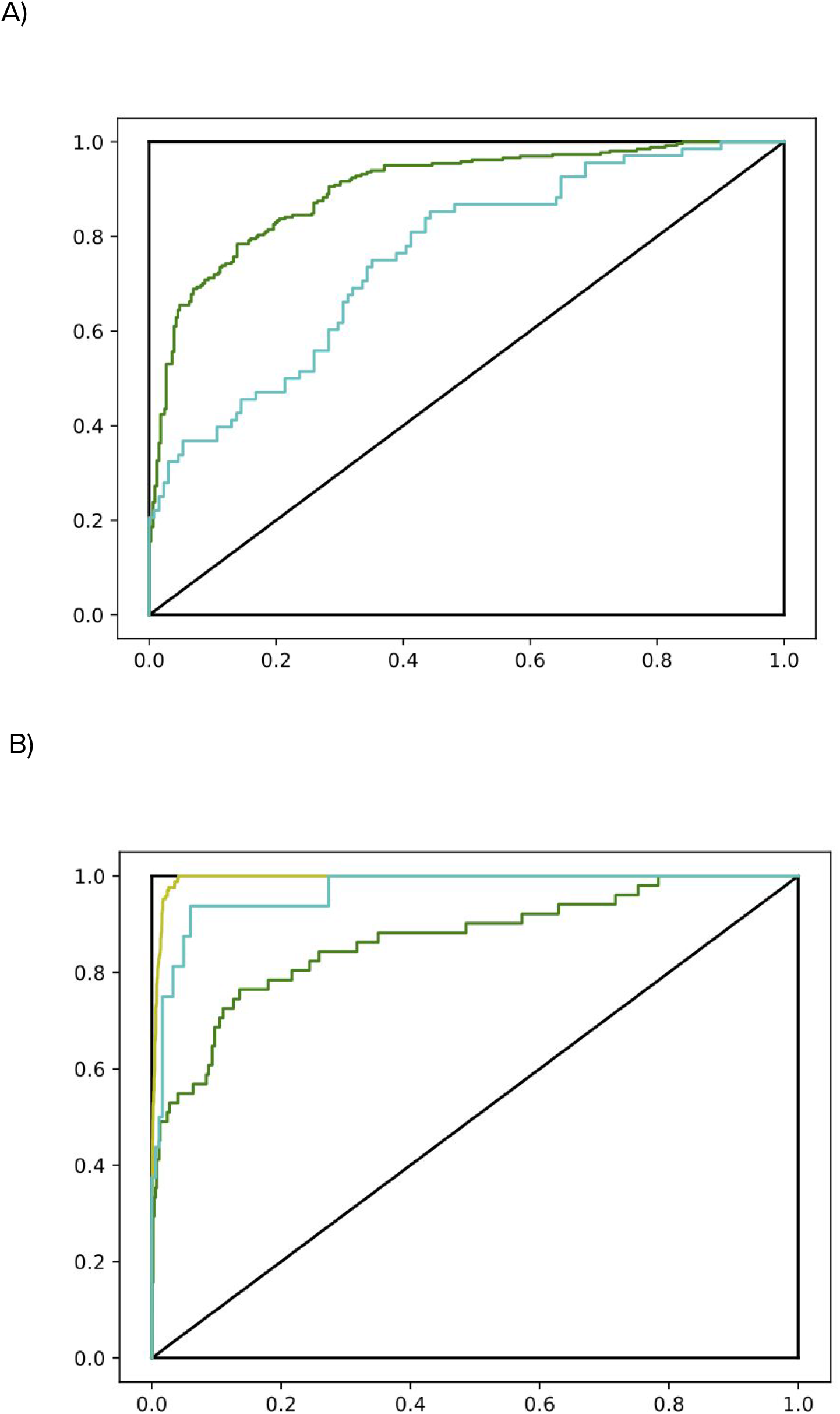
A. ROC curve measuring the accuracy of the model 1 that classifies calcified aorta from non-calcified aorta. The Cohen’s kappa values for training (green) and test sets (blue) are 0.58 and 0.33 respectively. B. ROC curve measuring the accuracy of the model 2 that classifies high-threshold aortic calcification. The original training and test sets are shown in green and blue respectively while the sandbox enriched training set is shown in yellow in B. The Cohen’s kappa values for original training, sandbox enriched training and test sets are 0.53, 0.71, and 0.73 respectively.

**Figure S11:**
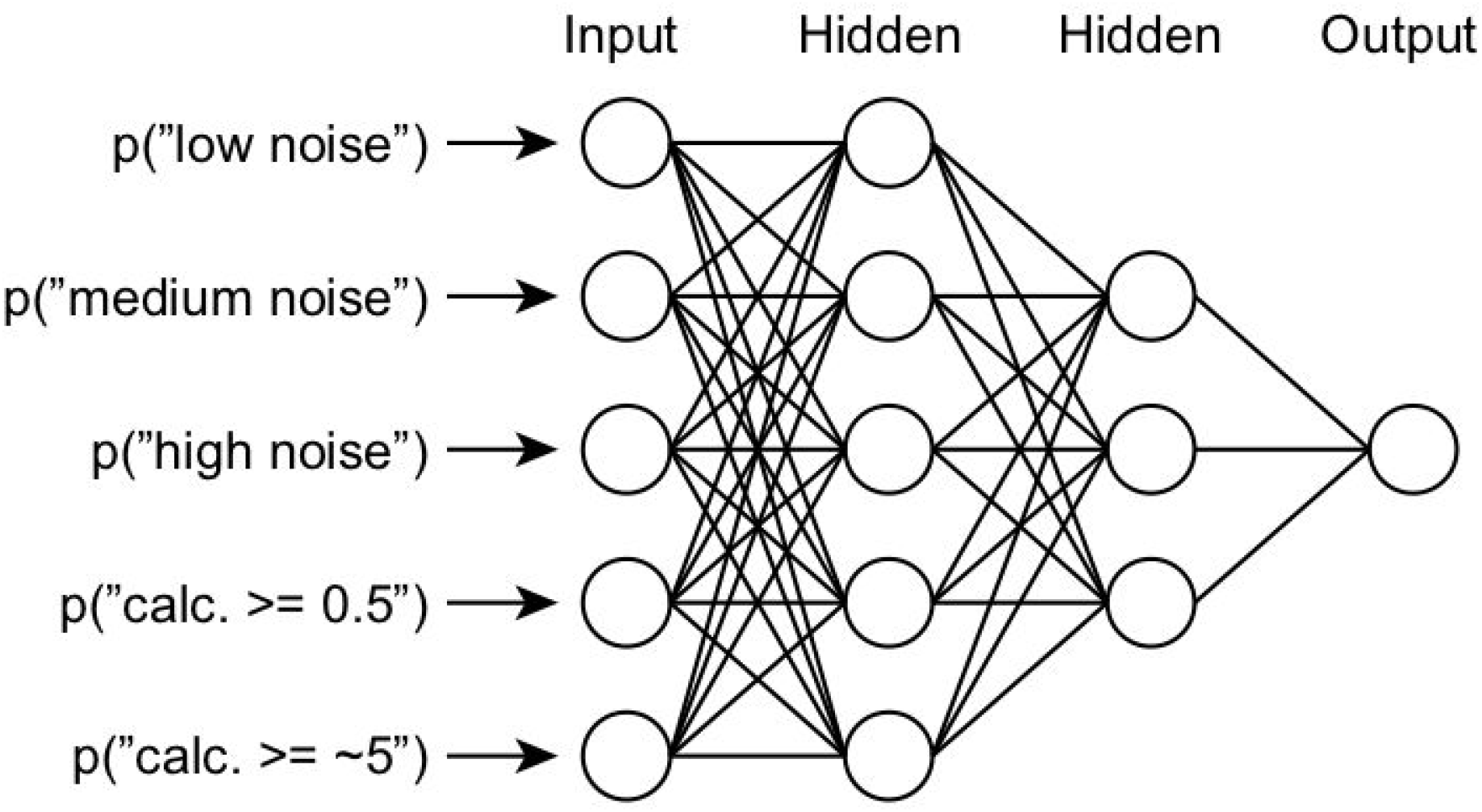
Architecture of neural network used to regress calcification values from output of models 1 and 2 for classifying calcified images from noncalcified images.

**Figure S12:**
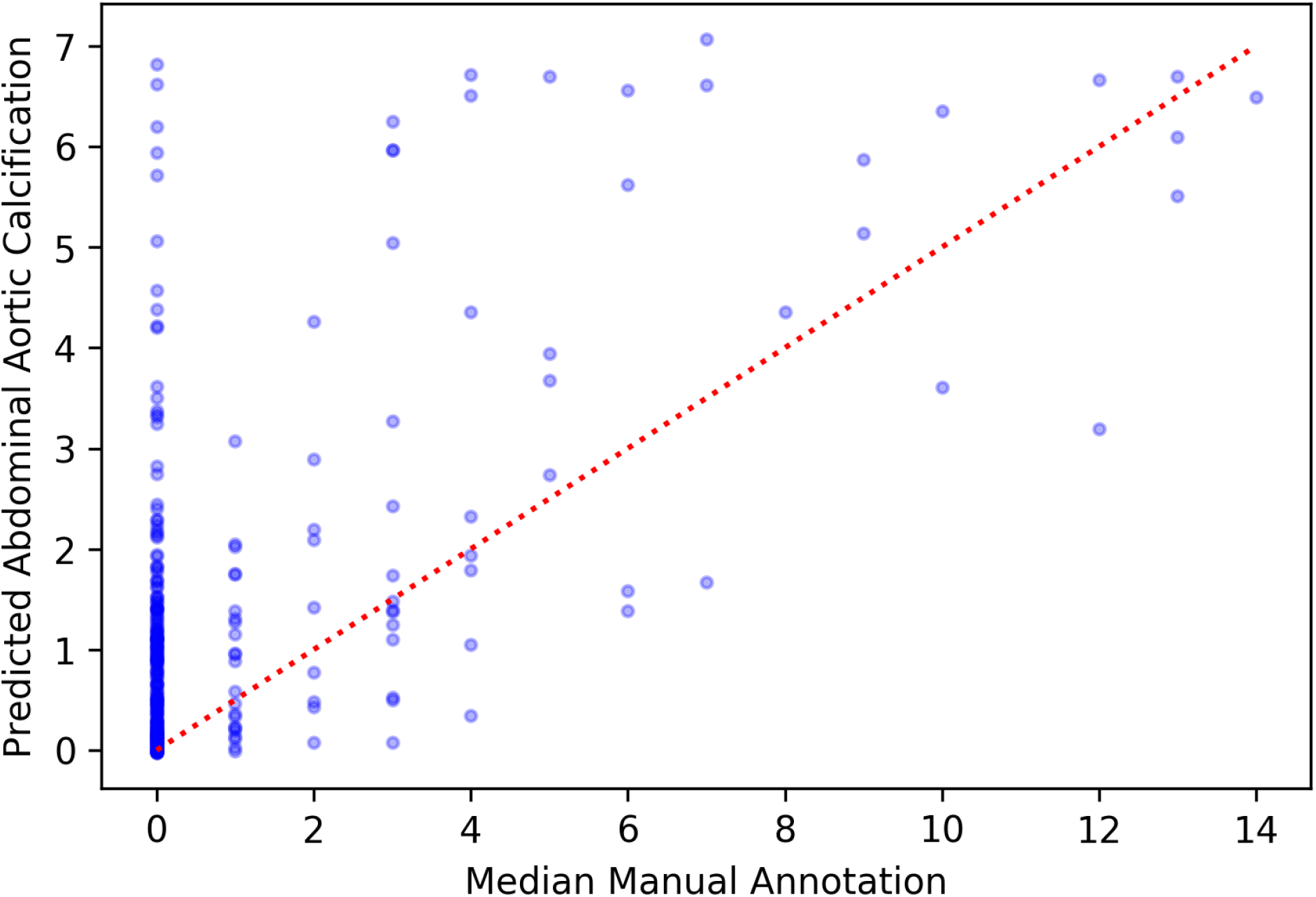
The comparison of predicted AAC to median annotation scores for the validation dataset from pipeline 2.

**Figure S13:**
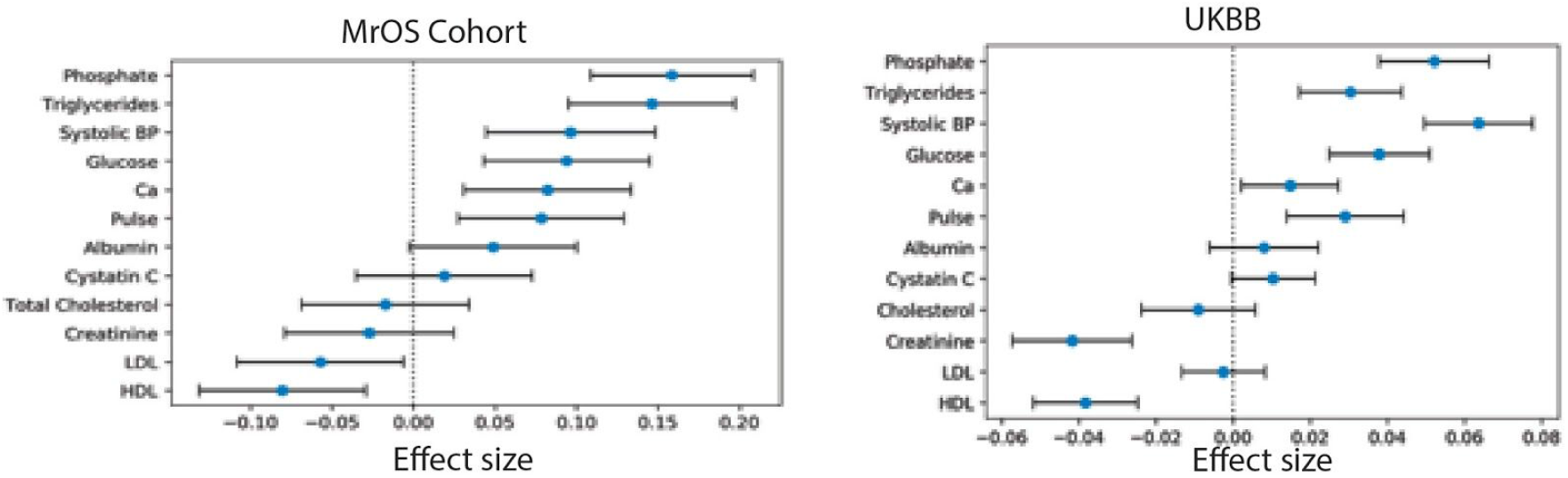
Association of biomarkers with AAC in the MrOS cohort and in this study. The biomarkers that are significantly associated with AAC in MrOS cohort are also significantly associated with predicted AAC in this study even though the effect sizes are slightly lower in the UK biobank cohort. All associations are calculated after adjusting for age and sex.

**Figure S14:**
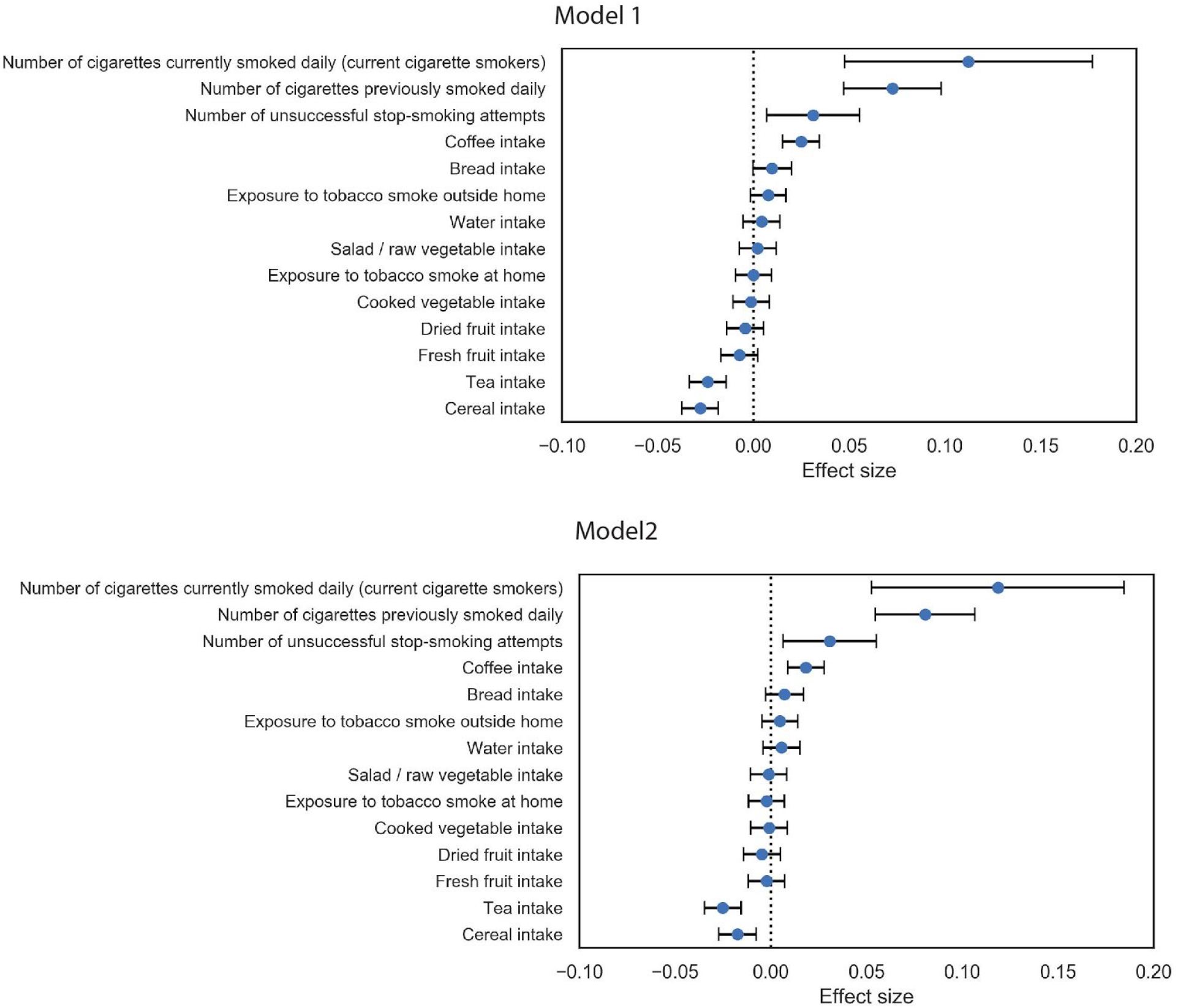
Association of predicted AAC with diet. Univariate regression analysis of risk factors at baseline for predicted AAC after adjusting for age and sex in model 1 and after adjusting for socioeconomic factors, BMI, and smoking status in addition to adjusting for age and sex in model 2. The blue dots represent mean effect size while the intervals represent standard errors for the effect size.

**Figure S15.**
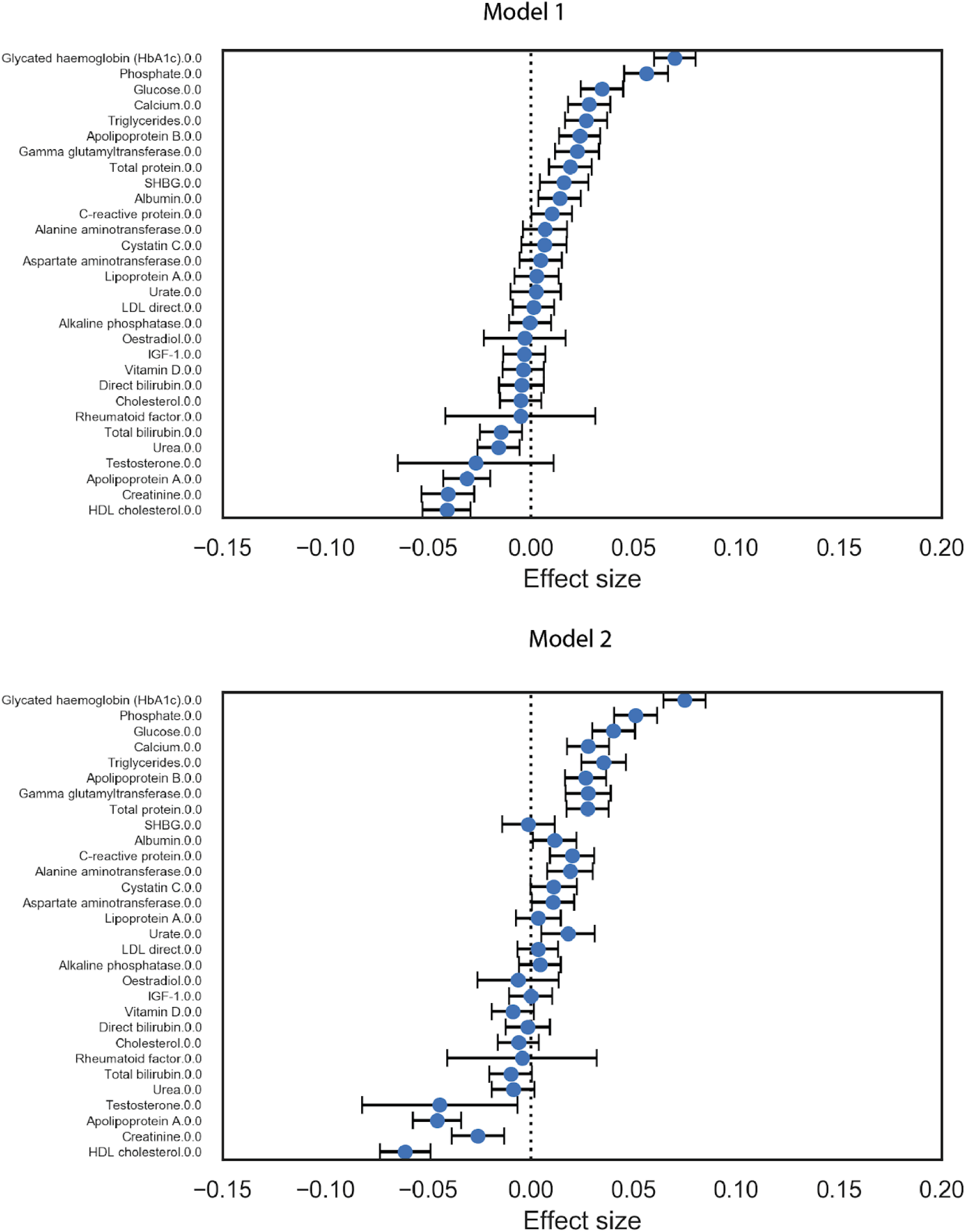
Association of AAC with all measured molecular biomarkers. Briefly, AAC is modelled as a function of each covariate adjusted for confounders (Methods). The blue dot represents the mean effect size per standard deviation of different covariates while error bars represent 95% confidence interval. All the covariates were measured during the baseline visit while AAC is estimated based on DEXA scans collected during the imaging visit.

**Figure S16.**
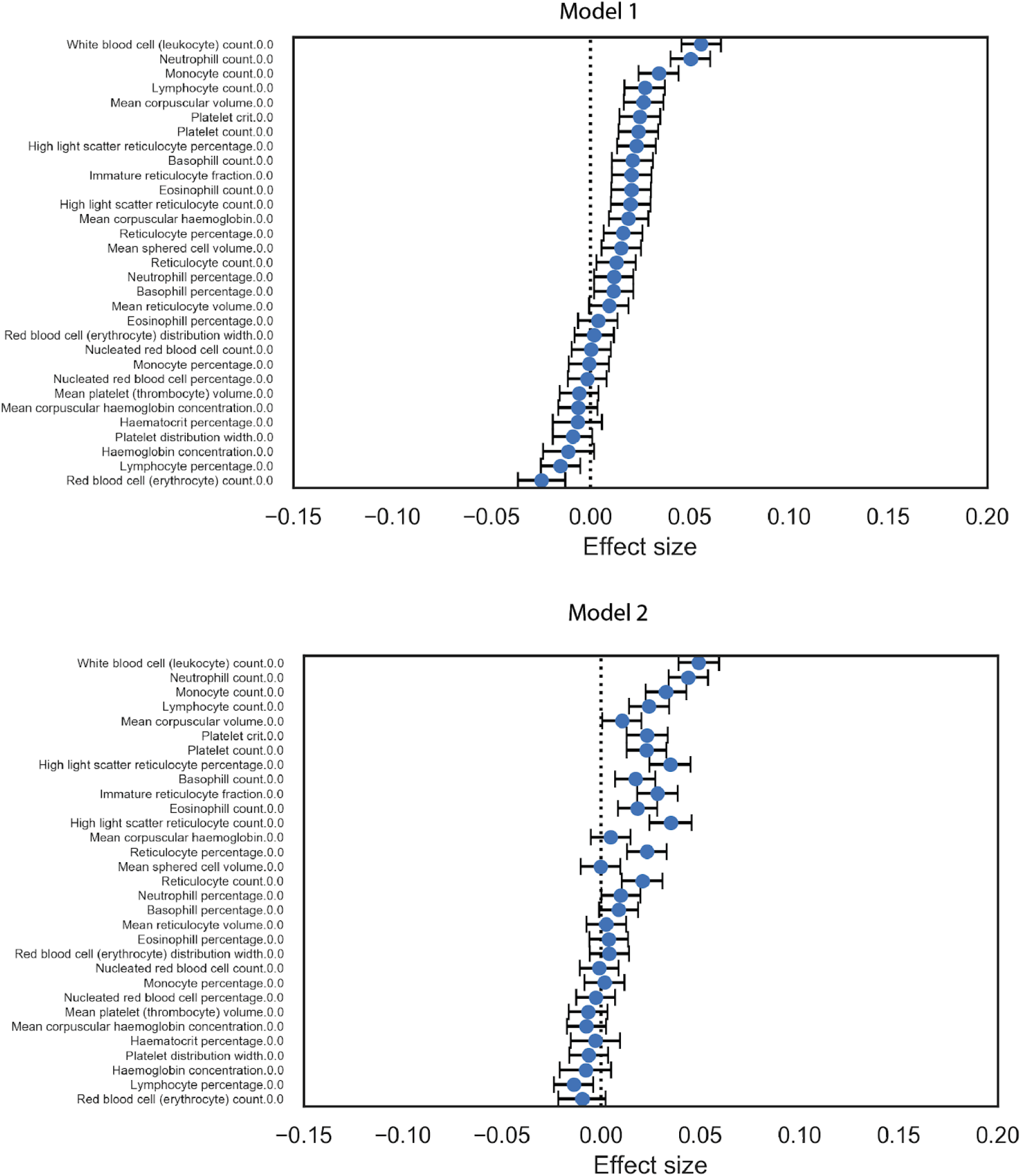
Association of AAC with all CBC markers. Briefly, AAC is modelled as a function of each covariate adjusted for confounders (Methods). The blue dot represents the mean effect size per standard deviation of different covariates while error bars represent 95% confidence interval. All the covariates were measured during the baseline visit while AAC is estimated based on DEXA scans collected during the imaging visit.

**Figure S17.**
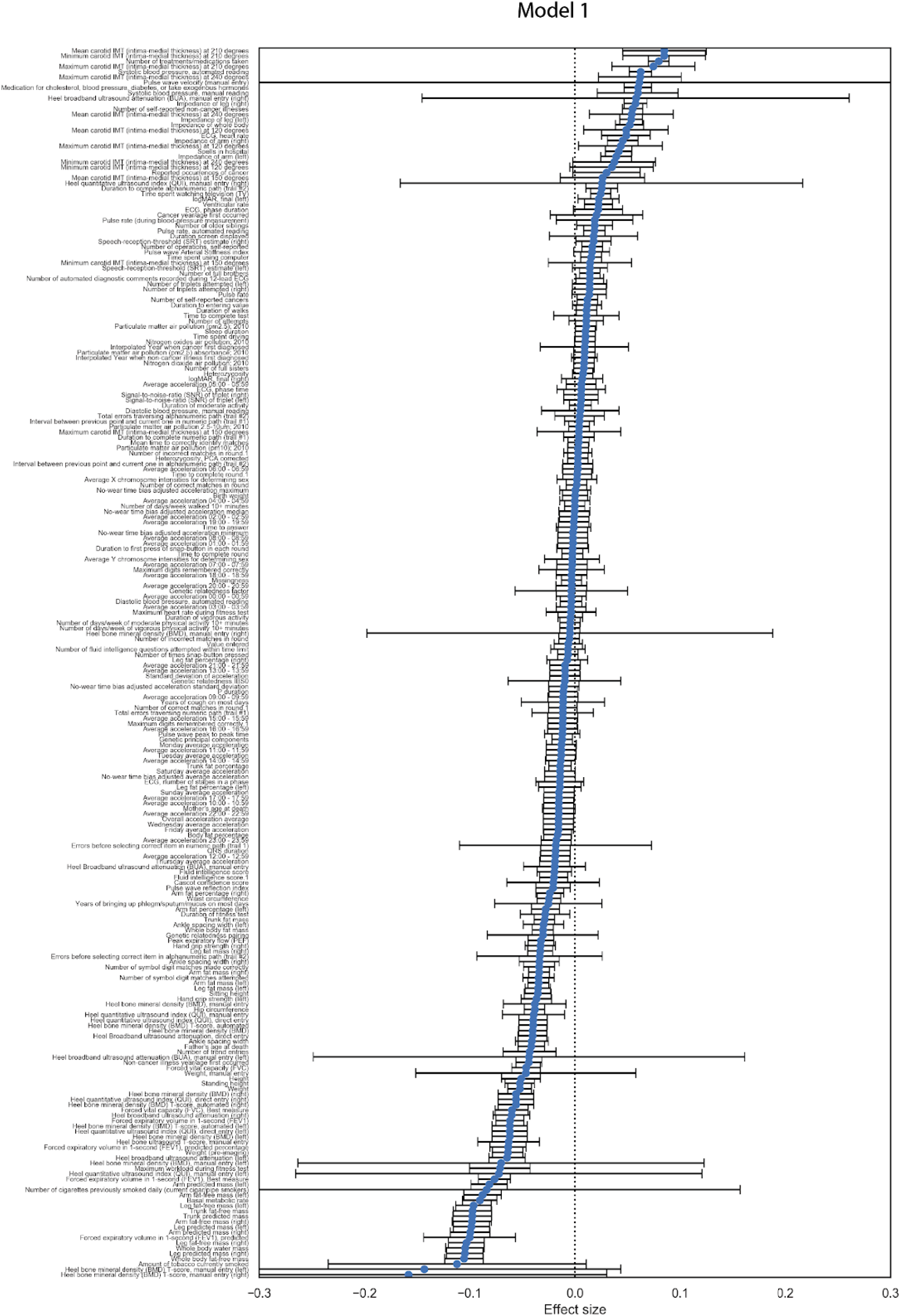
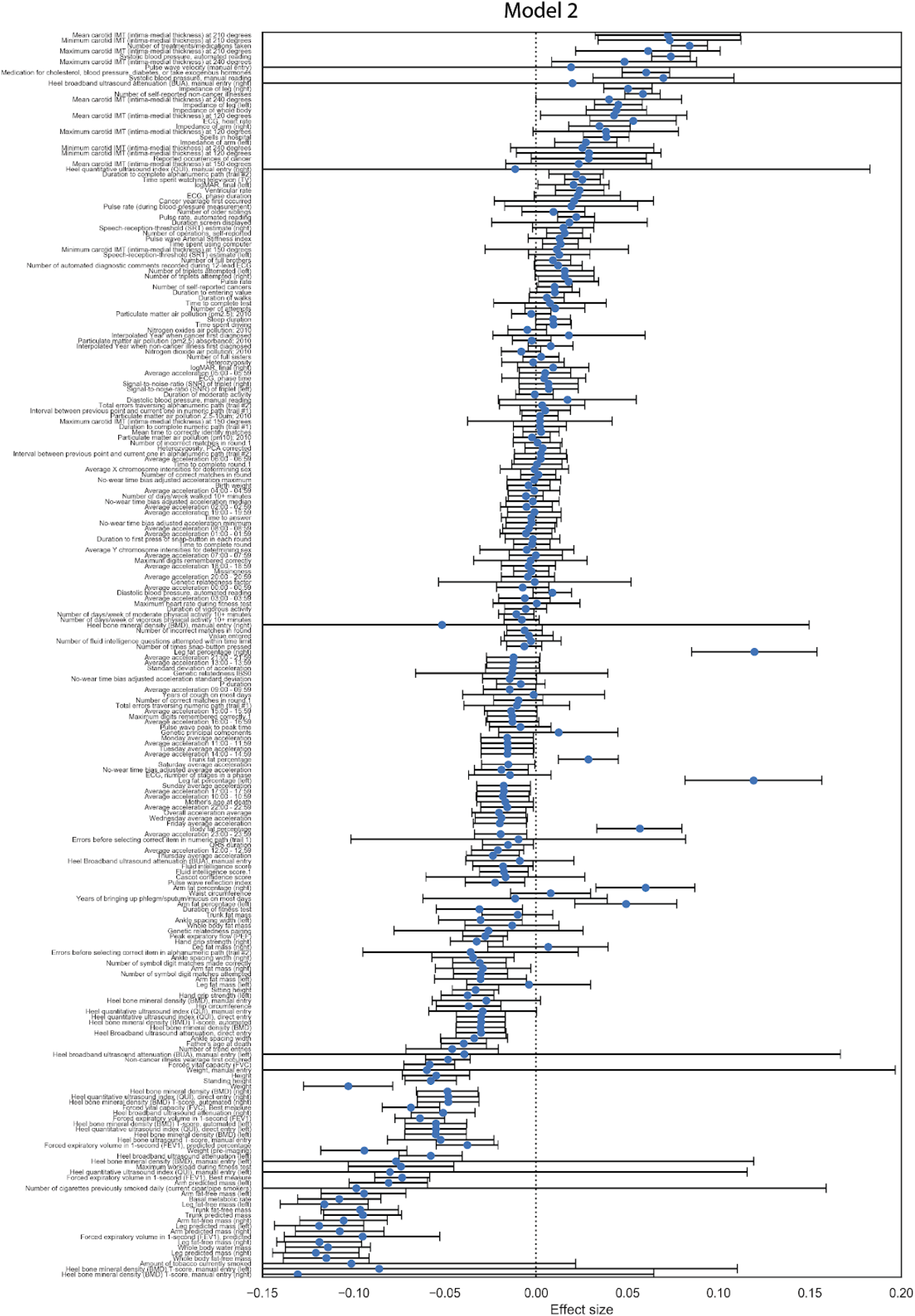
Association of AAC with all physiological measurements. Briefly, AAC is modelled as a function of each covariate adjusted for confounders (Methods). The blue dot represents the mean effect size per standard deviation of different covariates while error bars represent 95% confidence interval. All the covariates were measured during the baseline visit while AAC is estimated based on DEXA scans collected during the imaging visit.

**Figure S18:**
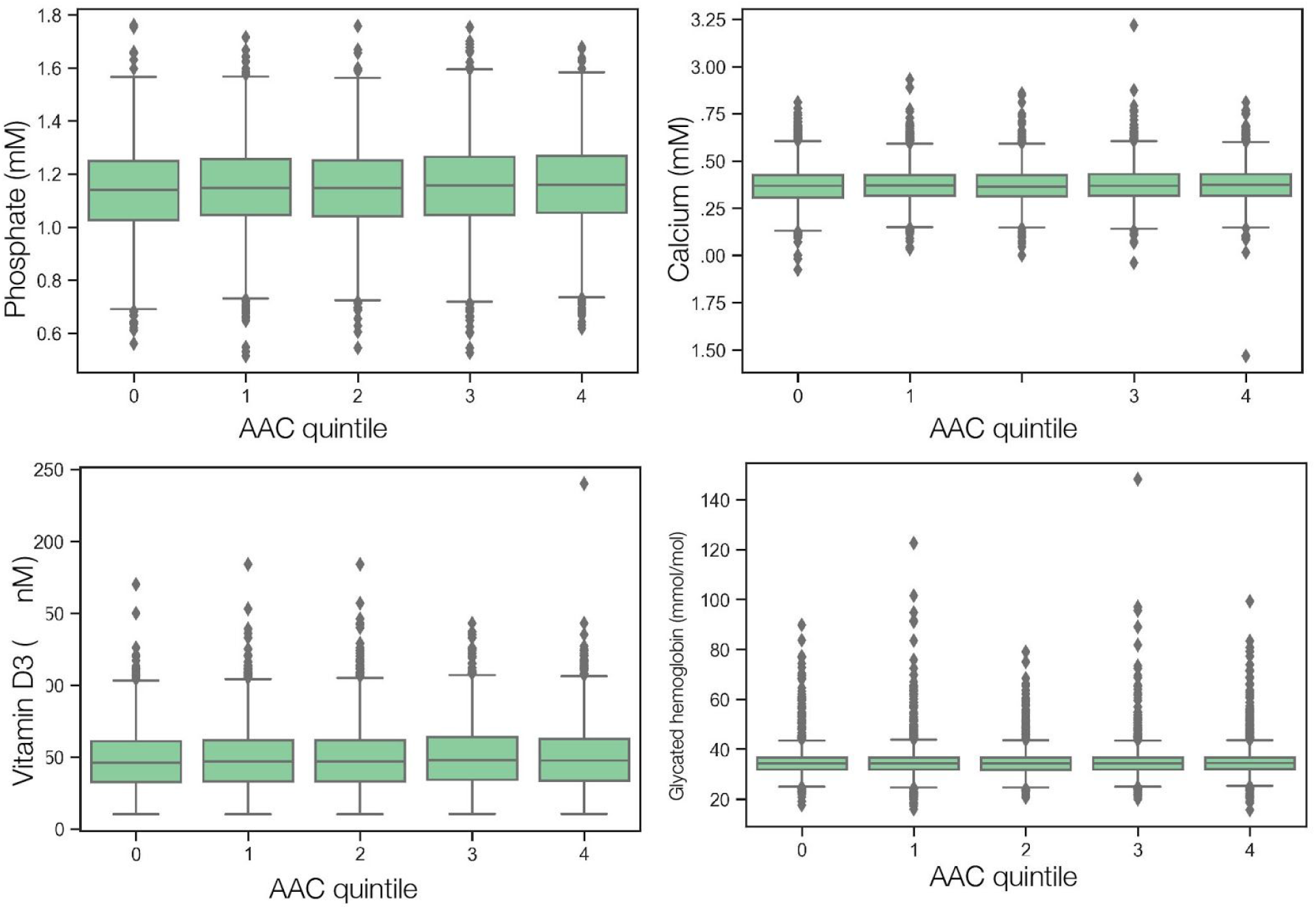
Box plot showing the level of different risk factors (serum Phosphate, Calcium, Vitamin D3, and glycated Hemoglobin HbA1c) of participants stratified according to level of calcification (higher AAC score represented by larger group number). There is very little change in the distribution of different biomarkers as a function of AAC group (except for HbA1c levels in the highest quintile of AAC) and most participants would be within clinically healthy levels for these biomarkers. The center line at each age represents the median biomarker level for all participants in that calcification category while the lower and upper boundaries of the box indicate the 25th and 75th percentile of the serum biomarker level for that calcification category. The lower and upper ends of the lines denote the 95% confidence interval of the serum biomarker level for that calcification category.

**Figure S19:**
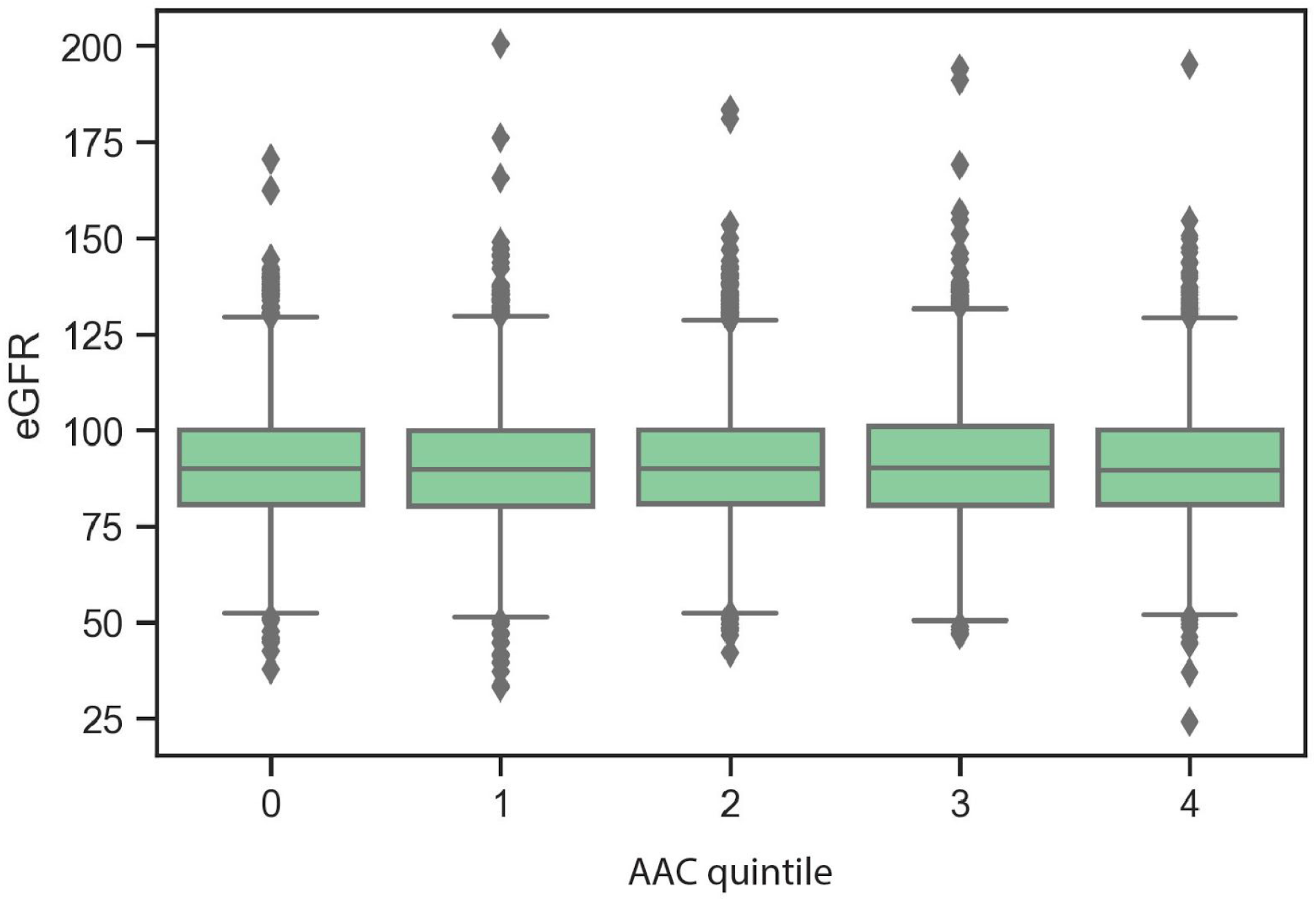
Box plot showing the variation of estimated glomerular filtration rate (eGFR) of participants grouped according to level of (higher AAC score represented by larger group number). The eGFR was calculated from Cystatin-C levels using the formula (Grubb et al. 2014). The eGFR for the participants with the highest levels of AAC (categories 3 and 4) tend to be lower, indicating that their kidneys are not functioning as efficiently as the other participants on average. The center line at each age represents the median biomarker level for all participants in that calcification category while the lower and upper boundaries of the box indicate the 25th and 75th percentile of the eGFR for that calcification category. The lower and upper ends of the lines denote the 95% confidence interval of the eGFR for participants within a particular calcification category.

**Figure S20.**
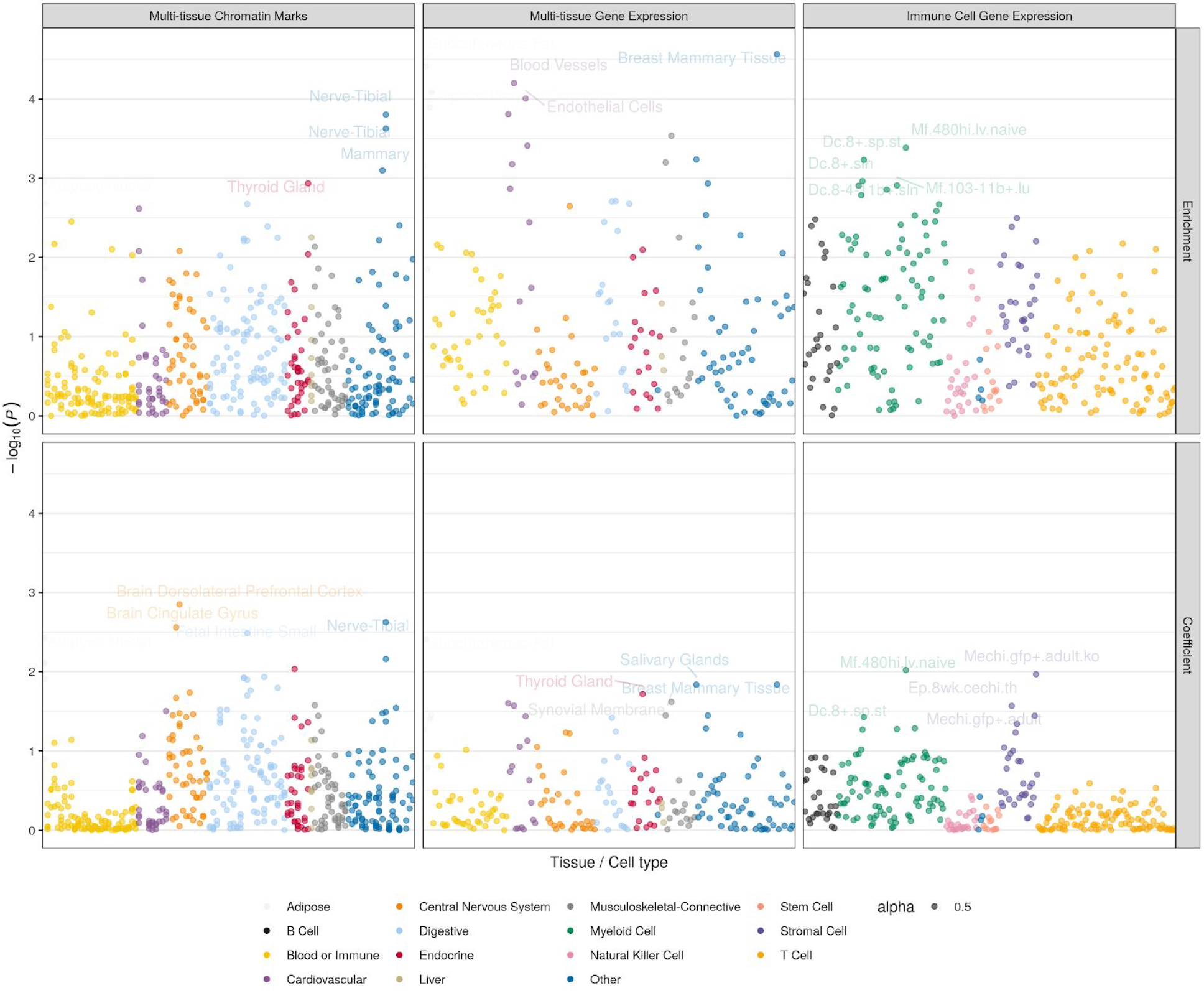
Heritability enrichment of AAC across different genomic annotations. P-values (y-axis) of either total enrichment (top row facet) or positive regression coefficients (tau; bottom row facet) across annotations (x-axis) and datasets (column facets). Enrichment is calculated by dividing the proportion of heritability explained by SNPs in an annotation by proportion of SNPs in an annotation (proportion_h2_/proportion_SNPs_). Regression coefficients represent the average contribution of an annotation to per-SNP heritability, correcting for annotations in the baseline model (Methods).

**Figure S21.**
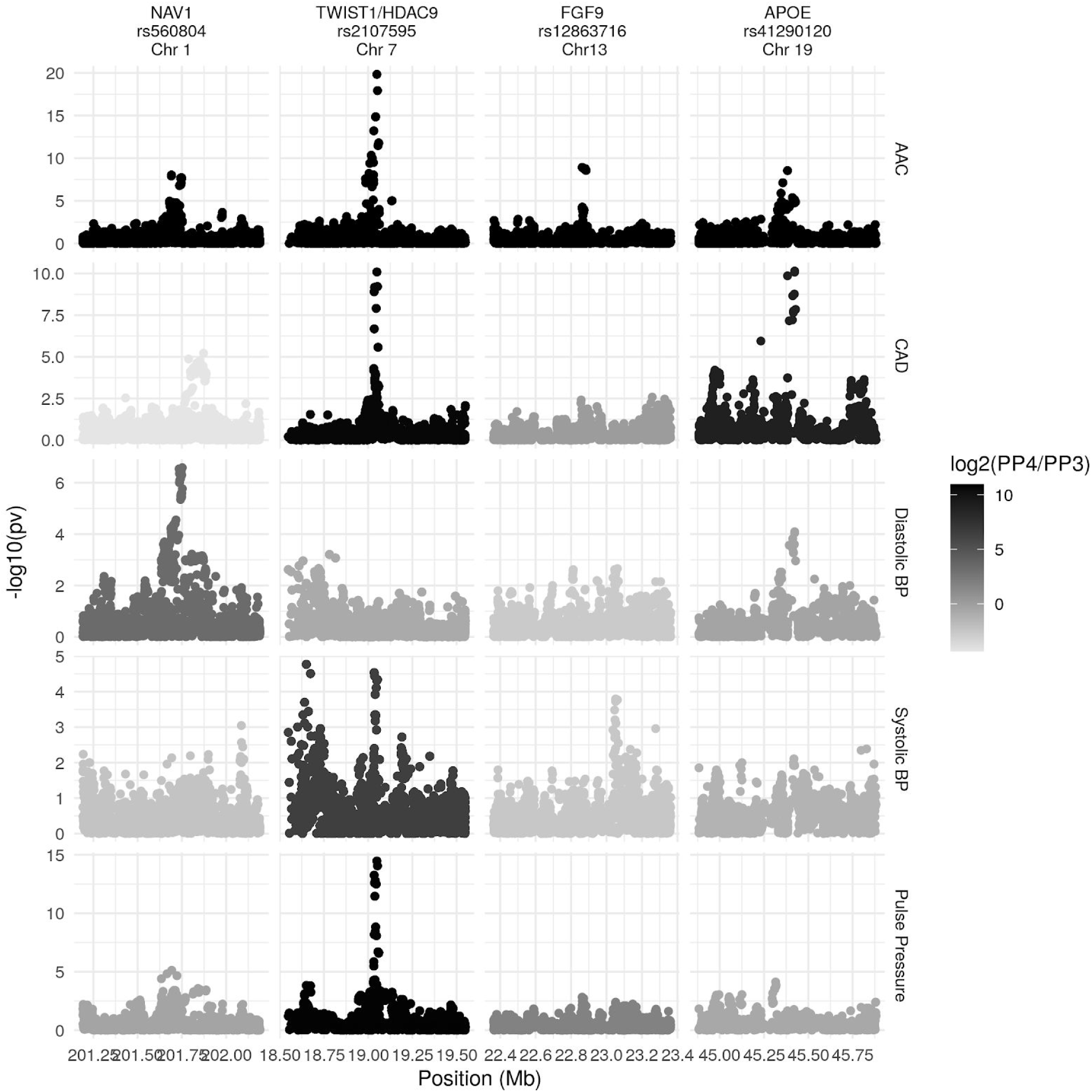
Colocalization of the lead signal at four loci where we find an association with AAC (top row) in non-UKBB cohorts. CAD=Coronary artery disease. BP=blood pressure. CAD summary statistics are from(Nikpay et al. 2015) and other traits are from (Hoffmann et al. 2017).

**Figure S22:**
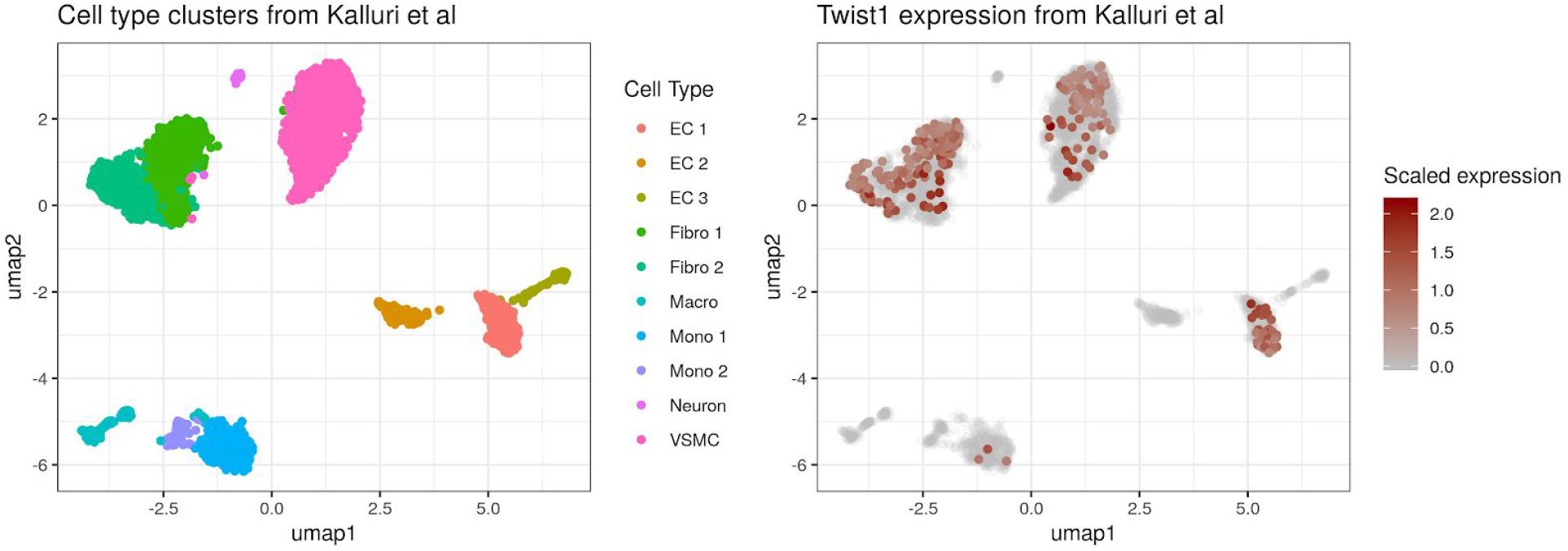
Expression of *Twist1* in mouse aorta cell subtypes, with clusters as identified in (Kalluri et al. 2019). EC1 corresponds to a subset of endothelial cells characterized by expression of genes involved in extracellular matrix organization. This corresponds to a cluster identified in (Lukowski et al. 2019) as having a mesenchymal phenotype, quiescence, and high mitochondrial content, preceding the others two more differentiated states in pseudotime.

**Figure S23:**
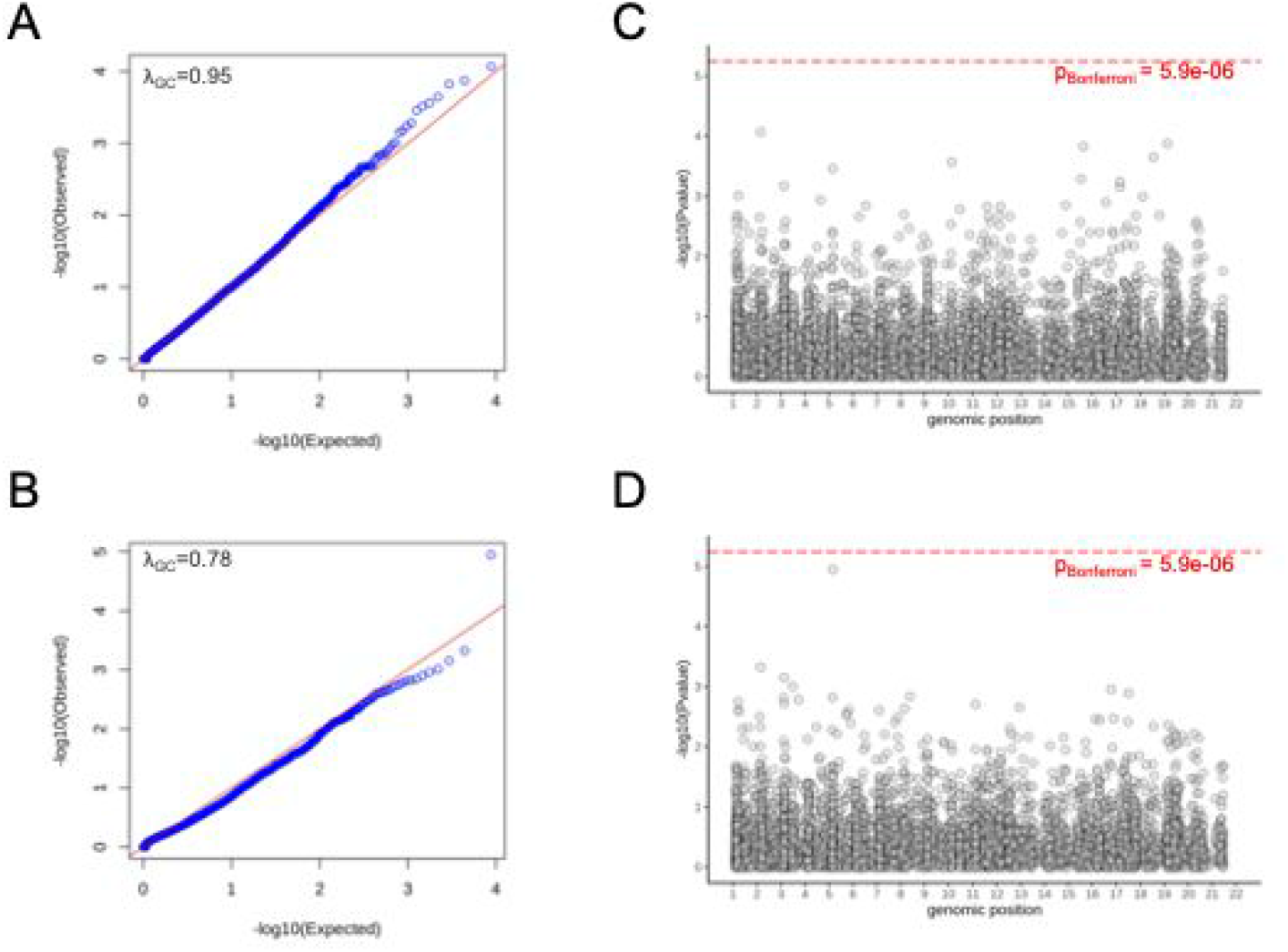
Rare variant association study (RVAS) of abdominal aortic calcification (AAC) in the UK Biobank. (A) Q-Q plot demonstrates calibration of optimized test statistics combining burden test and SKAT, as implemented in SAIGE-GENE, for a model in which the outcome variable is rank normalized AAC values for n=11,749 samples of European ancestry with both AAC quantification and exome sequence data. Predicted loss-of-function variants were grouped by gene. For details on the regression modelling, see Supplementary Methods. (B) Q-Q plot depicting association study with dichotomized outcomes. In the dichotomous RVAS, cases were defined by raw AAC score >=3 and controls were defined by raw AAC score <3. There were 1,274 cases and 10,475 controls (see Supplementary Methods). (C) Manhattan plot of association study from the continuous RVAS. (D) Manhattan plot of the association study from the dichotomous RVAS. There were no associations below Bonferroni significance threshold in either study.

**Figure S24:**
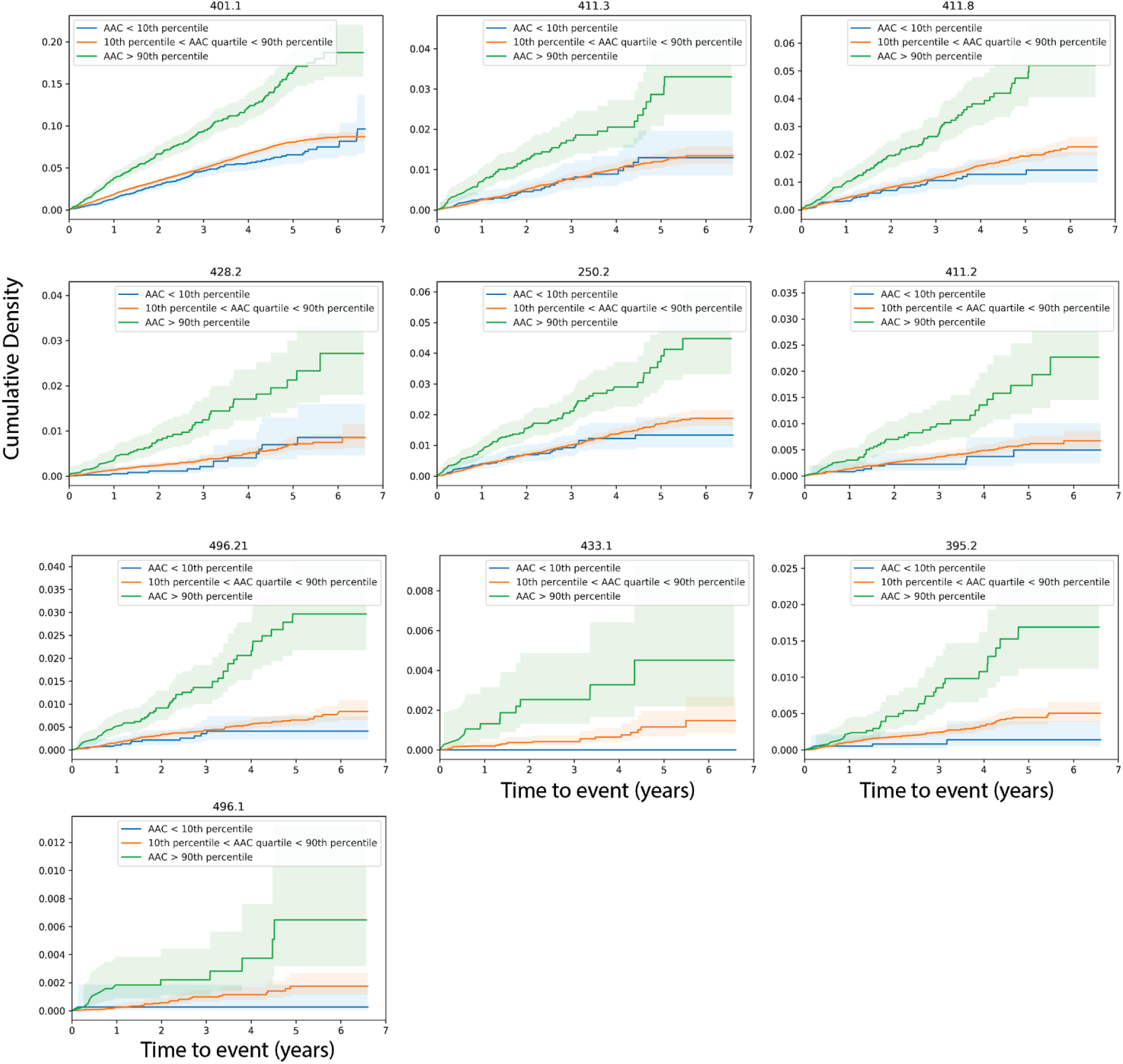
Cumulative density plots of event occurrence. The cumulative density plots of different event types that showed statistically significant disease associations after multiple hypothesis testing (p-value_Bonferroni_<0.05) using Cox proportional hazards. The green/red/blue lines indicates the cumulative incidents for the cohort stratified based on AAC levels. The shaded regions represent the 95% confidence interval for each subset of participants.

**Figure S25:**
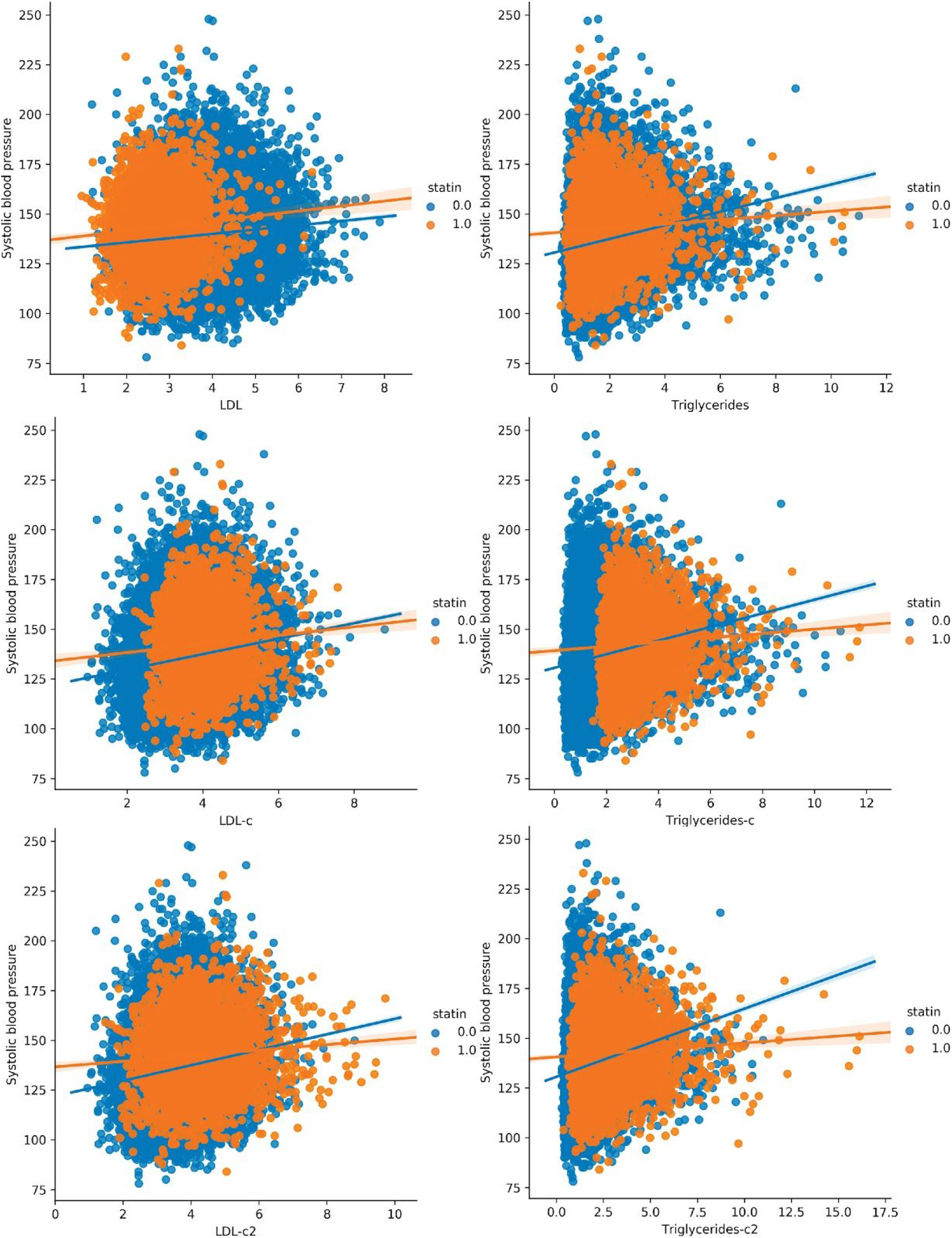
Effect of statins on LDL-Blood Pressure relationship. The relationship between baseline LDL and Triglyceride levels to blood pressure is plotted for all UK biobank participants before and after statin correction (c1 and c2 represent the two statin correction methods). Blue represents people who don’t take statin medication are shown in blue while statin takers are shown in orange. The uncorrected LDL levels are lower than that predicted by blood pressure for statin takers and the correction methods reduce this discrepancy.

**Figure S26:**
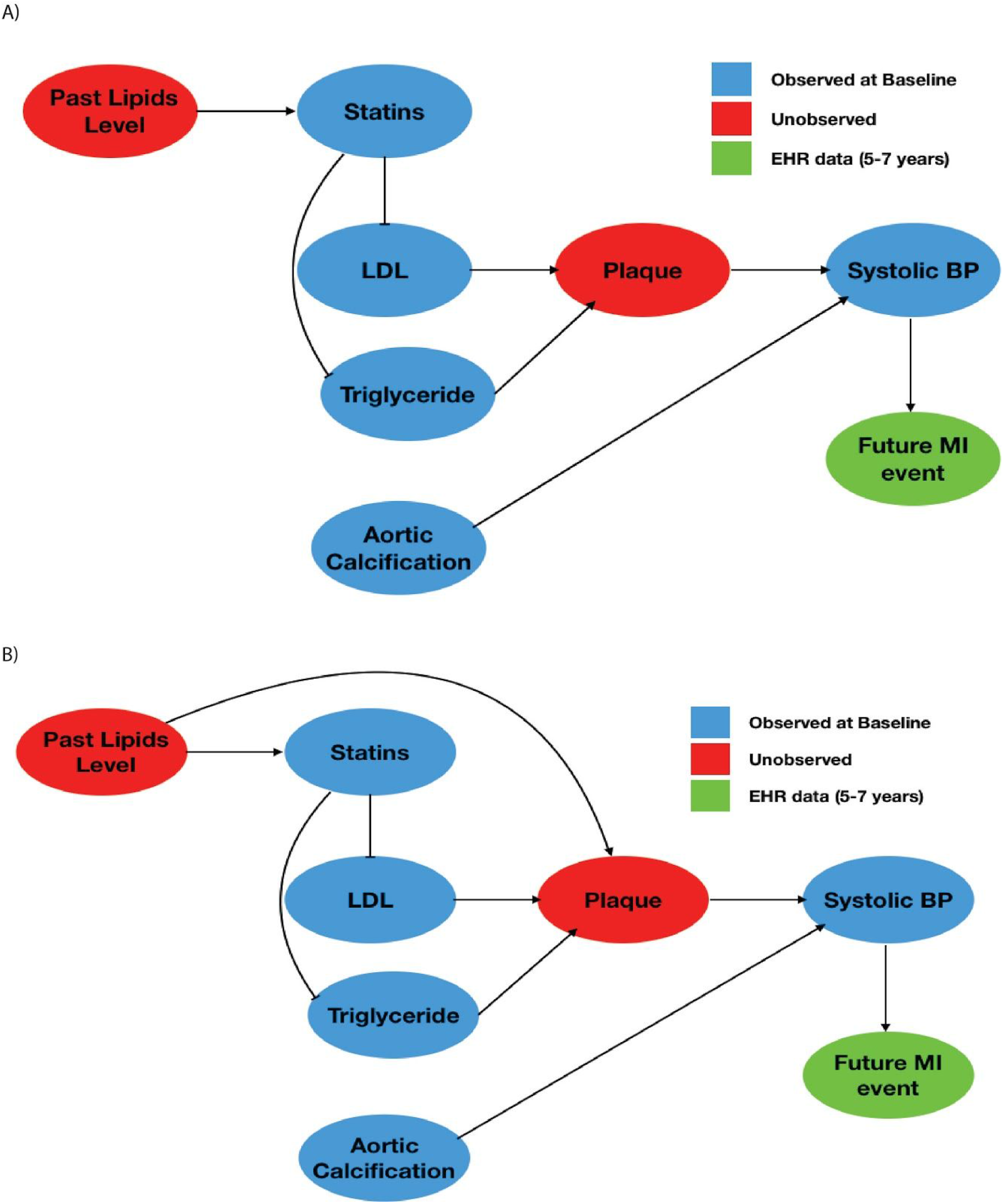
Graphical Models for MI event based on LDL and AAC. The relationship between different lipid levels and their predictions of future MI events. LDL and triglyceride levels affect plaque formation increasing systolic blood pressure and the probability of future myocardial infarction events. Aortic calcification forms a parallel independent pathway for systolic blood pressure increase. Statins reduce LDL and triglyceride levels and in (A) do not affect acute MI events as they only affect blood pressure through LDL levels. However, in **Figure S25**, we show that statin takers do have higher blood pressure than that predicted by LDL levels alone and this is because statins do not remove old plaque that is determined by past lipid levels (B). Hence, we apply statin correction to LDL levels to get an unbiased estimate of LDL risk and compare it to Aortic Calcification risk in this article.

**Figure S27:**
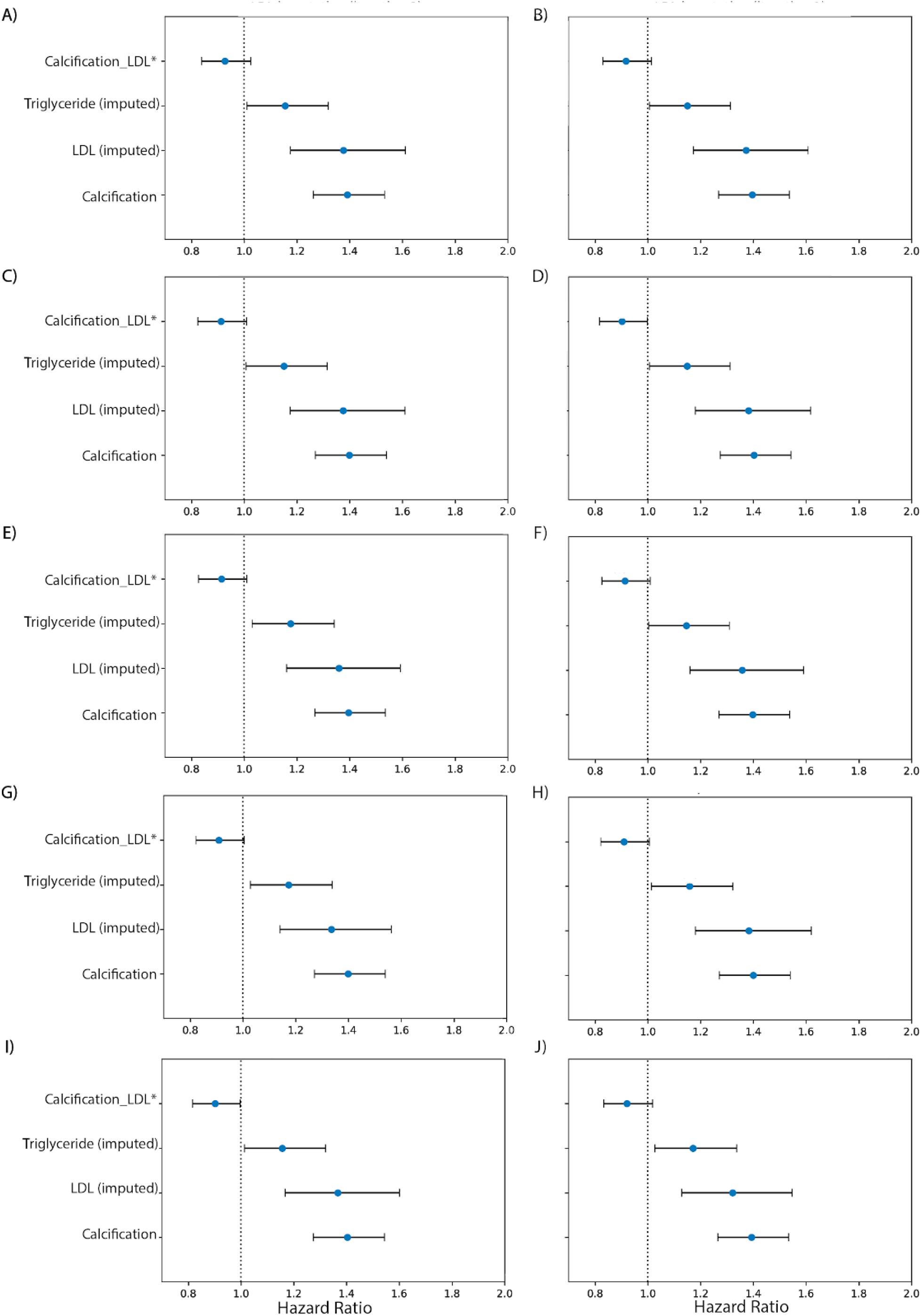
Consistency of LDL imputation procedure for comparing LDL risk to Calcification risk. Ten random initializations with MICE were used to impute the LDL level for statin users while LDL levels for non-statin users were used unadjusted. Cox proportional hazards were used to compare the hazard from LDL and aortic calcification over the UK biobank population. In spite of the variability from imputation, the hazard ratios for LDL and calcification risk levels were similar across all 10 models.

**Figure S28.**
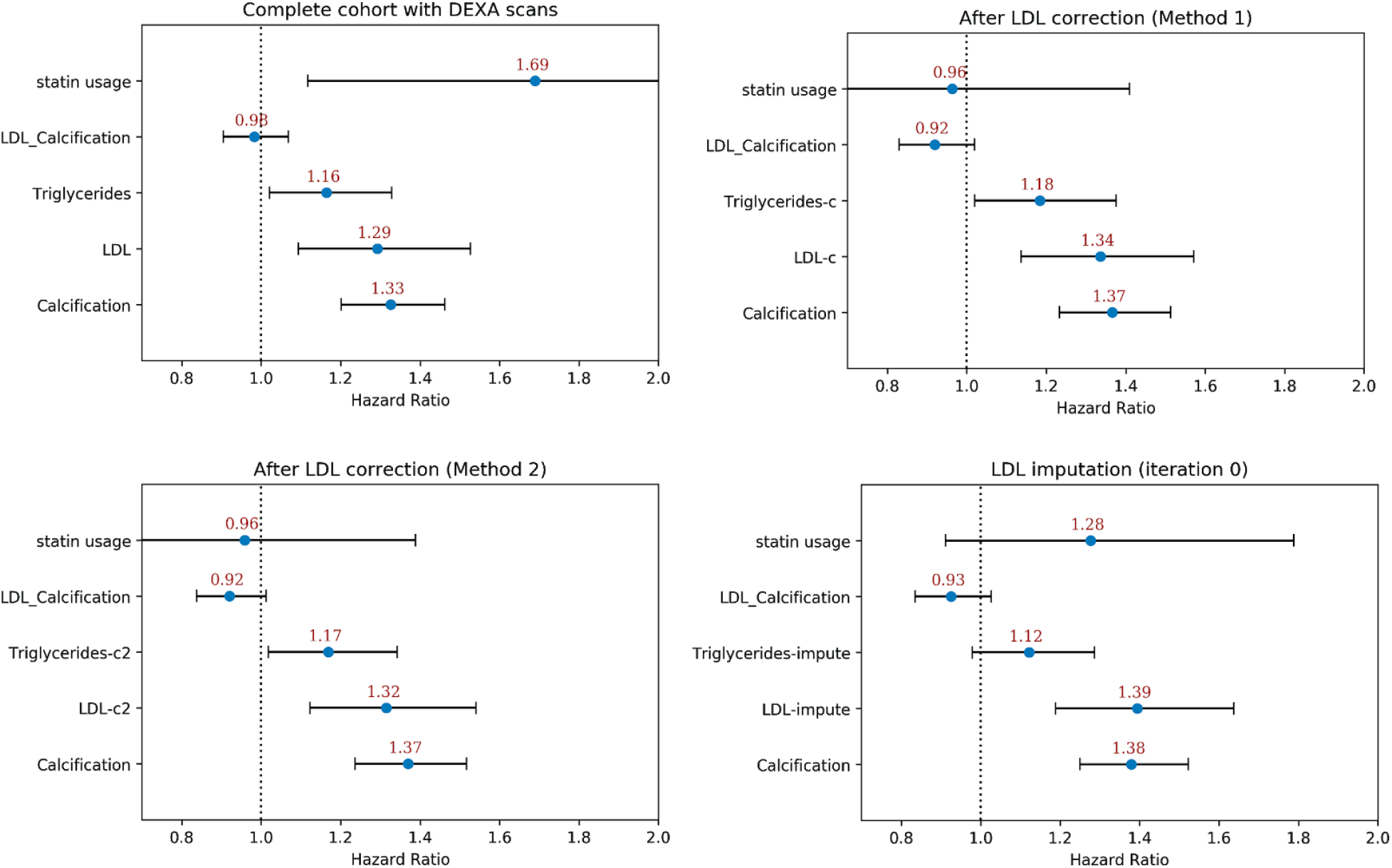
Statin usage status in CoxPH association of AAC and LDL for acute MI events. We compared the hazard ratios for statin usage, AAC, LDL, and triglycerides using a multivariate CoxPH model with age and sex covariates. (A) Whole cohort - no statin adjustment. (B) Whole cohort after adjusting for statin using method 1 - assumes that LDL levels are reduced by 1.25 mmol/L in statin users. (C) Whole cohort method 2 - assumes that LDL levels are reduced by 35% in statin users. (D) Whole cohort imputation of LDL levels based on remaining biomarkers and physiological measurements (Methods). The blue dots represent mean hazard ratio, while error bars represent 95% confidence interval.

**Figure S29.**
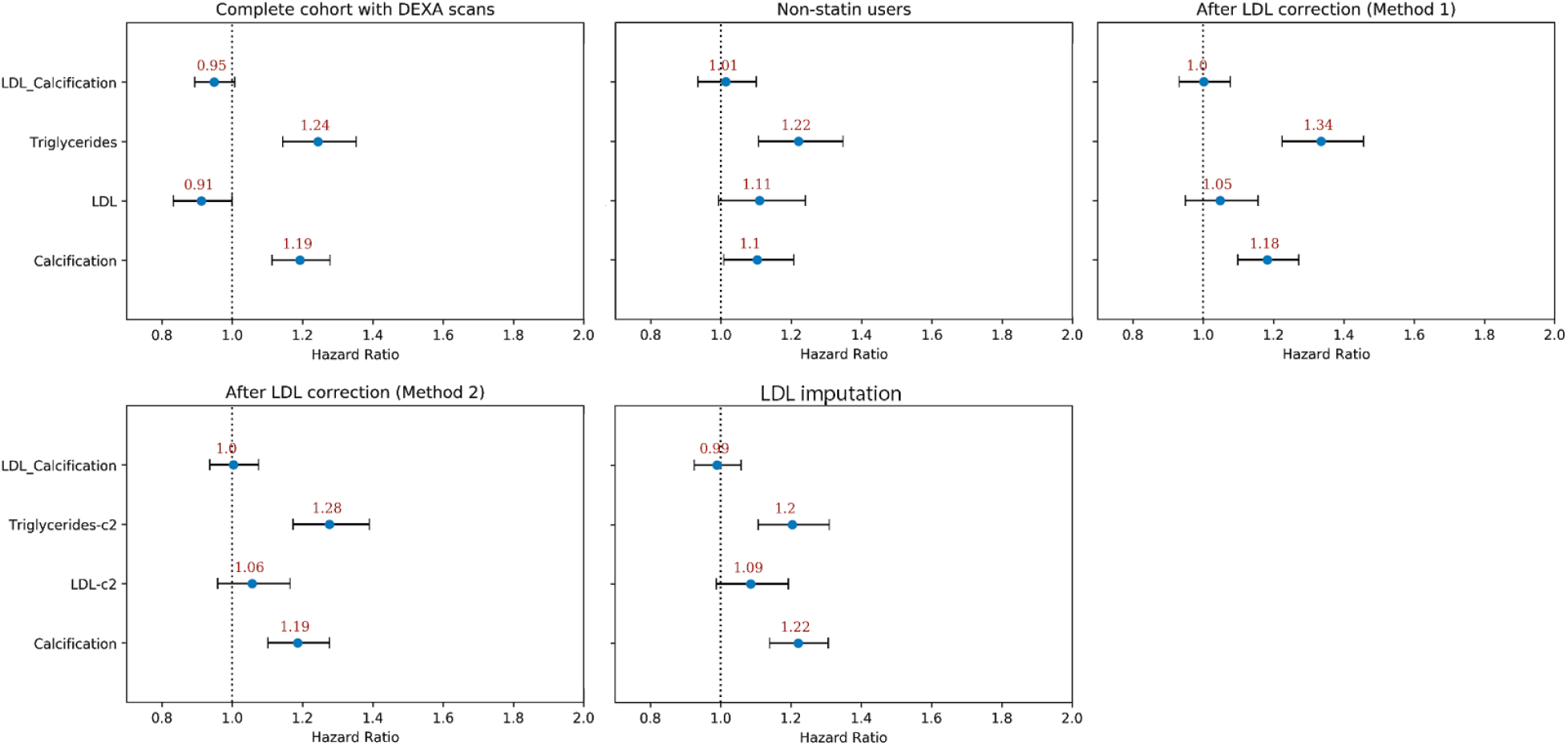
CoxPH association of AAC and LDL for CVD events post imaging visit. The CVD events represent composite outcomes from MI, angina pectoris, heart failure, ischemic heart disease, and occlusion and stenosis of precerebral arteries. We compared the hazard ratios for AAC, LDL, and triglycerides using a multivariate CoxPH model with age and sex covariates. (A) Whole cohort - no statin adjustment. (B) Subset of only non-statin users. (C) Whole cohort after adjusting for statin using method 1 - assumes that LDL levels are reduced by 1.25 mmol/L in statin users. (D) Whole cohort method 2 - assumes that LDL levels are reduced by 35% in statin users. (E) Whole cohort imputation of LDL levels based on remaining biomarkers and physiological measurements (Methods). The blue dots represent mean hazard ratio, while error bars represent 95% confidence intervals. Interaction term between LDL and calcification is starred. **This analysis was done with 2**.**9 years of median follow up time post imaging visit. LDL was measured during the baseline visit**.

### Supplementary Tables

**Table S1:**
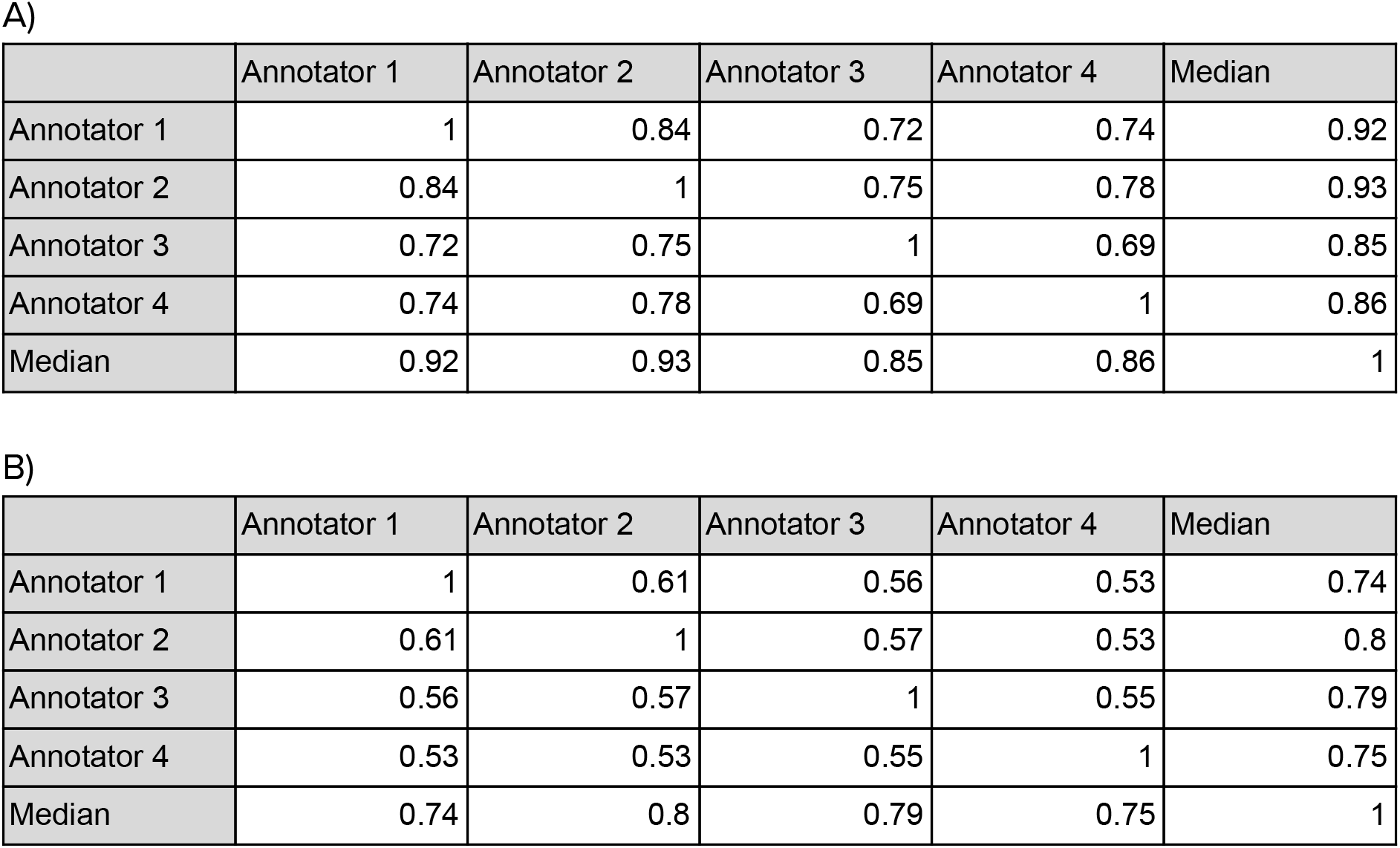
Variability of manual annotation over the training set for the 4 different annotators measured using (A) Pearson Correlation and (B) Spearman correlation.

|  | Annotator 1 | Annotator 2 | Annotator 3 | Annotator 4 | Median |
| --- | --- | --- | --- | --- | --- |
| Annotator 1 | 1 | 0.84 | 0.72 | 0.74 | 0.92 |
| Annotator 2 | 0.84 | 1 | 0.75 | 0.78 | 0.93 |
| Annotator 3 | 0.72 | 0.75 | 1 | 0.69 | 0.85 |
| Annotator 4 | 0.74 | 0.78 | 0.69 | 1 | 0.86 |
| Median | 0.92 | 0.93 | 0.85 | 0.86 | 1 |

|  | Annotator 1 | Annotator 2 | Annotator 3 | Annotator 4 | Median |
| --- | --- | --- | --- | --- | --- |
| Annotator 1 | 1 | 0.61 | 0.56 | 0.53 | 0.74 |
| Annotator 2 | 0.61 | 1 | 0.57 | 0.53 | 0.8 |
| Annotator 3 | 0.56 | 0.57 | 1 | 0.55 | 0.79 |
| Annotator 4 | 0.53 | 0.53 | 0.55 | 1 | 0.75 |
| Median | 0.74 | 0.8 | 0.79 | 0.75 | 1 |

**Table S2:** Variability of manual annotation over the validation set for the 4 different annotators measured using (A) Pearson Correlation and (B) Spearman correlation.

|  | Annotator 1 | Annotator 2 | Annotator 3 | Annotator 4 | Median |
| --- | --- | --- | --- | --- | --- |
| Annotator 1 | 1 | 0.86 | 0.78 | 0.75 | 0.94 |
| Annotator 2 | 0.86 | 1 | 0.73 | 0.67 | 0.93 |
| Annotator 3 | 0.78 | 0.73 | 1 | 0.7 | 0.85 |
| Annotator 4 | 0.75 | 0.67 | 0.7 | 1 | 0.73 |
| Median | 0.94 | 0.93 | 0.85 | 0.73 | 1 |

|  | Annotator 1 | Annotator 2 | Annotator 3 | Annotator 4 | Median |
| --- | --- | --- | --- | --- | --- |
| Annotator 1 | 1 | 0.68 | 0.67 | 0.56 | 0.83 |
| Annotator 2 | 0.68 | 1 | 0.71 | 0.51 | 0.88 |
| Annotator 3 | 0.67 | 0.71 | 1 | 0.55 | 0.84 |
| Annotator 4 | 0.56 | 0.51 | 0.55 | 1 | 0.57 |
| Median | 0.83 | 0.88 | 0.84 | 0.57 | 1 |

**Table S3:** Correlation of annotated abdominal aortic calcification with different phenotypes across MrOS and UKBB cohorts. Only phenotypes significantly correlated with aortic calcification in MrOS cohort are displayed in the above table.

|  | MrOS Cohort | UKBB Cohort |
| --- | --- | --- |
| Phenotype | Correlation | Correlation |
| Age | 0.3 | 0.28 |
| Systolic Blood Pressure | 0.15 | 0.18 |
| Pulse | 0.09 | 0.02 |
| Glucose | 0.08 | 0.06 |
| Cystatin C | 0.12 | 0.09 |
| HDL | -0.06 | -0.01 |
| Phosphate | 0.17 | 0.05 |
| Triglycerides | 0.1 | 0.04 |

**Table S4:** Association of predicted AAC with biomarkers Univariate regression analysis of risk factors at baseline for predicted AAC after adjusting for age and sex in model 1 and after adjusting for socioeconomic factors, BMI, and smoking status in addition to adjusting for age and sex in model 2. The estimate of the effect size and the standard error of the estimated effect size are given along with p-values for each coefficient in a univariate fit.

|  | Model 1 |  | Model 2 |  |
| --- | --- | --- | --- | --- |
| Biomarker | Effect size | P-value | Effect size | P-value |
| HDL cholesterol | -0.041 (-0.05) | 2.46E-09 | -0.061 (-0.07) | 1.33E-19 |
| Creatinine | -0.040 (-0.05) | 3.03E-07 | -0.026 (-0.04) | 3.53E-02 |
| Apolipoprotein A | -0.031 (-0.04) | 4.77E-05 | -0.046 (-0.06) | 1.60E-11 |
| Testosterone | -0.027 (-0.06) | 1.00E+00 | -0.044 (-0.08) | 1.00E+00 |
| Urea | -0.016 (-0.03) | 1.00E+00 | -0.009 (-0.02) | 1.00E+00 |
| Total bilirubin | -0.014 (-0.02) | 1.00E+00 | -0.010 (-0.02) | 1.00E+00 |
| Rheumatoid factor | -0.005 (-0.04) | 1.00E+00 | -0.004 (-0.04) | 1.00E+00 |
| Cholesterol | -0.005 (-0.01) | 1.00E+00 | -0.006 (-0.02) | 1.00E+00 |
| Direct bilirubin | -0.005 (-0.02) | 1.00E+00 | -0.001 (-0.01) | 1.00E+00 |
| Vitamin D | -0.004 (-0.01) | 1.00E+00 | -0.009 (-0.02) | 1.00E+00 |
| IGF-1 | -0.003 (-0.01) | 1.00E+00 | 0.000 (-0.01) | 1.00E+00 |
| Oestradiol | -0.003 (-0.02) | 1.00E+00 | -0.006 (-0.03) | 1.00E+00 |
| Alkaline phosphatase | -0.000 (-0.01) | 1.00E+00 | 0.005 (-0.01) | 1.00E+00 |
| LDL direct | 0.001 (-0.01) | 1.00E+00 | 0.004 (-0.01) | 1.00E+00 |
| Urate | 0.003 (-0.01) | 1.00E+00 | 0.018 (0.01) | 1.00E+00 |
| Lipoprotein A | 0.003 (-0.01) | 1.00E+00 | 0.004 (-0.01) | 1.00E+00 |
| Aspartate aminotransferase | 0.005 (-0.01) | 1.00E+00 | 0.011 (0.00) | 1.00E+00 |
| Cystatin C | 0.007 (-0.00) | 1.00E+00 | 0.011 (-0.00) | 1.00E+00 |
| Alanine aminotransferase | 0.007 (-0.00) | 1.00E+00 | 0.019 (0.01) | 3.27E-01 |
| C-reactive protein | 0.010 (0.00) | 1.00E+00 | 0.020 (0.01) | 1.20E-01 |
| Albumin | 0.014 (0.00) | 1.00E+00 | 0.012 (0.00) | 1.00E+00 |
| SHBG | 0.016 (0.00) | 1.00E+00 | -0.001 (-0.01) | 1.00E+00 |
| Total protein | 0.019 (0.01) | 1.40E-01 | 0.028 (0.02) | 8.58E-05 |
| Gamma glutamyltransferase | 0.023 (0.01) | 1.53E-02 | 0.028 (0.02) | 2.85E-04 |
| Apolipoprotein B | 0.024 (0.01) | 1.31E-03 | 0.027 (0.02) | 8.03E-05 |
| Triglycerides | 0.027 (0.02) | 1.51E-04 | 0.036 (0.02) | 6.95E-08 |
| Calcium | 0.028 (0.02) | 3.51E-05 | 0.028 (0.02) | 4.20E-05 |
| Glucose | 0.035 (0.02) | 2.02E-08 | 0.040 (0.03) | 1.53E-11 |
| Phosphate | 0.056 (0.05) | 1.97E-22 | 0.051 (0.04) | 2.17E-18 |
| Glycated haemoglobin (HbA1c) | 0.070 (0.06) | 2.43E-39 | 0.075 (0.06) | 5.58E-43 |

**Table S5:** Association of predicted AAC with physiological markers. Univariate regression analysis of risk factors at baseline for predicted AAC after adjusting for age and sex in model 1 and after adjusting for socioeconomic factors, BMI, and smoking status in addition to adjusting for age and sex in model 2. The estimate of the effect size and the standard error of the estimated effect size are given along with p-values for each coefficient in a univariate fit.

| Physiological Measure | Model 1 |  | Model 2 |  |
| --- | --- | --- | --- | --- |
|  | Effect size | P-value | Effect size | P-value |
| Heel bone mineral density (BMD) T-score, manual entry (right) | -0.158 (-0.35) | 1.00E+00 | -0.131 (-0.33) | 1.00E+00 |
| Heel bone mineral density (BMD) T-score, manual entry (left) | -0.143 (-0.33) | 1.00E+00 | -0.086 (-0.28) | 1.00E+00 |
| Amount of tobacco currently smoked | -0.112 (-0.23) | 1.00E+00 | -0.101 (-0.22) | 1.00E+00 |
| Whole body fat-free mass | -0.105 (-0.12) | 1.05E-26 | -0.115 (-0.14) | 3.10E-19 |
| Leg predicted mass (right) | -0.105 (-0.12) | 1.61E-29 | -0.121 (-0.14) | 7.60E-21 |
| Whole body water mass | -0.104 (-0.12) | 1.26E-26 | -0.114 (-0.14) | 3.71E-19 |
| Leg fat-free mass (right) | -0.103 (-0.12) | 1.45E-29 | -0.119 (-0.14) | 7.25E-21 |
| Forced expiratory volume in 1-second (FEV1), predicted | -0.100 (-0.14) | 3.18E-03 | -0.095 (-0.14) | 5.83E-03 |
| Arm predicted mass (right) | -0.100 (-0.12) | 2.38E-22 | -0.108 (-0.13) | 1.54E-15 |
| Leg predicted mass (left) | -0.098 (-0.12) | 9.96E-27 | -0.119 (-0.14) | 6.99E-19 |
| Arm fat-free mass (right) | -0.098 (-0.12) | 3.98E-22 | -0.106 (-0.13) | 2.98E-15 |
| Trunk predicted mass | -0.098 (-0.12) | 2.53E-22 | -0.095 (-0.12) | 2.08E-15 |
| Trunk fat-free mass | -0.097 (-0.12) | 9.34E-23 | -0.097 (-0.12) | 2.69E-16 |
| Leg fat-free mass (left) | -0.096 (-0.11) | 1.79E-26 | -0.116 (-0.14) | 1.29E-18 |
| Basal metabolic rate | -0.090 (-0.11) | 8.99E-25 | -0.108 (-0.13) | 9.76E-18 |
| Arm fat-free mass (left) | -0.088 (-0.11) | 1.52E-19 | -0.094 (-0.12) | 8.52E-13 |
| Number of cigarettes previously smoked daily (current cigar/pipe smokers) | -0.084 (-0.32) | 1.00E+00 | -0.099 (-0.36) | 1.00E+00 |
| Arm predicted mass (left) | -0.081 (-0.10) | 9.48E-18 | -0.081 (-0.10) | 1.05E-10 |
| Forced expiratory volume in 1-second (FEV1), Best measure | -0.077 (-0.09) | 2.28E-20 | -0.073 (-0.09) | 2.19E-18 |
| Heel quantitative ultrasound index (QUI), manual entry (left) | -0.072 (-0.27) | 1.00E+00 | -0.080 (-0.28) | 1.00E+00 |
| Maximum workload during fitness test | -0.071 (-0.10) | 5.42E-04 | -0.074 (-0.10) | 1.00E+00 |
| Heel bone mineral density (BMD), manual entry (left) | -0.070 (-0.26) | 1.00E+00 | -0.077 (-0.27) | 1.00E+00 |
| Heel broadband ultrasound attenuation (left) | -0.064 (-0.08) | 8.04E-11 | -0.058 (-0.08) | 3.54E-08 |
| Weight (pre-imaging) | -0.064 (-0.08) | 1.34E-15 | -0.094 (-0.12) | 3.45E-12 |
| Forced expiratory volume in 1-second (FEV1), predicted percentage | -0.064 (-0.08) | 1.43E-10 | -0.038 (-0.05) | 6.32E-03 |
| Heel bone ultrasound T-score, manual entry | -0.063 (-0.09) | 1.12E-02 | -0.052 (-0.08) | 2.21E-01 |
| Heel bone mineral density (BMD) (left) | -0.062 (-0.08) | 2.09E-10 | -0.055 (-0.07) | 8.32E-08 |
| Heel quantitative ultrasound index (QUI), direct entry (left) | -0.062 (-0.08) | 2.04E-10 | -0.055 (-0.07) | 8.51E-08 |
| Heel bone mineral density (BMD) T-score, automated (left) | -0.062 (-0.08) | 2.05E-10 | -0.055 (-0.07) | 8.53E-08 |
| Forced expiratory volume in 1-second (FEV1) | -0.062 (-0.08) | 4.50E-17 | -0.064 (-0.08) | 2.21E-17 |
| Heel broadband ultrasound attenuation (right) | -0.060 (-0.08) | 3.67E-09 | -0.051 (-0.07) | 4.91E-06 |
| Forced vital capacity (FVC), Best measure | -0.060 (-0.08) | 1.98E-11 | -0.069 (-0.08) | 1.01E-14 |
| Heel bone mineral density (BMD) T-score, automated (right) | -0.056 (-0.07) | 3.23E-08 | -0.048 (-0.06) | 1.20E-05 |
| Heel quantitative ultrasound index (QUI), direct entry (right) | -0.056 (-0.07) | 3.23E-08 | -0.048 (-0.06) | 1.20E-05 |
| Heel bone mineral density (BMD) (right) | -0.056 (-0.07) | 3.49E-08 | -0.049 (-0.07) | 9.31E-06 |
| Weight | -0.053 (-0.06) | 1.13E-17 | -0.103 (-0.13) | 7.32E-14 |
| Standing height | -0.052 (-0.07) | 9.29E-11 | -0.058 (-0.07) | 4.23E-13 |
| Height | -0.051 (-0.07) | 2.01E-05 | -0.055 (-0.07) | 2.55E-06 |
| Weight, manual entry | -0.047 (-0.15) | 1.00E+00 | -0.060 (-0.32) | 1.00E+00 |
| Forced vital capacity (FVC) | -0.047 (-0.06) | 8.27E-09 | -0.058 (-0.07) | 1.34E-13 |
| Non-cancer illness year/age first occurred | -0.044 (-0.06) | 9.60E-10 | -0.048 (-0.06) | 4.59E-12 |
| Heel broadband ultrasound attenuation (BUA), manual entry (left) | -0.044 (-0.25) | 1.00E+00 | -0.039 (-0.25) | 1.00E+00 |
| Number of trend entries | -0.043 (-0.07) | 3.54E-01 | -0.046 (-0.07) | 1.74E-01 |
| Father's age at death | -0.042 (-0.05) | 2.94E-08 | -0.040 (-0.05) | 1.79E-07 |
| Ankle spacing width | -0.041 (-0.06) | 1.20E-04 | -0.034 (-0.05) | 1.44E-01 |
| Heel Broadband ultrasound attenuation, direct entry | -0.040 (-0.05) | 3.65E-06 | -0.030 (-0.04) | 7.92E-03 |
| Heel bone mineral density (BMD) | -0.040 (-0.05) | 2.10E-06 | -0.031 (-0.04) | 3.85E-03 |
| Heel bone mineral density (BMD) T-score, automated | -0.040 (-0.05) | 3.00E-06 | -0.030 (-0.04) | 5.13E-03 |
| Heel quantitative ultrasound index (QUI), direct entry | -0.040 (-0.05) | 3.00E-06 | -0.030 (-0.04) | 5.13E-03 |
| Heel quantitative ultrasound index (QUI), manual entry | -0.039 (-0.07) | 1.00E+00 | -0.029 (-0.06) | 1.00E+00 |
| Hip circumference | -0.038 (-0.05) | 2.93E-12 | -0.037 (-0.05) | 2.04E-02 |
| Heel bone mineral density (BMD), manual entry | -0.038 (-0.07) | 1.00E+00 | -0.027 (-0.06) | 1.00E+00 |
| Hand grip strength (left) | -0.037 (-0.05) | 3.17E-04 | -0.038 (-0.05) | 1.79E-04 |
| Sitting height | -0.035 (-0.05) | 1.73E-05 | -0.033 (-0.05) | 1.59E-04 |
| Leg fat mass (left) | -0.035 (-0.05) | 4.75E-06 | -0.004 (-0.04) | 1.00E+00 |
| Arm fat mass (left) | -0.035 (-0.04) | 3.03E-09 | -0.030 (-0.06) | 1.00E+00 |
| Number of symbol digit matches attempted | -0.035 (-0.05) | 2.07E-03 | -0.030 (-0.05) | 2.97E-02 |
| Arm fat mass (right) | -0.034 (-0.04) | 6.29E-09 | -0.029 (-0.06) | 1.00E+00 |
| Number of symbol digit matches made correctly | -0.034 (-0.05) | 2.22E-03 | -0.031 (-0.05) | 1.76E-02 |
| Ankle spacing width (right) | -0.034 (-0.05) | 2.11E-01 | -0.035 (-0.06) | 1.00E+00 |
| Errors before selecting correct item in alphanumeric path (trail #2) | -0.034 (-0.09) | 1.00E+00 | -0.036 (-0.09) | 1.00E+00 |
| Leg fat mass (right) | -0.033 (-0.05) | 2.61E-05 | 0.007 (-0.03) | 1.00E+00 |
| Hand grip strength (right) | -0.033 (-0.05) | 3.34E-03 | -0.033 (-0.05) | 4.33E-03 |
| Peak expiratory flow (PEF) | -0.032 (-0.04) | 9.61E-05 | -0.028 (-0.04) | 3.38E-03 |
| Genetic relatedness pairing | -0.031 (-0.08) | 1.00E+00 | -0.026 (-0.08) | 1.00E+00 |
| Whole body fat mass | -0.031 (-0.04) | 8.44E-07 | -0.013 (-0.04) | 1.00E+00 |
| Ankle spacing width (left) | -0.030 (-0.05) | 1.00E+00 | -0.030 (-0.05) | 1.00E+00 |
| Trunk fat mass | -0.029 (-0.04) | 1.87E-06 | -0.010 (-0.03) | 1.00E+00 |
| Duration of fitness test | -0.028 (-0.05) | 1.00E+00 | -0.031 (-0.05) | 1.00E+00 |
| Arm fat percentage (left) | -0.028 (-0.04) | 1.69E-02 | 0.049 (0.02) | 2.72E-01 |
| Years of bringing up phlegm/sputum/mucus on most days | -0.025 (-0.08) | 1.00E+00 | -0.012 (-0.06) | 1.00E+00 |
| Waist circumference | -0.025 (-0.04) | 6.75E-03 | 0.008 (-0.01) | 1.00E+00 |
| Arm fat percentage (right) | -0.024 (-0.04) | 1.76E-01 | 0.060 (0.03) | 7.38E-03 |
| Pulse wave reflection index | -0.021 (-0.04) | 1.00E+00 | -0.022 (-0.04) | 1.00E+00 |
| Cascot confidence score | -0.021 (-0.06) | 1.00E+00 | -0.017 (-0.06) | 1.00E+00 |
| Fluid intelligence score.1 | -0.020 (-0.03) | 1.00E+00 | -0.018 (-0.03) | 1.00E+00 |
| Fluid intelligence score | -0.020 (-0.04) | 1.00E+00 | -0.018 (-0.03) | 1.00E+00 |
| Heel Broadband ultrasound attenuation (BUA), manual entry | -0.019 (-0.05) | 1.00E+00 | -0.009 (-0.04) | 1.00E+00 |
| Thursday average acceleration | -0.019 (-0.03) | 1.00E+00 | -0.024 (-0.04) | 6.50E-01 |
| Average acceleration 12:00 - 12:59 | -0.019 (-0.03) | 1.00E+00 | -0.021 (-0.04) | 1.00E+00 |
| QRS duration | -0.019 (-0.03) | 1.00E+00 | -0.015 (-0.03) | 1.00E+00 |
| Errors before selecting correct item in numeric path (trail 1) | -0.018 (-0.11) | 1.00E+00 | -0.010 (-0.10) | 1.00E+00 |
| Average acceleration 23:00 - 23:59 | -0.018 (-0.03) | 1.00E+00 | -0.019 (-0.03) | 1.00E+00 |
| Body fat percentage | -0.017 (-0.03) | 1.00E+00 | 0.057 (0.03) | 1.05E-03 |
| Friday average acceleration | -0.016 (-0.03) | 1.00E+00 | -0.020 (-0.03) | 1.00E+00 |
| Wednesday average acceleration | -0.016 (-0.03) | 1.00E+00 | -0.019 (-0.03) | 1.00E+00 |
| Overall acceleration average | -0.016 (-0.03) | 1.00E+00 | -0.020 (-0.04) | 1.00E+00 |
| Average acceleration 22:00 - 22:59 | -0.016 (-0.03) | 1.00E+00 | -0.016 (-0.03) | 1.00E+00 |
| Mother's age at death | -0.016 (-0.03) | 1.00E+00 | -0.017 (-0.03) | 1.00E+00 |
| Average acceleration 10:00 - 10:59 | -0.015 (-0.03) | 1.00E+00 | -0.018 (-0.03) | 1.00E+00 |
| Average acceleration 17:00 - 17:59 | -0.015 (-0.03) | 1.00E+00 | -0.018 (-0.03) | 1.00E+00 |
| Sunday average acceleration | -0.015 (-0.03) | 1.00E+00 | -0.018 (-0.03) | 1.00E+00 |
| Leg fat percentage (left) | -0.015 (-0.03) | 1.00E+00 | 0.119 (0.08) | 2.13E-07 |
| ECG, number of stages in a phase | -0.014 (-0.04) | 1.00E+00 | -0.014 (-0.04) | 1.00E+00 |
| No-wear time bias adjusted average acceleration | -0.014 (-0.03) | 1.00E+00 | -0.019 (-0.03) | 1.00E+00 |
| Saturday average acceleration | -0.014 (-0.03) | 1.00E+00 | -0.015 (-0.03) | 1.00E+00 |
| Trunk fat percentage | -0.014 (-0.02) | 1.00E+00 | 0.029 (0.01) | 3.20E-01 |
| Average acceleration 14:00 - 14:59 | -0.014 (-0.03) | 1.00E+00 | -0.016 (-0.03) | 1.00E+00 |
| Tuesday average acceleration | -0.013 (-0.03) | 1.00E+00 | -0.016 (-0.03) | 1.00E+00 |
| Average acceleration 11:00 - 11:59 | -0.013 (-0.03) | 1.00E+00 | -0.016 (-0.03) | 1.00E+00 |
| Monday average acceleration | -0.013 (-0.03) | 1.00E+00 | -0.016 (-0.03) | 1.00E+00 |
| Genetic principal components | -0.012 (-0.02) | 1.00E+00 | 0.012 (-0.02) | 1.00E+00 |
| Pulse wave peak to peak time | -0.012 (-0.03) | 1.00E+00 | -0.009 (-0.03) | 1.00E+00 |
| Average acceleration 16:00 - 16:59 | -0.012 (-0.03) | 1.00E+00 | -0.013 (-0.03) | 1.00E+00 |
| Maximum digits remembered correctly.1 | -0.012 (-0.03) | 1.00E+00 | -0.013 (-0.03) | 1.00E+00 |
| Average acceleration 15:00 - 15:59 | -0.012 (-0.03) | 1.00E+00 | -0.014 (-0.03) | 1.00E+00 |
| Total errors traversing numeric path (trail #1) | -0.012 (-0.04) | 1.00E+00 | -0.011 (-0.04) | 1.00E+00 |
| Number of correct matches in round.1 | -0.012 (-0.03) | 1.00E+00 | -0.010 (-0.02) | 1.00E+00 |
| Years of cough on most days | -0.011 (-0.05) | 1.00E+00 | -0.001 (-0.04) | 1.00E+00 |
| Average acceleration 09:00 - 09:59 | -0.011 (-0.03) | 1.00E+00 | -0.015 (-0.03) | 1.00E+00 |
| P duration | -0.011 (-0.02) | 1.00E+00 | -0.008 (-0.02) | 1.00E+00 |
| No-wear time bias adjusted acceleration standard deviation | -0.011 (-0.03) | 1.00E+00 | -0.014 (-0.03) | 1.00E+00 |
| Genetic relatedness IBS0 | -0.010 (-0.06) | 1.00E+00 | -0.013 (-0.07) | 1.00E+00 |
| Standard deviation of acceleration | -0.010 (-0.02) | 1.00E+00 | -0.013 (-0.03) | 1.00E+00 |
| Average acceleration 13:00 - 13:59 | -0.010 (-0.02) | 1.00E+00 | -0.013 (-0.03) | 1.00E+00 |
| Average acceleration 21:00 - 21:59 | -0.010 (-0.02) | 1.00E+00 | -0.012 (-0.03) | 1.00E+00 |
| Leg fat percentage (right) | -0.008 (-0.03) | 1.00E+00 | 0.120 (0.09) | 4.28E-09 |
| Number of times snap-button pressed | -0.007 (-0.02) | 1.00E+00 | -0.007 (-0.02) | 1.00E+00 |
| Number of fluid intelligence questions attempted within time limit | -0.007 (-0.02) | 1.00E+00 | -0.003 (-0.02) | 1.00E+00 |
| Value entered | -0.007 (-0.02) | 1.00E+00 | -0.004 (-0.02) | 1.00E+00 |
| Number of incorrect matches in round | -0.006 (-0.02) | 1.00E+00 | -0.006 (-0.02) | 1.00E+00 |
| Heel bone mineral density (BMD), manual entry (right) | -0.005 (-0.20) | 1.00E+00 | -0.052 (-0.25) | 1.00E+00 |
| Number of days/week of vigorous physical activity 10+ minutes | -0.005 (-0.01) | 1.00E+00 | -0.008 (-0.02) | 1.00E+00 |
| Number of days/week of moderate physical activity 10+ minutes | -0.005 (-0.01) | 1.00E+00 | -0.011 (-0.02) | 1.00E+00 |
| Duration of vigorous activity | -0.004 (-0.02) | 1.00E+00 | -0.006 (-0.02) | 1.00E+00 |
| Maximum heart rate during fitness test | -0.004 (-0.03) | 1.00E+00 | 0.000 (-0.02) | 1.00E+00 |
| Average acceleration 03:00 - 03:59 | -0.004 (-0.02) | 1.00E+00 | -0.006 (-0.02) | 1.00E+00 |
| Diastolic blood pressure, automated reading | -0.004 (-0.01) | 1.00E+00 | 0.009 (-0.00) | 1.00E+00 |
| Average acceleration 00:00 - 00:59 | -0.004 (-0.02) | 1.00E+00 | -0.007 (-0.02) | 1.00E+00 |
| Genetic relatedness factor | -0.004 (-0.06) | 1.00E+00 | -0.001 (-0.05) | 1.00E+00 |
| Average acceleration 20:00 - 20:59 | -0.004 (-0.02) | 1.00E+00 | -0.005 (-0.02) | 1.00E+00 |
| Missingness | -0.003 (-0.01) | 1.00E+00 | -0.003 (-0.01) | 1.00E+00 |
| Average acceleration 18:00 - 18:59 | -0.003 (-0.02) | 1.00E+00 | -0.004 (-0.02) | 1.00E+00 |
| Maximum digits remembered correctly | -0.003 (-0.03) | 1.00E+00 | -0.003 (-0.03) | 1.00E+00 |
| Average acceleration 07:00 - 07:59 | -0.003 (-0.02) | 1.00E+00 | -0.000 (-0.01) | 1.00E+00 |
| Average Y chromosome intensities for determining sex | -0.003 (-0.03) | 1.00E+00 | -0.005 (-0.03) | 1.00E+00 |
| Time to complete round | -0.003 (-0.01) | 1.00E+00 | -0.002 (-0.01) | 1.00E+00 |
| Duration to first press of snap-button in each round | -0.002 (-0.02) | 1.00E+00 | -0.002 (-0.02) | 1.00E+00 |
| Average acceleration 01:00 - 01:59 | -0.002 (-0.02) | 1.00E+00 | -0.006 (-0.02) | 1.00E+00 |
| Average acceleration 08:00 - 08:59 | -0.002 (-0.02) | 1.00E+00 | -0.004 (-0.02) | 1.00E+00 |
| No-wear time bias adjusted acceleration minimum | -0.002 (-0.02) | 1.00E+00 | -0.003 (-0.02) | 1.00E+00 |
| Time to answer | -0.001 (-0.02) | 1.00E+00 | -0.003 (-0.02) | 1.00E+00 |
| Average acceleration 19:00 - 19:59 | -0.001 (-0.02) | 1.00E+00 | -0.001 (-0.02) | 1.00E+00 |
| Average acceleration 02:00 - 02:59 | -0.001 (-0.01) | 1.00E+00 | -0.005 (-0.02) | 1.00E+00 |
| No-wear time bias adjusted acceleration median | -0.001 (-0.02) | 1.00E+00 | -0.002 (-0.02) | 1.00E+00 |
| Number of days/week walked 10+ minutes | -0.000 (-0.01) | 1.00E+00 | -0.006 (-0.02) | 1.00E+00 |
| Average acceleration 04:00 - 04:59 | -0.000 (-0.01) | 1.00E+00 | -0.001 (-0.02) | 1.00E+00 |
| Birth weight | -0.000 (-0.01) | 1.00E+00 | -0.004 (-0.02) | 1.00E+00 |
| No-wear time bias adjusted acceleration maximum | 0.000 (-0.01) | 1.00E+00 | -0.001 (-0.02) | 1.00E+00 |
| Number of correct matches in round | 0.001 (-0.01) | 1.00E+00 | 0.001 (-0.01) | 1.00E+00 |
| Average X chromosome intensities for determining sex | 0.002 (-0.02) | 1.00E+00 | -0.001 (-0.02) | 1.00E+00 |
| Time to complete round.1 | 0.002 (-0.01) | 1.00E+00 | 0.000 (-0.01) | 1.00E+00 |
| Average acceleration 06:00 - 06:59 | 0.002 (-0.01) | 1.00E+00 | 0.002 (-0.01) | 1.00E+00 |
| Interval between previous point and current one in alphanumeric path (trail #2) | 0.003 (-0.01) | 1.00E+00 | 0.002 (-0.01) | 1.00E+00 |
| Heterozygosity, PCA corrected | 0.003 (-0.01) | 1.00E+00 | 0.004 (-0.01) | 1.00E+00 |
| Number of incorrect matches in round.1 | 0.003 (-0.01) | 1.00E+00 | 0.001 (-0.01) | 1.00E+00 |
| Particulate matter air pollution (pm10); 2010 | 0.003 (-0.01) | 1.00E+00 | -0.002 (-0.01) | 1.00E+00 |
| Mean time to correctly identify matches | 0.003 (-0.01) | 1.00E+00 | 0.003 (-0.01) | 1.00E+00 |
| Duration to complete numeric path (trail #1) | 0.004 (-0.01) | 1.00E+00 | 0.002 (-0.01) | 1.00E+00 |
| Maximum carotid IMT (intima-medial thickness) at 150 degrees | 0.004 (-0.04) | 1.00E+00 | 0.002 (-0.04) | 1.00E+00 |
| Particulate matter air pollution 2.5-10um; 2010 | 0.004 (-0.01) | 1.00E+00 | 0.002 (-0.01) | 3.23E+02 |
| Interval between previous point and current one in numeric path (trail #1) | 0.004 (-0.01) | 1.00E+00 | 0.005 (-0.01) | 1.00E+00 |
| Total errors traversing alphanumeric path (trail #2) | 0.004 (-0.02) | 1.00E+00 | 0.003 (-0.02) | 1.00E+00 |
| Diastolic blood pressure, manual reading | 0.005 (-0.03) | 1.00E+00 | 0.017 (-0.02) | 1.00E+00 |
| Duration of moderate activity | 0.005 (-0.01) | 1.00E+00 | -0.001 (-0.01) | 1.00E+00 |
| Signal-to-noise-ratio (SNR) of triplet (left) | 0.006 (-0.01) | 1.00E+00 | 0.007 (-0.01) | 1.00E+00 |
| Signal-to-noise-ratio (SNR) of triplet (right) | 0.006 (-0.01) | 1.00E+00 | 0.007 (-0.01) | 1.00E+00 |
| ECG, phase time | 0.006 (-0.02) | 1.00E+00 | 0.004 (-0.02) | 1.00E+00 |
| Average acceleration 05:00 - 05:59 | 0.006 (-0.01) | 1.00E+00 | 0.005 (-0.01) | 1.00E+00 |
| logMAR, final (right) | 0.007 (-0.01) | 1.00E+00 | 0.009 (-0.01) | 1.00E+00 |
| Heterozygosity | 0.007 (-0.00) | 1.00E+00 | -0.002 (-0.02) | 1.00E+00 |
| Number of full sisters | 0.008 (-0.00) | 1.00E+00 | 0.003 (-0.01) | 1.00E+00 |
| Nitrogen dioxide air pollution; 2010 | 0.008 (-0.00) | 1.00E+00 | -0.008 (-0.02) | 1.00E+00 |
| Interpolated Year when non-cancer illness first diagnosed | 0.009 (-0.00) | 1.00E+00 | 0.008 (-0.00) | 1.00E+00 |
| Particulate matter air pollution (pm2.5) absorbance; 2010 | 0.009 (-0.00) | 1.00E+00 | -0.002 (-0.01) | 1.00E+00 |
| Interpolated Year when cancer first diagnosed | 0.009 (-0.03) | 1.00E+00 | 0.018 (-0.02) | 1.00E+00 |
| Nitrogen oxides air pollution; 2010 | 0.009 (-0.00) | 1.00E+00 | -0.005 (-0.02) | 1.00E+00 |
| Time spent driving | 0.010 (-0.00) | 1.00E+00 | 0.010 (-0.00) | 1.00E+00 |
| Sleep duration | 0.010 (0.00) | 1.00E+00 | 0.009 (-0.00) | 1.00E+00 |
| Particulate matter air pollution (pm2.5); 2010 | 0.010 (0.00) | 1.00E+00 | -0.003 (-0.01) | 1.00E+00 |
| Number of attempts | 0.010 (-0.01) | 1.00E+00 | 0.010 (-0.01) | 1.00E+00 |
| Time to complete test | 0.011 (-0.02) | 1.00E+00 | 0.008 (-0.02) | 1.00E+00 |
| Duration of walks | 0.011 (0.00) | 1.00E+00 | 0.006 (-0.00) | 1.00E+00 |
| Duration to entering value | 0.011 (-0.00) | 1.00E+00 | 0.010 (-0.00) | 1.00E+00 |
| Number of self-reported cancers | 0.012 (0.00) | 1.00E+00 | 0.010 (0.00) | 1.00E+00 |
| Pulse rate | 0.013 (-0.00) | 1.00E+00 | 0.018 (0.00) | 1.00E+00 |
| Number of triplets attempted (right) | 0.013 (-0.00) | 1.00E+00 | 0.016 (-0.00) | 1.00E+00 |
| Number of triplets attempted (left) | 0.014 (-0.00) | 1.00E+00 | 0.016 (-0.00) | 1.00E+00 |
| Number of automated diagnostic comments recorded during 12-lead ECG | 0.014 (0.00) | 1.00E+00 | 0.012 (-0.00) | 1.00E+00 |
| Number of full brothers | 0.014 (0.00) | 1.00E+00 | 0.009 (-0.00) | 1.00E+00 |
| Speech-reception-threshold (SRT) estimate (left) | 0.014 (-0.00) | 1.00E+00 | 0.013 (-0.00) | 1.00E+00 |
| Minimum carotid IMT (intima-medial thickness) at 150 degrees | 0.014 (-0.03) | 1.00E+00 | 0.011 (-0.03) | 1.00E+00 |
| Time spent using computer | 0.015 (0.01) | 1.00E+00 | 0.013 (0.00) | 1.00E+00 |
| Pulse wave Arterial Stiffness index | 0.016 (-0.00) | 1.00E+00 | 0.013 (-0.00) | 1.00E+00 |
| Number of operations, self-reported | 0.016 (0.01) | 6.01E-01 | 0.016 (0.01) | 8.55E-01 |
| Speech-reception-threshold (SRT) estimate (right) | 0.017 (0.00) | 1.00E+00 | 0.015 (-0.00) | 1.00E+00 |
| Duration screen displayed | 0.017 (-0.02) | 1.00E+00 | 0.018 (-0.02) | 1.00E+00 |
| Pulse rate, automated reading | 0.018 (0.01) | 1.64E-01 | 0.022 (0.01) | 8.92E-03 |
| Number of older siblings | 0.018 (0.00) | 1.00E+00 | 0.010 (-0.01) | 1.00E+00 |
| Pulse rate (during blood-pressure measurement) | 0.019 (-0.02) | 1.00E+00 | 0.019 (-0.02) | 1.00E+00 |
| Cancer year/age first occurred | 0.020 (-0.02) | 1.00E+00 | 0.021 (-0.02) | 1.00E+00 |
| ECG, phase duration | 0.022 (-0.00) | 1.00E+00 | 0.023 (-0.00) | 1.00E+00 |
| Ventricular rate | 0.022 (0.01) | 5.10E-01 | 0.024 (0.01) | 2.32E-01 |
| logMAR, final (left) | 0.022 (0.00) | 1.00E+00 | 0.020 (0.00) | 1.00E+00 |
| Time spent watching television (TV) | 0.024 (0.01) | 3.74E-04 | 0.025 (0.02) | 1.37E-04 |
| Duration to complete alphanumeric path (trail #2) | 0.025 (0.01) | 4.27E-01 | 0.022 (0.01) | 1.00E+00 |
| Heel quantitative ultrasound index (QUI), manual entry (right) | 0.025 (-0.17) | 1.00E+00 | -0.012 (-0.21) | 1.00E+00 |
| Mean carotid IMT (intima-medial thickness) at 150 degrees | 0.026 (-0.01) | 1.00E+00 | 0.023 (-0.02) | 1.00E+00 |
| Reported occurrences of cancer | 0.030 (-0.00) | 1.00E+00 | 0.029 (-0.00) | 1.00E+00 |
| Minimum carotid IMT (intima-medial thickness) at 120 degrees | 0.035 (-0.00) | 1.00E+00 | 0.029 (-0.01) | 1.00E+00 |
| Minimum carotid IMT (intima-medial thickness) at 240 degrees | 0.037 (-0.00) | 1.00E+00 | 0.025 (-0.01) | 1.00E+00 |
| Impedance of arm (left) | 0.039 (0.02) | 7.07E-05 | 0.027 (0.01) | 8.39E-01 |
| Spells in hospital | 0.041 (0.03) | 5.97E-08 | 0.038 (0.03) | 9.54E-07 |
| Maximum carotid IMT (intima-medial thickness) at 120 degrees | 0.043 (0.00) | 1.00E+00 | 0.038 (-0.00) | 1.00E+00 |
| Impedance of arm (right) | 0.045 (0.03) | 4.83E-07 | 0.035 (0.02) | 2.59E-02 |
| ECG, heart rate | 0.048 (0.03) | 2.39E-02 | 0.053 (0.03) | 5.20E-03 |
| Mean carotid IMT (intima-medial thickness) at 120 degrees | 0.048 (0.01) | 1.00E+00 | 0.043 (0.00) | 1.00E+00 |
| Impedance of whole body | 0.052 (0.04) | 1.05E-11 | 0.044 (0.03) | 1.06E-04 |
| Impedance of leg (left) | 0.053 (0.04) | 3.56E-19 | 0.045 (0.03) | 8.48E-09 |
| Mean carotid IMT (intima-medial thickness) at 240 degrees | 0.054 (0.01) | 1.00E+00 | 0.040 (-0.00) | 1.00E+00 |
| Number of self-reported non-cancer illnesses | 0.054 (0.04) | 1.77E-25 | 0.058 (0.05) | 9.44E-29 |
| Impedance of leg (right) | 0.057 (0.05) | 4.75E-22 | 0.050 (0.04) | 5.29E-11 |
| Heel broadband ultrasound attenuation (BUA), manual entry (right) | 0.058 (-0.15) | 1.00E+00 | 0.020 (-0.19) | 1.00E+00 |
| Systolic blood pressure, manual reading | 0.059 (0.02) | 1.00E+00 | 0.070 (0.03) | 1.98E-01 |
| Medication for cholesterol, blood pressure, diabetes, or take exogenous hormones | 0.060 (0.05) | 1.74E-16 | 0.060 (0.05) | 8.81E-17 |
| Pulse wave velocity (manual entry) | 0.061 (-0.71) | 1.00E+00 | 0.019 (-0.50) | 1.00E+00 |
| Maximum carotid IMT (intima-medial thickness) at 240 degrees | 0.062 (0.02) | 1.00E+00 | 0.048 (0.01) | 1.00E+00 |
| Systolic blood pressure, automated reading | 0.062 (0.05) | 6.29E-28 | 0.074 (0.06) | 5.77E-39 |
| Maximum carotid IMT (intima-medial thickness) at 210 degrees | 0.074 (0.03) | 1.10E-01 | 0.061 (0.02) | 1.00E+00 |
| Number of treatments/medications taken | 0.080 (0.07) | 6.66E-54 | 0.084 (0.07) | 6.17E-59 |
| Minimum carotid IMT (intima-medial thickness) at 210 degrees | 0.085 (0.05) | 1.17E-02 | 0.073 (0.03) | 1.24E-01 |
| Mean carotid IMT (intima-medial thickness) at 210 degrees | 0.085 (0.05) | 1.43E-02 | 0.072 (0.03) | 1.84E-01 |

**Table S6:** Association of predicted AAC with complete blood count markers. Univariate regression analysis of risk factors at baseline for predicted AAC after adjusting for age and sex in model 1 and after adjusting for socioeconomic factors, BMI, and smoking status in addition to adjusting for age and sex in model 2. The estimate of the effect size and the standard error of the estimated effect size are given along with p-values for each coefficient in a univariate fit.

| CBC Measure | Model 1 |  | Model 2 |  |
| --- | --- | --- | --- | --- |
|  | Effect size | P-value | Effect size | P-value |
| Red blood cell (erythrocyte) count | -0.025 (-0.04) | 2.07E-02 | -0.010 (-0.02) | 1.00E+00 |
| Lymphocyte percentage | -0.015 (-0.03) | 1.00E+00 | -0.014 (-0.02) | 1.00E+00 |
| Haemoglobin concentration | -0.011 (-0.02) | 1.00E+00 | -0.008 (-0.02) | 1.00E+00 |
| Platelet distribution width | -0.009 (-0.02) | 1.00E+00 | -0.006 (-0.02) | 1.00E+00 |
| Haematocrit percentage | -0.007 (-0.02) | 1.00E+00 | -0.003 (-0.02) | 1.00E+00 |
| Mean corpuscular haemoglobin concentration | -0.006 (-0.02) | 1.00E+00 | -0.007 (-0.02) | 1.00E+00 |
| Mean platelet (thrombocyte) volume | -0.006 (-0.02) | 1.00E+00 | -0.007 (-0.02) | 1.00E+00 |
| Nucleated red blood cell percentage | -0.002 (-0.01) | 1.00E+00 | -0.003 (-0.01) | 1.00E+00 |
| Monocyte percentage | -0.001 (-0.01) | 1.00E+00 | 0.002 (-0.01) | 1.00E+00 |
| Nucleated red blood cell count | 0.000 (-0.01) | 1.00E+00 | -0.001 (-0.01) | 1.00E+00 |
| Red blood cell (erythrocyte) distribution width | 0.002 (-0.01) | 1.00E+00 | 0.004 (-0.01) | 1.00E+00 |
| Eosinophill percentage | 0.004 (-0.01) | 1.00E+00 | 0.004 (-0.01) | 1.00E+00 |
| Mean reticulocyte volume | 0.009 (-0.00) | 1.00E+00 | 0.003 (-0.01) | 1.00E+00 |
| Basophill percentage | 0.012 (0.00) | 1.00E+00 | 0.009 (-0.00) | 1.00E+00 |
| Neutrophill percentage | 0.012 (0.00) | 1.00E+00 | 0.010 (0.00) | 1.00E+00 |
| Reticulocyte count | 0.013 (0.00) | 1.00E+00 | 0.021 (0.01) | 3.12E-02 |
| Mean sphered cell volume | 0.015 (0.01) | 1.00E+00 | -0.000 (-0.01) | 1.00E+00 |
| Reticulocyte percentage | 0.016 (0.01) | 5.24E-01 | 0.023 (0.01) | 2.75E-03 |
| Mean corpuscular haemoglobin | 0.019 (0.01) | 7.10E-02 | 0.005 (-0.01) | 1.00E+00 |
| High light scatter reticulocyte count | 0.020 (0.01) | 3.23E-02 | 0.035 (0.02) | 5.42E-08 |
| Eosinophill count | 0.021 (0.01) | 2.07E-02 | 0.018 (0.01) | 1.11E-01 |
| Immature reticulocyte fraction | 0.021 (0.01) | 2.15E-02 | 0.028 (0.02) | 3.12E-05 |
| Basophill count | 0.021 (0.01) | 2.58E-02 | 0.017 (0.01) | 4.40E-01 |
| High light scatter reticulocyte percentage | 0.023 (0.01) | 1.87E-03 | 0.035 (0.02) | 2.96E-08 |
| Platelet count | 0.024 (0.01) | 1.48E-03 | 0.023 (0.01) | 4.37E-03 |
| Platelet crit | 0.025 (0.01) | 1.17E-03 | 0.023 (0.01) | 4.64E-03 |
| Mean corpuscular volume | 0.027 (0.02) | 5.82E-05 | 0.010 (0.00) | 1.00E+00 |
| Lymphocyte count | 0.027 (0.02) | 5.53E-05 | 0.024 (0.01) | 1.44E-03 |
| Monocyte count | 0.034 (0.02) | 1.54E-08 | 0.033 (0.02) | 1.25E-07 |
| Neutrophill count | 0.050 (0.04) | 4.64E-21 | 0.044 (0.03) | 1.71E-15 |
| White blood cell (leukocyte) count | 0.056 (0.05) | 1.34E-25 | 0.049 (0.04) | 3.97E-19 |

**Table S13:**
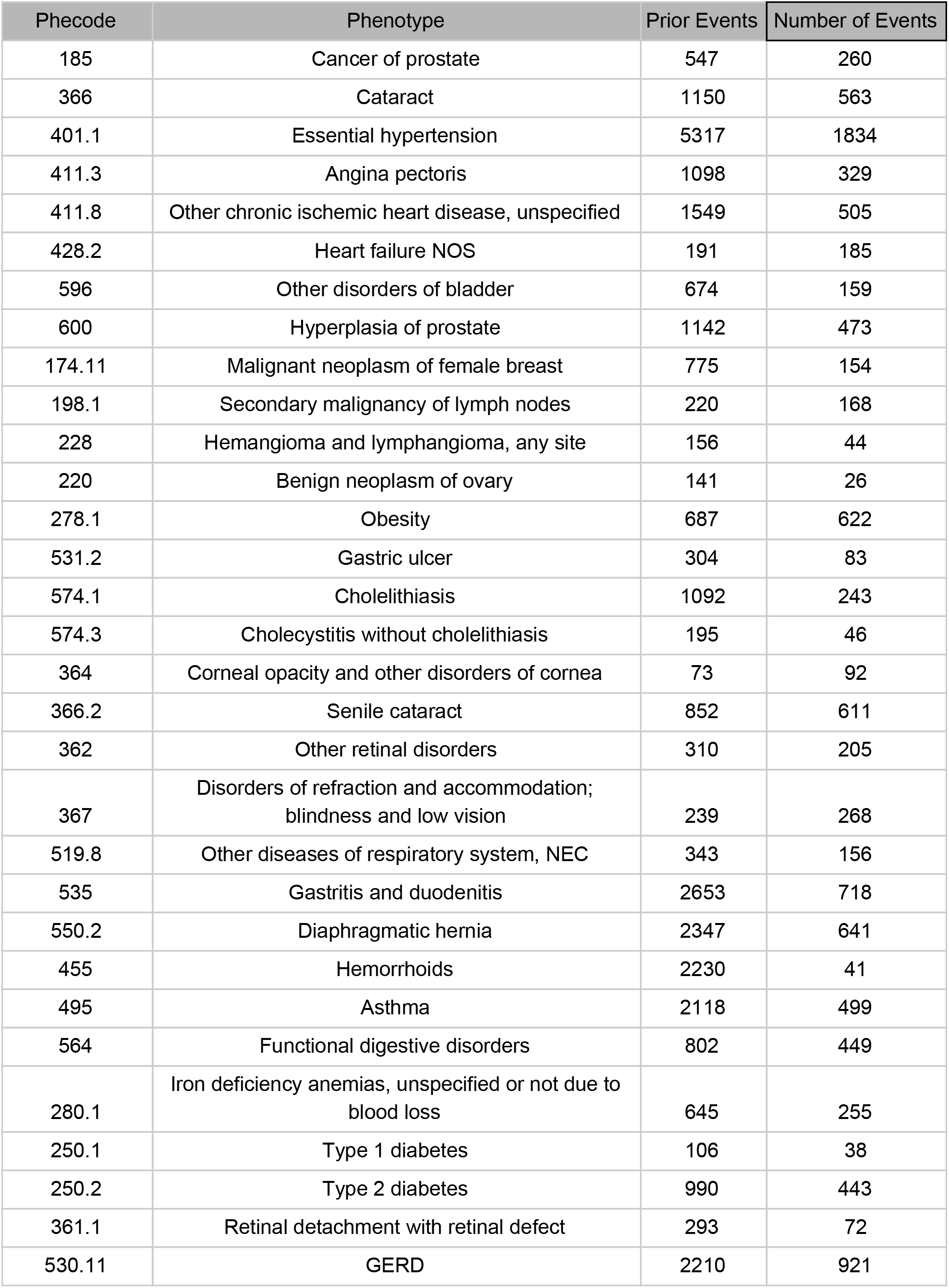
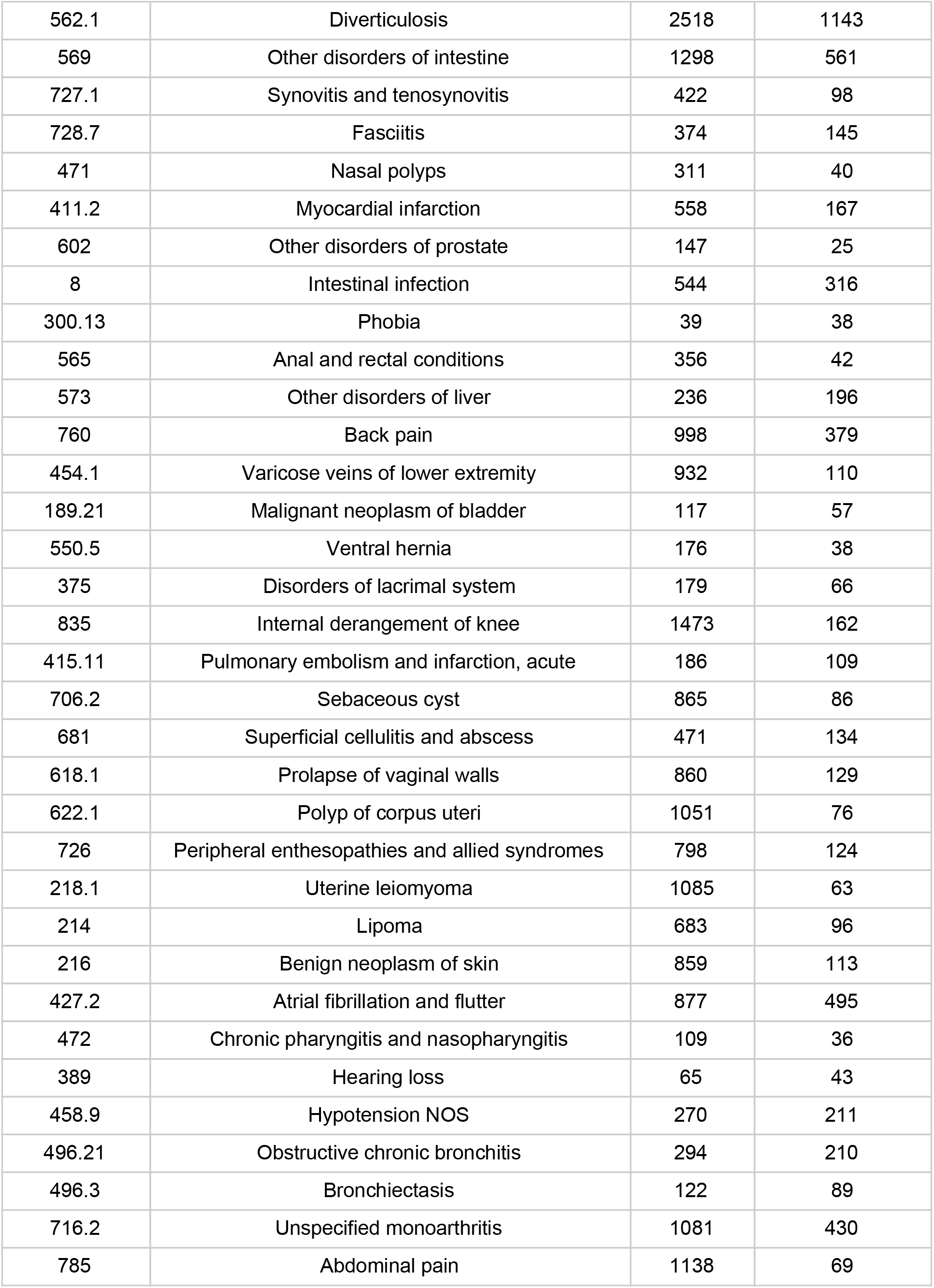
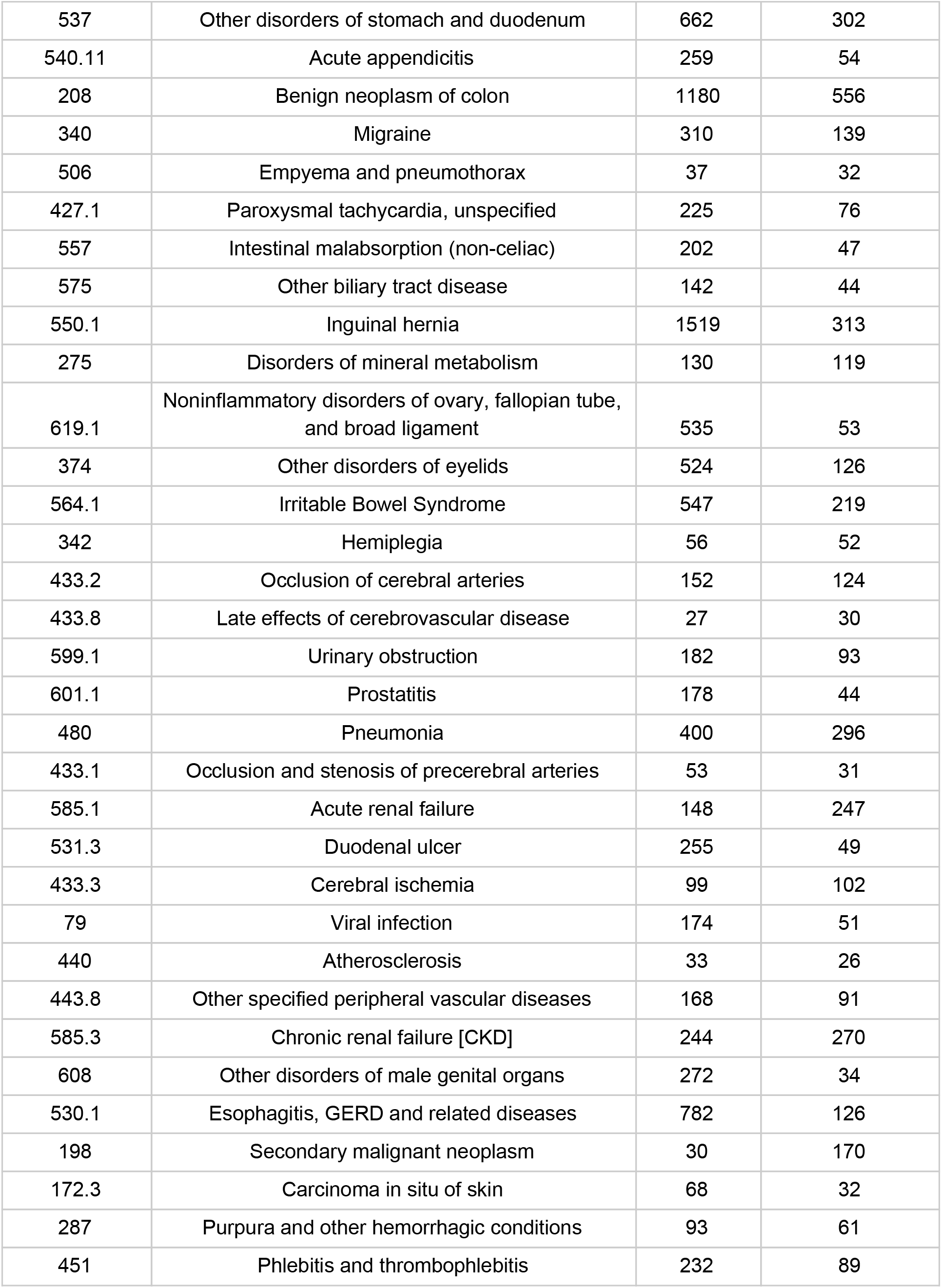
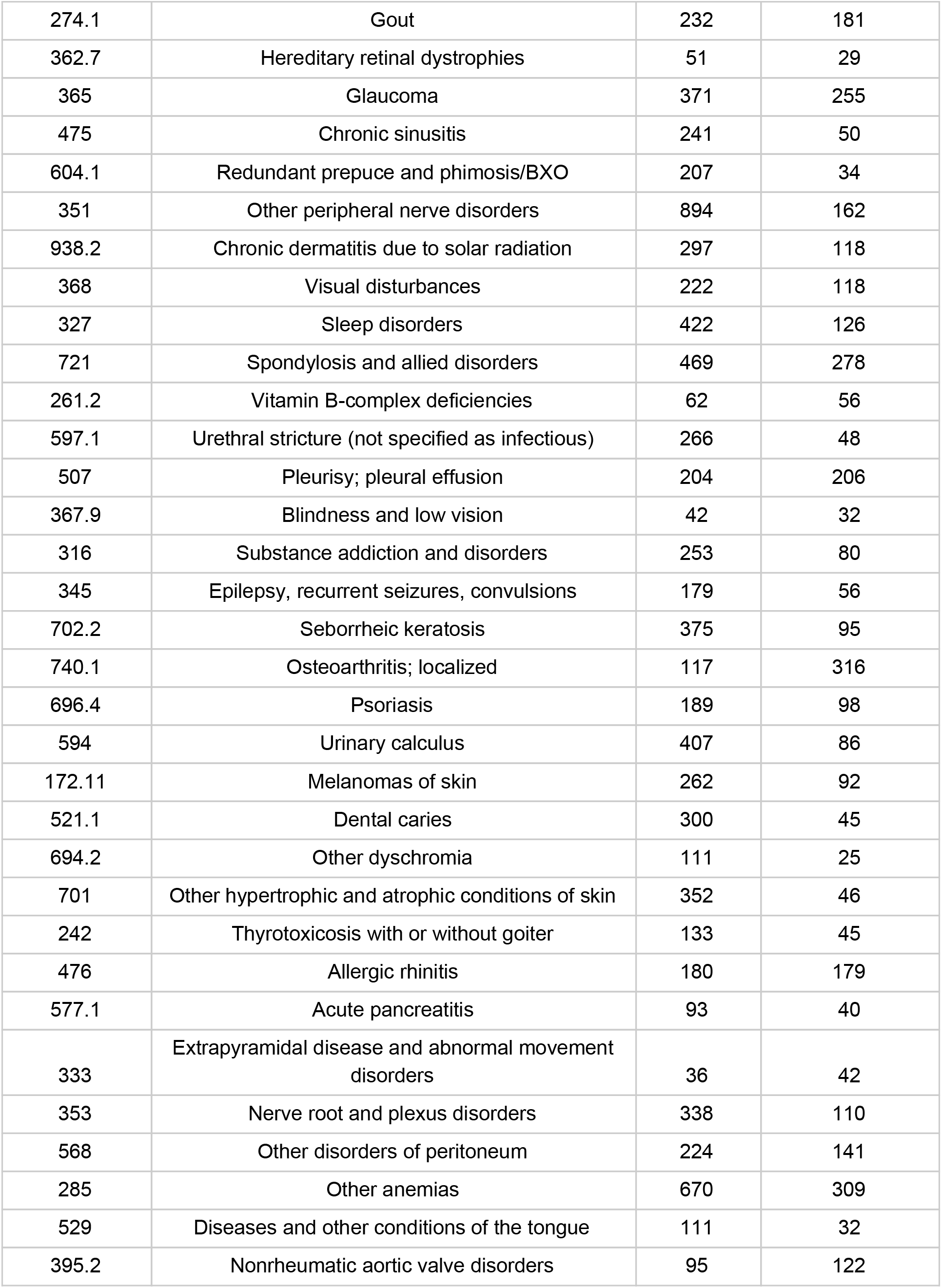
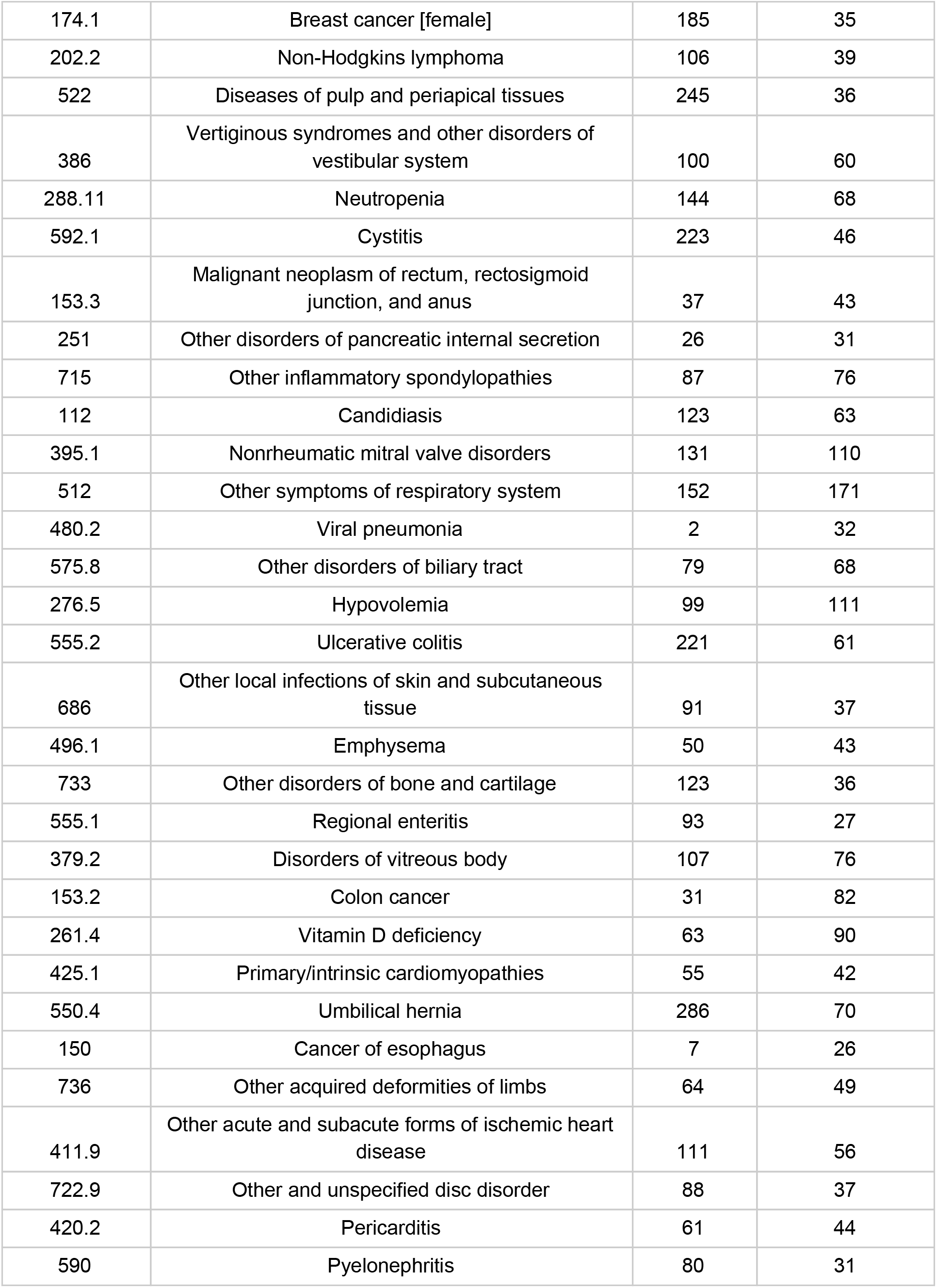
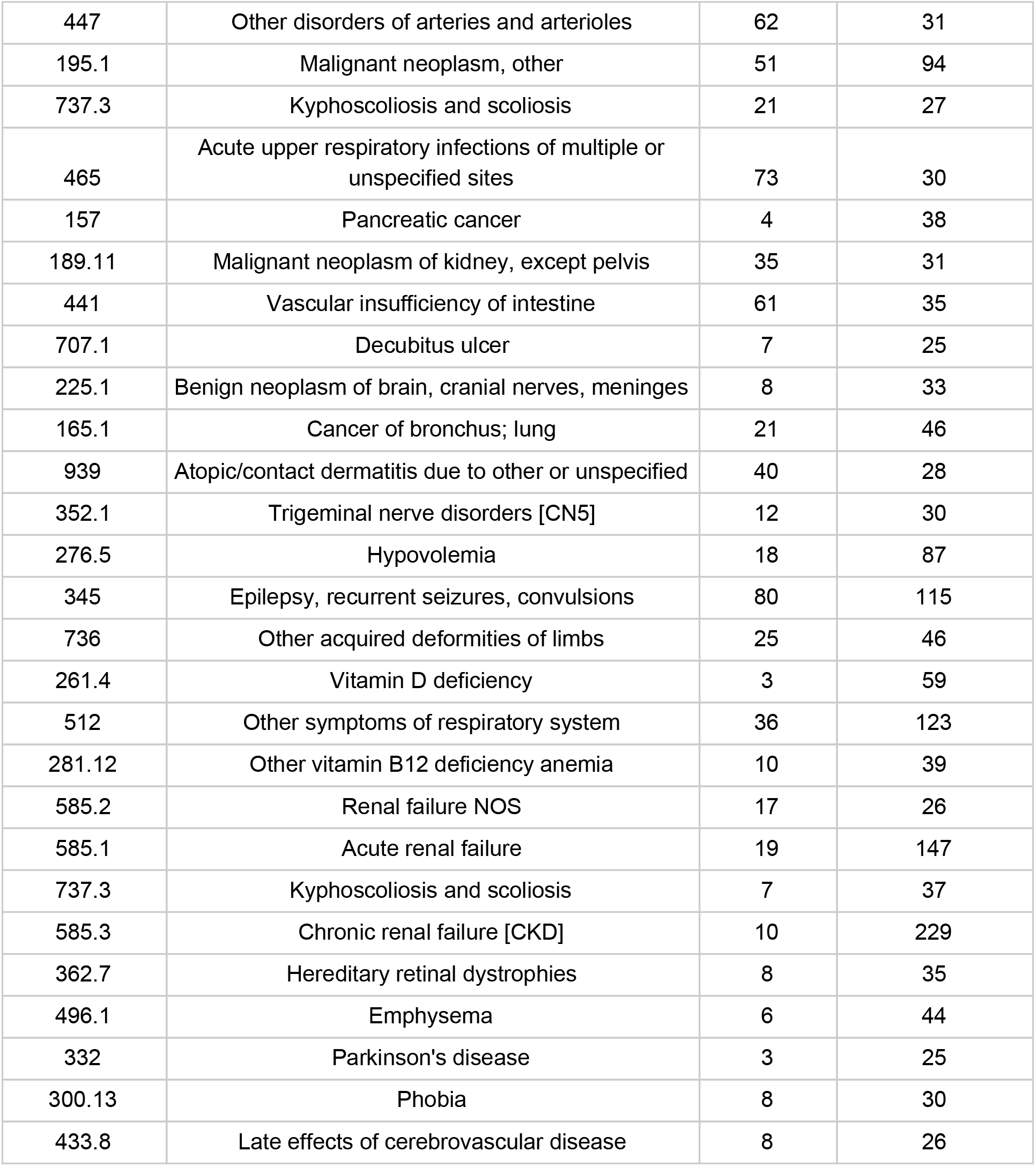
The PheWAS codes for which the CoxPH model was applied. We also show the number of people with the disease at baseline as well as the number of events during the follow up time.

#### Additional tables are in an additional sheet

Table S7: SNPs in 95% credible set. Where there is a secondary signal, the var_conditional column gives the ID of the SNP conditioned on.

Table S8: Standardized effect size and p-value at the lead SNP at each locus in this study and in each of the contributing studies. Values are also given for the lead SNP at the RAP1GAP locus identified in CHARGE.

Table S9: Genetic correlation between AAC and 785 complex traits calculated using LD Score Regression.

Table S10: Colocalization with expression in different tissues.

Table S11: Colocalization with complex traits in UK Biobank.

Table S12: Colocalization with cardiovascular traits from independent studies.

## Footnotes

1 http://biobank.ctsu.ox.ac.uk/crystal/crystal/docs/DXA_explan_doc.pdf

